# Dissecting the pleiotropic genetic architecture of suicide attempt, suicidal ideation, and thirteen correlated traits

**DOI:** 10.64898/2025.12.07.25341749

**Authors:** Alfonso Martone, Isabella Folego-Temoteo, Gabriel R. Fries, Anna R. Docherty, Allison E. Ashley-Koch, European College of Neuropsychopharmacology Network on Suicide Research and Prevention, Cibele E. Bandeira, Eugenio H. Grevet, Claiton H.D Bau, Janita Bralten, Chiara Fabbri, Alessandro Serretti, Diego L. Rovaris, Giuseppe Fanelli

## Abstract

Suicide attempts (SA) and suicidal ideation (SI) are major public health concerns with incompletely defined biology. We investigated shared genetic architecture between SA or SI and thirteen correlated phenotypes using genome-wide association study summary statistics from 46,350 to 975,353 individuals. Local Analysis of [co]Variant Association (LAVA) identified locus-specific genetic covariance, while conjunctional false discovery rate (conjFDR) analyses detected single-nucleotide polymorphysms jointly associated with suicide phenotypes and external traits. Functional annotation and enrichment analyses characterised pathways and tissue-specific expression. LAVA identified sixteen loci with significant local correlations, mapping to 493 genes. After conditioning on major depressive disorder and post-traumatic stress disorder, several locus–trait-pair correlations remained significant, including SI–ADHD, whose mapped genes were differentially expressed in hypothalamus, cortical regions, and peripheral tissues. Correlated loci implicated ion transport and transcriptional regulation. ConjFDR identified shared loci mapping 798 genes. These genes showed enrichment for pathways related to cell adhesion, neurogenesis, signal transduction, chromatin regulation, immune processes, and protein secretion. Stratified analyses showed SA–based pairs enriched for gene sets related to brain morphology, cognition, and sleep regulation, whereas SI–based pairs for neuroticism, body mass index, and gastrointestinal traits. Across pairs, shared loci displayed both concordant and discordant effect directions. Both SA and SI were enriched for gene sets involving glycine, serine, and threonine metabolism, systemic lupus erythematosus, and DNA damage- and telomere stress–induced senescence. Recurrently mapped genes exhibited region- and developmental stage–specific brain expression. These findings refine the genetic architecture of suicide and implicate neurodevelopmental, immune, metabolic, and chromatin-related mechanisms in suicidal thoughts and behaviours.

## 1. Introduction

Suicide-related phenotypes—including suicidal ideation (SI) and suicide attempt (SA), hereby jointly referred to as suicidal thoughts and behaviours [STBs])—represent a major public health challenge and contribute substantially to global morbidity and mortality (Suominen et al., 2004; World Health Organisation, 2023). More than 720,000 individuals die by suicide annually, accounting for approximately 10–15% of all SAs (Suominen et al., 2004; “WHO’s key facts about suicide,” n.d.). In the United States alone, 13.2 million individuals reported SI in 2022, with 1.9 million attempts and 49,000 resulting deaths (CDC, 2025). Together with major depressive disorder (MDD), SA is the strongest known predictor of subsequent death by suicide (Cavanagh et al., 2003; Franklin et al., 2017), but the biological mechanisms underlying both SI and SA remain insufficiently characterised. Although SI and SA are often conceptualised as points along a severity continuum (DiBlasi et al., 2021), emerging evidence indicates that they exhibit only partial overlap in clinical correlates and genetic liability (Klonsky et al., 2021a). Twin studies report heritability estimates for SI ranging from 36% to 47%, even after accounting for psychiatric comorbidities, indicating residual genetic contributions independent of co-occurring disorders (Fu et al., 2002). For SA, family- and twin-based heritability estimates range from 30% to 50% (Voracek and Loibl, 2007), with single-nucleotide polymorphism (SNP)-based heritability from genome-wide association studies (GWASs) ranging between 3.5% and 4.6% depending on phenotype definition and analytical strategy applied (Ruderfer et al., 2020a). For SI, recent GWAS estimated SNP-based heritability at 4.6% –unadjusted– and 2.3% when conditioned on post-traumatic stress disorder (PTSD)

(Ashley-Koch et al., 2023), with intermediate values when adjusting for MDD, bipolar disorder (BD), or schizophrenia (SCZ). While psychiatric diagnoses—particularly MDD—remain major risk factors for suicidality (Ruderfer et al., 2020b; Strawbridge et al., 2019), these estimates indicate that genetic risk for SI and SA is not fully explained by comorbid psychopathology, supporting the hypothesis of both shared and distinct biological mechanisms.

Epidemiological data indicate that approximately 80% of individuals who attempt suicide meet criteria for at least one psychiatric disorder at the time of the attempt (Davis et al., 2020; Nock et al., 2010). Yet, suicidal thoughts and behaviours are not confined to any single diagnosis and can be observed across nearly all major nosographic categories. The strongest associations are reported with MDD, but elevated risk is also reported in SCZ, autism spectrum disorder (ASD), BD, and attention-deficit/hyperactivity disorder (ADHD) (Docherty et al., 2023a; McCall and Black, 2013; Oquendo et al., 2024; Sanchez-Carro et al., 2023). These associations are mirrored at the genomic level: genetic correlation analyses have consistently shown shared polygenic contributions between suicide risk and a wide array of psychiatric traits (Bertolote and Fleischmann, 2002; Li et al., 2023). Importantly, suicidal phenotypes have also been linked to several non-psychiatric traits, including physical health, behavioural, and socio-demographic factors. Traits such as smoking behaviour, body mass index (BMI), chronic pain, and educational attainment (EA) have all been associated with suicide at both the phenotypic and genomic levels (Docherty et al., 2023b; McCall and Black, 2013; Mullins et al., 2022a; Sariaslan et al., 2022; Sher, 2024; Zinchuk et al., 2024). These findings suggest that suicide vulnerability extends beyond psychiatric nosology and may involve biological, behavioural, and socio-environmental pathways.

Despite the growing recognition of these associations, most prior research has focused on global genetic correlations, which provide genome-wide averages of polygenic overlap but do not account for regional specificity or the direction of effects at individual loci (Ruderfer et al., 2020a). A limitation of these analyses is that they do not identify the specific genomic regions contributing most to the shared genetic architecture of suicide and its associated traits. Local patterns of genetic sharing may reveal loci or regions where the direction of effect for suicide risk is concordant or divergent from that of psychiatric and non-psychiatric traits (Fanelli et al., 2025; Kontou and Bagos, 2024), providing novel insights into the biological mechanisms underlying these relationships.

To address these limitations, local genetic correlation methods, such as those implemented in Local Analysis of [co]Variant Association (LAVA), estimate regional genetic covariance across linkage disequilibrium (LD) semi-independent segments, allowing for the identification of trait-specific overlap and heterogeneity in directionality (Werme et al., 2022). Complementary pleiotropy-informed approaches, including the conjunctional false discovery rate (conjFDR), increase power to detect shared loci by leveraging cross-trait enrichment patterns and enable the identification of specific SNPs jointly associated with multiple traits (Andreassen et al., 2013; Smeland et al., 2020). These methods are well suited to investigate phenotypes like SI and SA, where genetic architecture is likely to involve both overlapping and distinct pleiotropic components with other traits.

In this study, we systematically assess local patterns of convergent or divergent genetic sharing between suicide phenotypes and other traits by integrating local genetic correlation and conjFDR methods. We focus on SI and SA and examine their locus-specific overlap with a curated set of psychiatric and non-psychiatric traits previously shown to be genetically correlated at the global level (Docherty et al., 2023a; Mullins et al., 2022b). Specifically, our objectives are to: (1) identify specific genomic regions contributing to the shared genetic liability between SI, SA and other psychiatric/non-psychiatric traits; (2) determine whether these loci or regions show concordant or discordant directions of effect; and (3) refine our understanding of shared biological mechanisms. By increasing the resolution at which genetic sharing is examined, our findings aim to inform suicide risk stratification and support future biomarker discovery and therapeutic development.

## 2. Experimental procedures

### 2.1. Input summary statistics of GWASs

We included in the analyses thirteen traits based on prior evidence of global genetic correlation with SA or SI (Ashley-Koch et al., 2023; Docherty et al., 2023b; Mullins et al., 2022b). The selected traits include: ADHD, ASD, BD, BMI, cigarettes per day (CPD), behavioural disinhibition (DSN), EA, ever smoker (ESMK), insomnia (INS), multi-site chronic pain (MSCP), neuroticism (NEU), risk-tolerance (RSKT), and SCZ. MDD and PTSD were included only for sensitivity analyses because they are highly collinear with suicidality at the genome-wide level: the largest meta-analyses report rg’s ∼0.8 for SA/SI with either MDD or PTSD (Ashley-Koch et al., 2023; Docherty et al., 2023b). For each phenotype, including SA and SI, we obtained summary statistics from the most recent and comprehensive GWAS available at the time of analysis. Quality control steps excluded variants considered ambiguous, duplicated, rare (MAF < 0.01), and with poor imputation quality (INFO < 0.8). Details of the GWAS sources and sample sizes for each phenotype are provided in Table 1.

**Table 1.** Summary statistics and phenotypes.

| Trait/disorder | Authors, year | PMID | Total N (N cases) |
| --- | --- | --- | --- |
| ADHD | Demontis et al., 2023 | 36702997 | 225 534 (38 691) |
| ASD | Grove et al., 2019 | 30804558 | 46 350 (18 381) |
| BD | Mullins et al., 2021 | 34002096 | 413 466 (41 917) |
| BMI | Yengo et al., 2019 | 30124842 | 456 426 (NA) |
| CPD | Saunders et al., 2022 | 30643251 | 618,489 (NA) |
| DSN | RK Linnér et al., 2019 | 30643258 | 315 894 (NA) |
| EA | Okbay et al., 2022 | 35361970 | 765 283 (NA) |
| ESMK | RK Linnér et al., 2019 | 30643258 | 518 633 (NA) |
| INS | Watanabe et al., 2022 | 35835914 | 386 988 (109 548) |
| MSCP | Johnston et al., 2019 | 31194737 | 412 985 (NA) |
| NEU | Nagel et al., 2018 | 29942085 | 390 278 (NA) |
| RSKT | RK Linnér et al., 2019 | 30643258 | 975 353 (NA) |
| SA | Docherty et al., 2023 | 30124842 | 815 178 (35 786) |
| SCZ | Trubetskoy et al., 2022 | 35396580 | 100 468 (39 910) |
| SI | Ashley-Koch et al., 2023 | 36940203 | 612 381 (99 814) |

ASD, Autism Spectrum Disorder; BD, Bipolar Disorder; BMI, Body Mass Index; CPD, Cigarettes per Day; DSN, Disinhibition; EA, Educational Attainment; ESMK, Ever Smoker; INS, Insomnia; MSCP, Multi-Site Chronic Pain; NEU, Neuroticism; RSKT, Risk Tolerance; SA, Suicide Attempt; SCZ, Schizophrenia; SI, Suicidal Ideation; NA, Non Applicable.

### 2.2. Local genetic correlation analyses - LAVA

We applied the LAVA (Local Analysis of [co]Variant Association) framework to estimate local genetic correlations (ρ) between suicide-related phenotypes and a range of psychiatric and non-psychiatric traits. LAVA partitions the genome into semi-independent loci based on linkage disequilibrium (LD) patterns, thereby enabling the identification of specific genomic regions that contribute to global genetic correlations. Each locus contains at least 1,000 SNPs with MAF > 0.01 across all summary statistics, resulting in 2,495 distinct loci. The partitioning script is publicly available at https://github.com/cadeleeuw/lava-partitioning (Werme et al., 2022).

To identify loci with evidence of local genetic signal, we first conducted univariate heritability tests for each phenotype within each locus. Only loci that showed significant local SNP-based heritability in both phenotypes (P < 0.00002, Bonferroni-corrected for 2,495 loci; 0.05/2,495) were included in the subsequent bivariate local genetic correlation analyses. Bivariate results were then adjusted for multiple testing using a second Bonferroni correction that accounted for the total number of loci tested across all phenotype pairs within each suicide phenotype. Because MDD and PTSD were later used as covariates in sensitivity analyses, their bivariate-tested loci were also included in the correction to ensure a consistent multiple-testing framework. To control for sample overlap, bivariate LD score regression intercepts were estimated and incorporated into the LAVA model (Bulik-Sullivan et al., 2015).

Next, we performed partial correlation analyses (ρₓᵧ|𝓏) within the LAVA framework to isolate independent local effects. We first selected loci showing significant local correlations with more than one phenotype pair. For each of these loci, we conducted pairwise conditional analyses, where the correlation between two phenotypes was conditioned on a third correlated phenotype to distinguish shared from independent effects. For example, if locus 32 showed significant correlations for both SA–ADHD and SA–BMI, we performed two conditional tests: (a) SA–ADHD conditioned on BMI, and (b) SA–BMI conditioned on ADHD.

We then focused on loci shared between suicide behaviors and either MDD or PTSD. Given the strong genome-wide correlations between these disorders and suicide traits, we performed partial genetic correlation analyses conditioning on MDD, PTSD, or both, for loci that were significantly associated (P< 0.05) with suicide behaviors, these disorders, and at least one additional target trait. This strategy enabled us to assess whether the associations with suicide phenotypes persisted after accounting for potential confounding effects of MDD and PTSD. Although suicide behaviors share substantial genetic overlap with both disorders, they are not entirely coextensive—highlighting the presence of genetic components specific to suicide traits. Only loci that remained significant after all conditioning scenarios were carried forward to the enrichment analyses described below.

### 2.3. Conjunctional false discovery rate (conjFDR) analyses

To identify SNPs jointly associated with SA or SI and each of the thirteen selected traits, we applied the conjFDR approach (Andreassen et al., 2013; Smeland et al., 2020). This method builds upon the condFDR framework, which boosts GWAS discovery by applying an empirical bayesian statistical framework, conditioning SNPs-associated p-values of a primary phenotype on SNPs’ nominal p-values from a secondary trait (Andreassen et al., 2013; Smeland et al., 2020). For a pair of phenotypes, two condFDR values can be computed for each SNP, conditioning the first trait on the second one and vice versa. The highest of these two condFDR values is then defined as the SNP’s conjFDR (Andreassen et al., 2013). For phenotypes with overlapping samples, such as SA, sample overlap correction was applied following the instruction the protocol reported by the developers (https://github.com/precimed/pleiofdr). The MHC region (chr6:26000000-34000000) was excluded from all conjFDR analyses, as recommended for this analytical framework due to its complex LD structure and potential to inflate pleiotropy estimates (Andreassen et al., 2013; Schwartzman and Lin, 2011). Quantile–quantile (Q–Q) plots were generated to evaluate polygenic enrichment across trait pairs.

Statistically significant associations (i.e., candidate SNPs) were identified with a maximum FDR of 5%. Independent significant SNPs were defined as candidate SNPs with a LD *r^2^* < 0.6 with each other, whereas a LD *r^2^* < 0.1 defined lead SNPs (Watanabe et al., 2017). Candidate SNPs in LD *r^2^*< 0.6 with a given lead SNP defined a genomic locus. Novel loci resulting from conjFDR analyses were defined as loci not identified in parent, single-trait GWASs. To evaluate the effect direction of each lead SNP identified with conjFDR, the corresponding Z-scores from the individual trait GWASs were extracted and the signs compared to see if the effect was discordant (opposite sign) or concordant (same sign). Finally, the identified shared genetic loci through the conjFDR approach were also compared to LAVA’s results by checking for genomic ranges overlap using the GenomicRanges R package (Lawrence et al., 2013).

### 2.4 Functional annotation, gene mapping, and gene set analysis

Functional annotation for the shared genetic loci identified in the previous steps was conducted with FUMA (Watanabe et al., 2017), incorporating annotations from the RegulomeDB (Boyle et al., 2012) and 15-core chromatin states (Kundaje et al., 2015), representing expression Quantitative Trait Loci (eQTL) and genomic regions’ accessibility, respectively. The MHC region (chr6:26000000-34000000) was excluded. To prioritize the most deleterious variants, a Combined Annotation Dependent Depletion (CADD) Score ≥ 12.37 was used.

To retrieve the list of genes located within each locus identified in the LAVA analysis that survived all partial analysis, we utilised the NCBI Genome Data Viewer (GDV). Subsequently, the GENE2FUNC module from FUMA was employed to assess tissue-specific expression patterns and to explore potential enrichment in biological pathways and gene sets for the genes mapped to each locus.

Three different SNP-to-gene mapping strategies were used for conjFDR significant results, including (1) positional mapping (with a 10kb window size), (2) 3D chromatin interaction mapping, and (3) eQTL mapping. Gene set enrichment and tissue-specific expression analyses were performed using Multi-marker Analysis of Genomic Association (MAGMA), as implemented in FUMA. Four reference datasets were used to assess spatial and temporal patterns of gene activity. These included 54 distinct tissue types and 30 aggregated tissue categories from GTEx v8, and 29 developmental stages alongside 11 broader developmental windows from BrainSpan. The prenatal stages represented in BrainSpan span from 8 to 37 post-conception weeks, allowing detailed mapping of gene expression trajectories from early neurodevelopment through late gestation.

## 3. Results

### 3.1. Local genetic correlations between suicidal and other psychiatric and non-psychiatric phenotypes

#### 3.1.1 Global genetic correlations and locus-level selection

We reexamined the genetic correlations with LD Score Regression (LDSC) among STBs and target phenotypes using the latest GWAS summary statistics. Our findings were consistent with previous published findings (Docherty et al., 2023b), as summarised in Supplementary Figure 1 and Table S1. In the univariate LAVA analysis, 1,073 loci exhibited significant local SNP-based heritability for SA and at least one other tested phenotype, while 3,528 loci met this criterion for SI and were considerate in Bonferroni correction (P < 0.05/1,073 = 0.00004 for SA; P < 0.05/3,528 = 0.00001 for SI). Only these loci were retained for subsequent bivariate analyses (Tables S2–S27).

#### 3.1.2 Significant local genetic correlations for SA and SI

Among the 925 SA–associated loci, 15 demonstrated significant local genetic correlations with other traits (Table S28). These included 3 loci with ADHD, also 3 with BD, 2 each with DSN, EA and SCZ, and 1 locus each with BMI, ESMK and NEU. The top correlations primarily observed were between SA and DSN at locus chr11:28577719-29530460 (ρ=0.99, P=1.3e-4), followed by ESMK at locus chr6:25684630-26396200 (ρ=0.89, P=2.8e-2), and EA at locus chr1:151146408-152535630 (ρ= -0.89, P=1.5e-3), respectively.

For SI, 13 out of 3,528 tested loci exhibited significant local genetic correlations with other phenotypes (Table S28). In regards to SI-EA, 5 loci were significant: chr2:103305258-105120578 (ρ= -0.47, P=1.7e-2), chr5:166863414-168342743 (ρ= -0.72, P=4.6e-2), chr9:128785783-129617771 (ρ= -0.57, P=3.1e-2), chr12:120567741-121817509 (ρ= -0.71, P=4.9e-3) and chr17:45883902-47516224 (ρ= -0.74, P=1.1e-3). With SCZ, there are 3 correlations, on loci chr4:113916003-115306149 (ρ=0.73, P=1.9e-2), chr6:25684630-26396200 (ρ=0.71, P=9.2e-4) and loci chr11:112755447-113889019 (ρ=0.42, P=3.0e-2). Two loci were associated between SI and ADHD, on loci chr10:133818037-134856054 (ρ=0.83, P=1,3e-2) and chr11:28577719-29530460 (ρ=0.72, P=4e-3), as well as with NEU on loci chr11:112755447-113889019 (ρ=0.51, P=1.7e-4), chr18:52512524-53762996 (ρ=0.72, P=2.5e-2). Lastly, there were only 1 locus for SI and ESMK (ρ=0.86, P=3.2e-4).

Three loci demonstrated local pleiotropy across multiple traits. At locus chr1:43512670–45167235, SA was associated with both ADHD and EA in opposite directions (ρ=0.71, P=1.8e-3 and ρ= -0.56, P=1.8e-3; respectively). In contrast, for SI, locus chr11:28577719–29530460 showed correlations with both ADHD and ESMK in the same direction (ρ=0.72, P=4e-3 and ρ=0.86, P=3.2e-4), as did locus chr11:112755447-113889019 for SI with NEU and SCZ (ρ=0.51, P=1.7e-4 and ρ=0.42, P=3e-2).

#### 3.1.3 Conditional analyses and robustness to MDD/PTSD

Conditional analyses were then performed to assess trait-specific effects at these loci. Most correlations lost significance after conditioning (Table S29). For instance, the correlation between SA and ADHD at locus chr1:43512670-45167235 became non-significant when conditioned on EA (ρ=0.54, P=0.12), as well as the correlation between SA and EA when conditioned on ADHD (ρ=0.13, P=0.74). These findings suggest that other factors may be driving the observed correlations between these phenotypes within this region. Only one remained significant, on locus chr11:112755447-113889019 (ρ=0.7, P=1.8e-3), indicating that the correlation between SI and NEU is independent of SCZ.

Given the high correlation between STBs with MDD and PTSD, we identified significant loci (P < 0.05) between them (Tables S30-33) and conditioned the overlapped correlated loci with the same loci from the main analysis to account for MDD and PTSD confounding effects. In this step 5 loci lost significance, which indicates that MDD and/or PTSD might better explain the correlation between STBs and these traits on specific loci (Table S29). In loci chr6:30070718-30715006 for SA and BD, loci chr7:113339387-115321301 for SA and DSN and loci chr18:52512524-53762996 for SI and NEU, both PTSD and MDD influenced the correlation. The correlation was influenced only by PTSD in locus chr2:103305258-105120578 for SI and EA, and in locus chr11:112755447-113889019 for SI and NEU, while MDD influenced locus chr11:28577719-29530460 correlation between SA and DSN. In total, 17 correlations persisted significantly even after controlling for all potential confounding effects (Table 2 and Fig 1). The full list of genes within each locus is provided in Table S34 and was used for the enrichment analysis of LAVA results.

**Figure 1.**
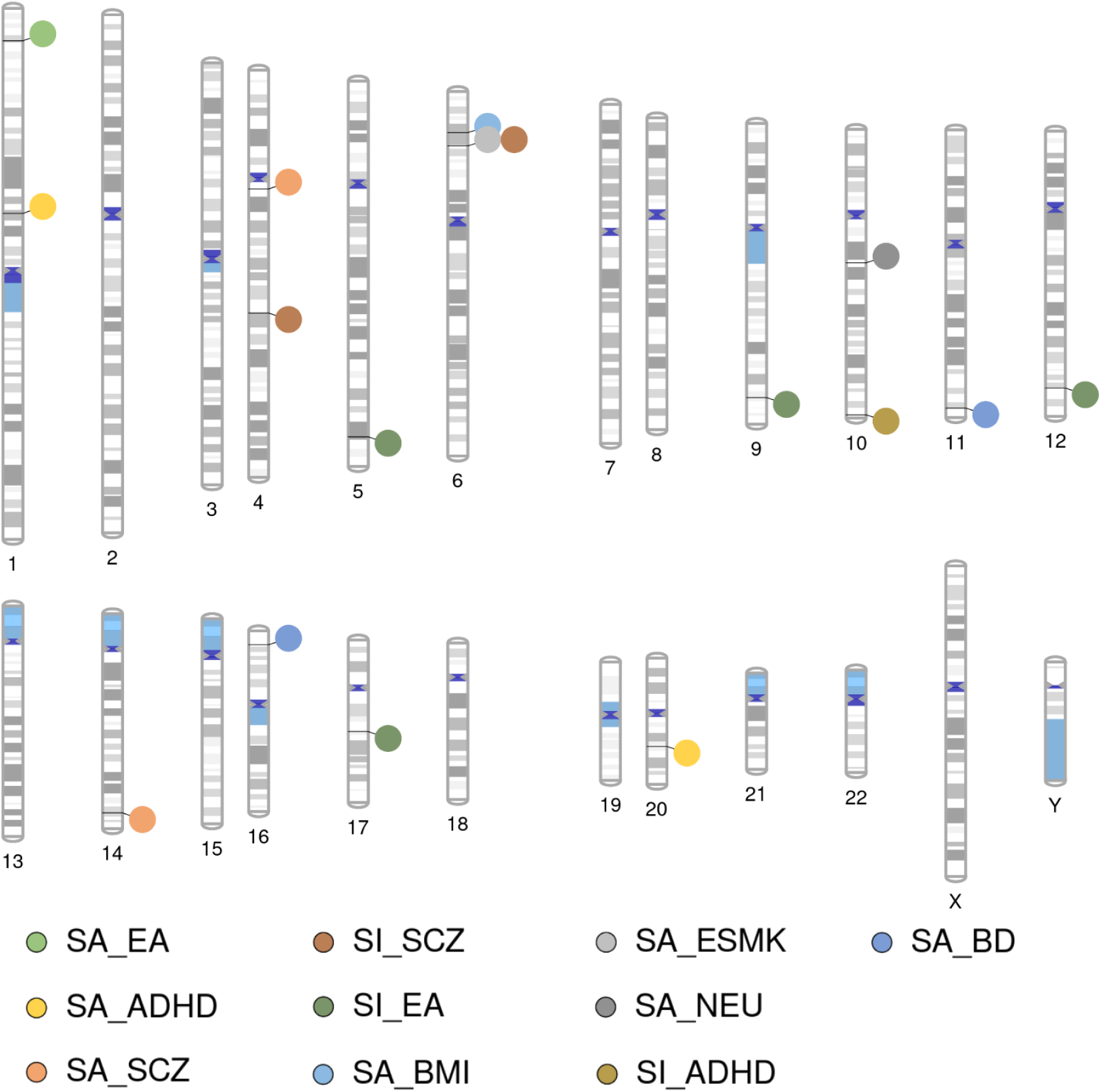
Chromosome view of local, pairwise genetic correlations between suicide attempt/ideation vs the other 13 correlated traits. ADHD, Attention Deficit and Hyperactivity Disorder; BD, Bipolar Disorder; BMI, Body Mass Index; EA, Educational Attainment; ESMK, Ever Smoked; NEU, Neuroticism; SA, Suicide Attempt; SCZ, Schizophrenia; SI, Suicidal Ideation.

**Table 2.** Results from local correlation analysis after controlling for confounding effects.

| Trait 1 | Trait 2 | Locus | Chr | Start | Stop | N SNPs | Local rg ($\rho_0$) | P | P-corr* | z | Partial rg ($\rho_p$) | P |
| --- | --- | --- | --- | --- | --- | --- | --- | --- | --- | --- | --- | --- |
| SA | ADHD | 84 | 1 | 96147577 | 97721186 | 2815 | 0.75 | 1.4e-5 | 0.013 | MDD | 0.73 | 0.001 |
|  |  | 2408 | 20 | 44072211 | 45673603 | 3248 | 0.66 | 4.3e-5 | 0.040 | PTSD | 0.51 | 0.005 |
|  | BD | 1735 | 11 | 130500784 | 131424361 | 1905 | 0.68 | 3.9e-5 | 0.036 | MDD | 0.64 | 0.002 |
|  |  | 2109 | 16 | 7051430 | 7704354 | 2610 | 0.73 | 4.5e-5 | 0.041 | MDD | 0.78 | 5.3e-4 |
|  | BMI | 943 | 6 | 19449153 | 20418020 | 830 | 0.72 | 6.4e-6 | 0.006 | - | - | - |
|  | EA | 110 | 1 | 151146408 | 152535630 | 1844 | -0.89 | 1.4e-6 | 0.001 | - | - | - |
|  | ESMK | 950 | 6 | 25684630 | 26396200 | 1737 | 0.89 | 2.6e-5 | 0.024 | MDD | 0.89 | 0.018 |
|  | NEU | 1544 | 10 | 62626466 | 64069687 | 2712 | 0.78 | 1.4e-5 | 0.004 | - | - | - |
|  | SCZ | 652 | 4 | 55681702 | 57103100 | 3214 | 0.78 | 4.0e-5 | 0.037 | - | - | - |
|  |  | 2026 | 14 | 99474534 | 100786189 | 2514 | 0.66 | 3.0e-5 | 0.028 | - | - | - |
| SI | ADHD | 1608 | 10 | 133818037 | 134856054 | 2341 | 0.83 | 3.2e-6 | 0.011 | MDD | 1 | 1.4e-05 |
|  | EA | 915 | 5 | 166863414 | 168342743 | 3137 | -0.72 | 1.1e-5 | 0.039 | MDD | -0.70 | 0.002 |
|  |  | 1472 | 9 | 128785783 | 129617771 | 2038 | -0.57 | 7.6e-6 | 0.027 | PTSD | -0.48 | 0.008 |
|  |  |  |  |  |  |  |  |  |  | MDD | -0.45 | 0.015 |
|  |  | 1849 | 12 | 120567741 | 121817509 | 3124 | -0.71 | 1.2e-6 | 0.004 | PTSD | -0.58 | 7.1e-05 |
|  |  |  |  |  |  |  |  |  |  | MDD | -0.56 | 0.004 |
|  |  | 2209 | 17 | 45883902 | 47516224 | 3414 | -0.74 | 3.1e-7 | 0.001 | MDD | -0.72 | 1.3e-05 |
|  | SCZ | 701 | 4 | 113916003 | 115306149 | 2826 | 0.73 | 4.6e-6 | 0.019 | - | - | - |
|  |  | 950 | 6 | 25684630 | 26396200 | 1803 | 0.71 | 2.2e-7 | 0.001 | - | - | - |

\*Bonferroni correction for all bivariate tests (NSA = 1,083; NSI= 4,112).
ADHD, Attention Deficit and Hyperactivity Disorder; BD, Bipolar Disorder; BMI, Body Mass Index; Chr, Chromosome; EA, Educational Attainment; ESMK, Ever Smoker; N SNPs, Number os SNPs located within locus region; NEU, Neuroticism; P, p-value; P-corr, p-value corrected with Bonferroni correction for multiple testing; RG, genetic correlation, SA, Suicide Attempt; SCZ, Schizophrenia; SI, Suicidal Ideation; SNP, Single Nucleotide Polymorphism; z, confounding variable controlled; -, not tested.

#### 3.1.4 Functional annotation of LAVA loci

Using the GENE2FUNC module on the FUMA platform, we identified differential gene expression in at least 10 different tissues for the genes located within loci associated with specific phenotype pairs (Supplementary Figures S2–S5). As expected, several genes involved in the connection between phenotypes were differentially expressed in brain tissues. Genes located within loci mediating the SI–ADHD association (e.g., JAKMIP3, DPYSL4, INPP5A, CFAP46) were up-regulated in four brain regions: hypothalamus, frontal cortex, cortex, and anterior cingulate cortex (Supplementary Figure 4B). Additionally, some genes were differentially expressed in non-neural tissues: genes located within SI–ADHD loci were down-regulated in blood vessels, esophagus, and aorta (Supplementary Figure 4A–B). Furthermore, SA–EA genes were up-regulated in the skin, including suprapubic (non–sun-exposed) and lower leg (sun-exposed) regions (Supplementary Figure 2A–B). SA–ESMK genes showed increased expression in liver, small intestine, and colon (Supplementary Figure 3), while SI–EA genes were up-regulated in the transverse and sigmoid colon, small intestine, stomach, and kidney cortex (Supplementary Figure 3A–B).

In addition, several gene sets were identified across phenotype pairs (Supplementary Figures S6–S9). Notably, genes located within SI–EA loci were enriched for gene sets associated with embryonic development (e.g., embryonic organ development, rhombomere development), morphogenesis (e.g., cranial nerve morphogenesis, embryonic organ morphogenesis), regulation of biological processes (e.g., negative regulation of lamellipodium assembly, positive regulation of cytosolic calcium ion transport), cellular components (e.g., chromatin, chromosome, ribonucleoprotein complex), and molecular functions, including DNA-binding transcription factor activity, transcription regulator activity, and sequence-specific DNA binding (Supplementary Figure 8). Additionally, genes within SA–ESMK and SI–SCZ loci were enriched for transport-related processes, such as urate and phosphate ion transport, as well as transmembrane transport activity, including sodium, metal, monoatomic ion, salt, and inorganic molecular entity transport (Supplementary Figures 7 and 9).

### 3.2. Cross-trait genetic associations identified via conjunctional FDR analyses

#### 3.2.1 Polygenic enrichment and jointly associated loci

Results from the conjFDR analysis are summarised in Table 3 and Supplementary Table S35, with top loci for each phenotype pair shown in Table 4. Polygenic enrichment was observed for most SA phenotype pairs, with the exception of SA–EA and SA–INS. This was evident from Q–Q plots, which showed some degree of upward and leftward deviation when conditioned for progressively smaller p-value thresholds (Supplementary Figures S10-S35). Q–Q plots for SI–based pairs exhibited similar features but with smaller deviations. Notably, SI–EA, as SA–EA and SA–INS above, displayed no relevant deviation from the expected distribution, indicating that no polygenic enrichment was found for these pairs. These results are consistent with the findings in terms of novel identified loci (Table 3) where none was found for SA–EA and SI–EA. Among the other trait combinations, the number of jointly associated new loci ranged from 1 (SA–ASD) to 33 (SA–DSN) for SA–based pairs, and from 3 (SI–INS) to 29 (SI–ADHD) for SI–based pairs.

**Table 3.**
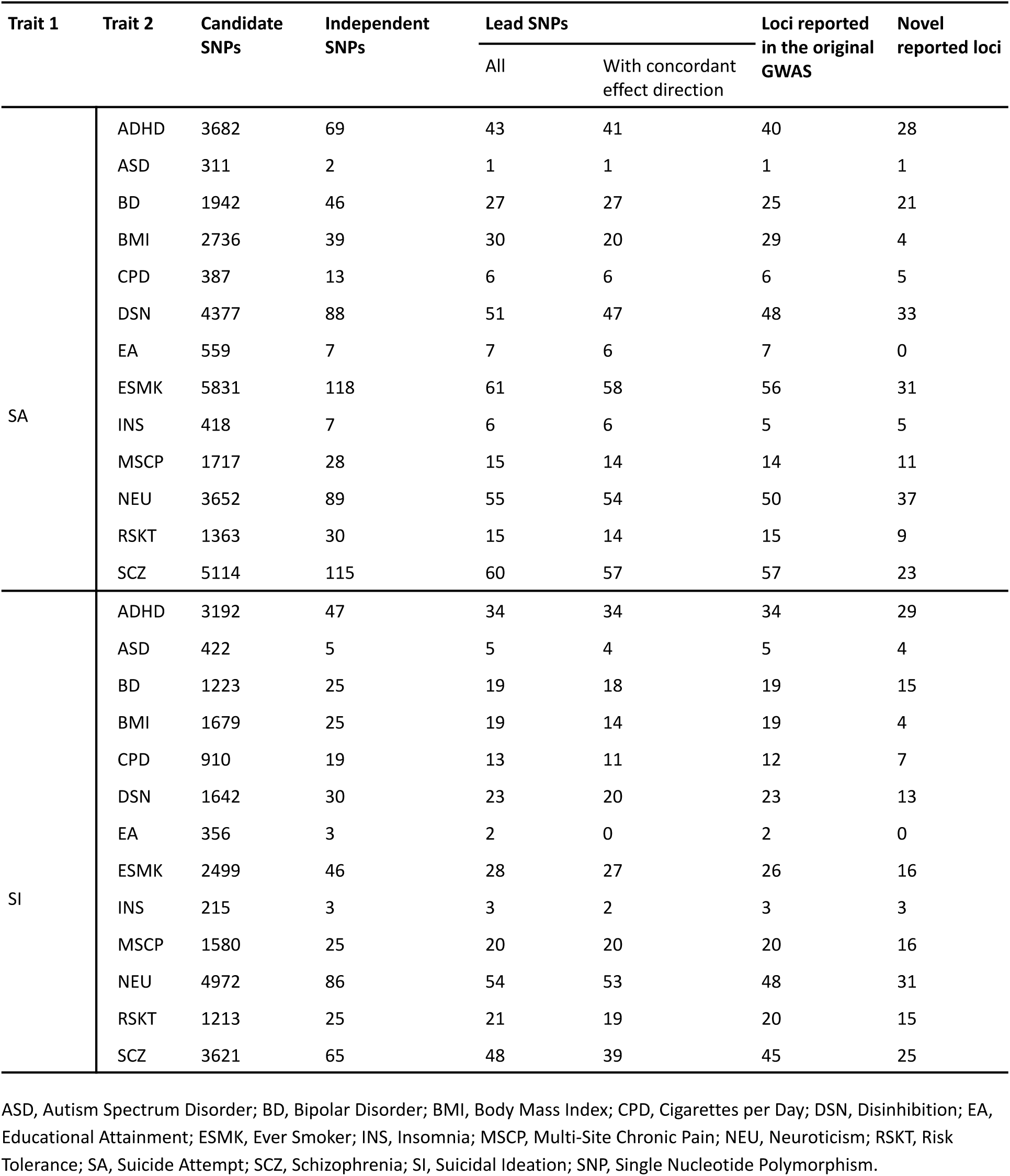
Overview of ConjFDR results.

**Table 4.** ConjFDR top loci (FDR < 0.001) per trait-pairs and relative mapped genes.

| SNP | Trait pair<br>(ED trait 1 / ED trait 2) | Mapped gene(s) |
| --- | --- | --- |
| rs3791129 | SA-ADHD (+ / +) | *ARTN, ST3GAL3, B4GALT2, CCDC24, SLC6A9, ATP6V0B, RP11-7O11.3, IPO13* |
| rs2503185 | SA-BMI (- / -) | - |
| rs35869525 | SA-BD (- / -) | Several genes of different families, like *BTN* (MHC-like genes), *HIST*, *ZSCAN* |
| rs8039305 | SA-BD (- / -) | *FES, FURIN* |
| rs9641538 | SA-BD (+ / +) | - |
| rs4630328 | SA-CPD (- / -) | *DRD2* |
| rs62474713 | SA-DIS (+ / +) | - |
| rs2503185 | SA-DIS (- / -) | - |
| rs17514846 | SA-DIS (- / -) | - |
| rs2582897 | SA-DIS (- / -) | - |
| rs2503185 | SA-ESMK (- / -) | - |
| rs12798900 | SA-NEU (+ / +) | - |
| rs35526527 | SA-NEU (- / -) | Several *BTN\**, *OR\**, *ZSCAN\**, and *HIST\** genes |
| rs4275159 | SA-RSKT (+ / +) | - |
| rs6224 | SA-SCZ (- / -) | *FES, FURIN* |
| rs4245150 | SA-SCZ (- / -) | *DRD2* |
| rs35869525 | SA-SCZ (- / -) | Several genes of different families, like *BTN* (MHC-like genes), *HIST*, *ZSCAN* |
| rs10835363 | SI-ADHD (- / -) | - |
| rs1823673 | SI-BMI (- / -) | - |
| rs7200879 | SI-BMI (- / +) | *FBXL19, STX1B, HSD3B7, SETD1A, STX4, KAT8, RNF40, ZNF646, VKORC1, ORAI3, RIM72, PRSS36, C16orf93, RP11-196G11.2, AC135048.13, RP11-388M20.9, INO80E* |
| rs1836798 | SI-DIS (+ / +) | - |
| rs10767730 | SI-DIS (- / -) | - |
| rs264921 | SI-ESMK (- / -) | - |
| rs1836798 | SI-ESMK (+ / +) | - |
| rs10835363 | SI-ESMK (+ / +) | - |
| rs62098014 | SI-MSCP (+ / +) | - |
| rs56403421 | SI-NEU (+ / +) | *CTD-2171N6.1* |
| rs17503448 | SI-NEU (+ / +) | - |
| rs1116313 | SI-NEU (+ / +) | - |
| rs56403421 | SI-SCZ (+ / +) | *CTD-2171N6.1* |

ASD, Autism Spectrum Disorder; BD, Bipolar Disorder; BMI, Body Mass Index; CPD, Cigarettes per Day; DSN, Disinhibition; EA, Educational Attainment; ED, Effect Direction; ESMK, Ever Smoker; INS, Insomnia; MSCP, Multi-Site Chronic Pain; NEU, Neuroticism; RSKT, Risk Tolerance; SA, Suicide Attempt; SCZ, Schizophrenia; SI, Suicidal Ideation; SNP, Single Nucleotide Polymorphism.

Cross-trait associations—defined by jointly associated loci (conjFDR < 0.05), independent significant SNPs, and lead SNPs—were identified for SA–ESMK, SA–SCZ, SI–NEU, SA–DSN, SA–ADHD, SA–NEU, and SI–SCZ. Among all trait pairs, SI–INS yielded the fewest significant loci (Table 3, Figure 2).

**Figure 2.**
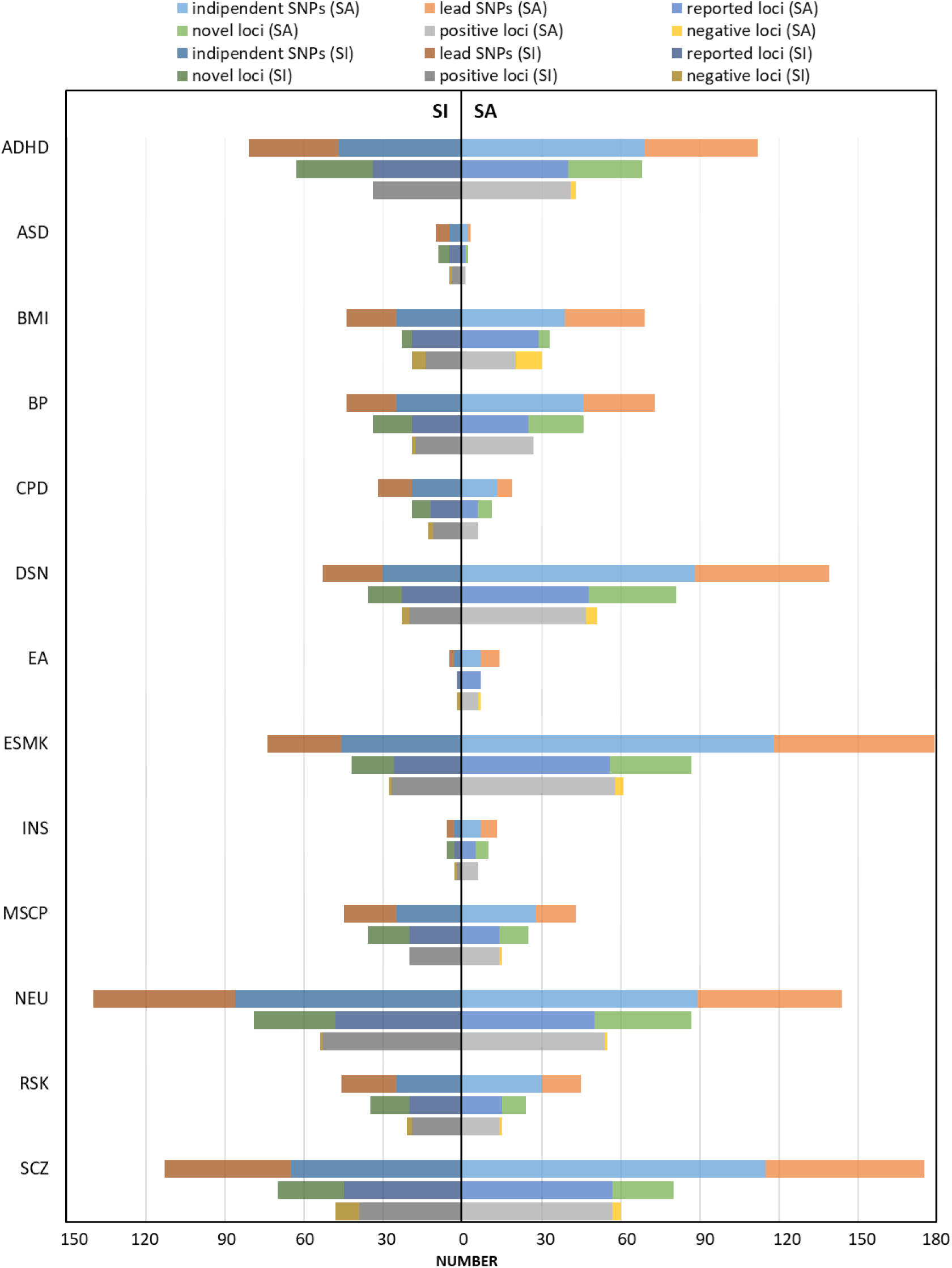
Overview of ConjFDR results. Independent SNPs: number of independent SNPs significatively associated with both traits; lead SNPs: number of lead SNPs; negative loci: number of loci with opposite effect directions in the two traits; novel loci: number of loci significantly associated with both traits that were not significant in the original GWAS; positive loci: number of loci with the same effect directions in the two traits; reported loci: number of loci significantly associated with both traits. ADHD, Attention Deficit and Hyperactivity Disorder; ASD, Autism Spectrum Disorder; BD, Bipolar Disorder; CPD, Cigarettes per Day; DSN, Disinhibition; EA, Educational Attainment; ESMK, Ever Smoker; INS, Insomnia; MSCP, Multi-Site Chronic Pain; NEU, Neuroticism; RSKT, Risk Tolerance; SA, Suicide Attempt; SCZ, Schizophrenia; SI, Suicidal Ideation.

Overall, SI–based analyses yielded fewer jointly associated loci than SA–based comparisons, with the exception of the CPD and NEU pairs, where SI results surpassed those for SA.

#### 3.2.2 Effect directions, overlap with LAVA, and functional characterisation

Across phenotype pairs, the majority of lead SNPs exhibited a positive effect direction based on their individual trait z-scores (Table 4). When assessing cross-trait concordance, however, all pairs included a subset of discordant SNPs—defined as those with opposite effect directions between the two traits—ranging from 2% to 30% (Table 4). Complete concordance was observed for SA–ASD, SA–BD, SA–CPD, SA–INS, SI–ADHD, and SI–MSCP (all with positive effects in both traits), and for SI–EA (all with negative effects).

Of the 484 unique lead SNPs, only two (rs6593600, rs12888948) overlapped with genomic regions showing significant local genetic correlations in the same phenotype pairs after correcting for MDD and PTSD, corresponding to the loci chr1:96147577-97721186 for SA-ADHD and chr14:99474534-100786189 for SA-SCZ, respectively (Table 5).

**Table 5.**
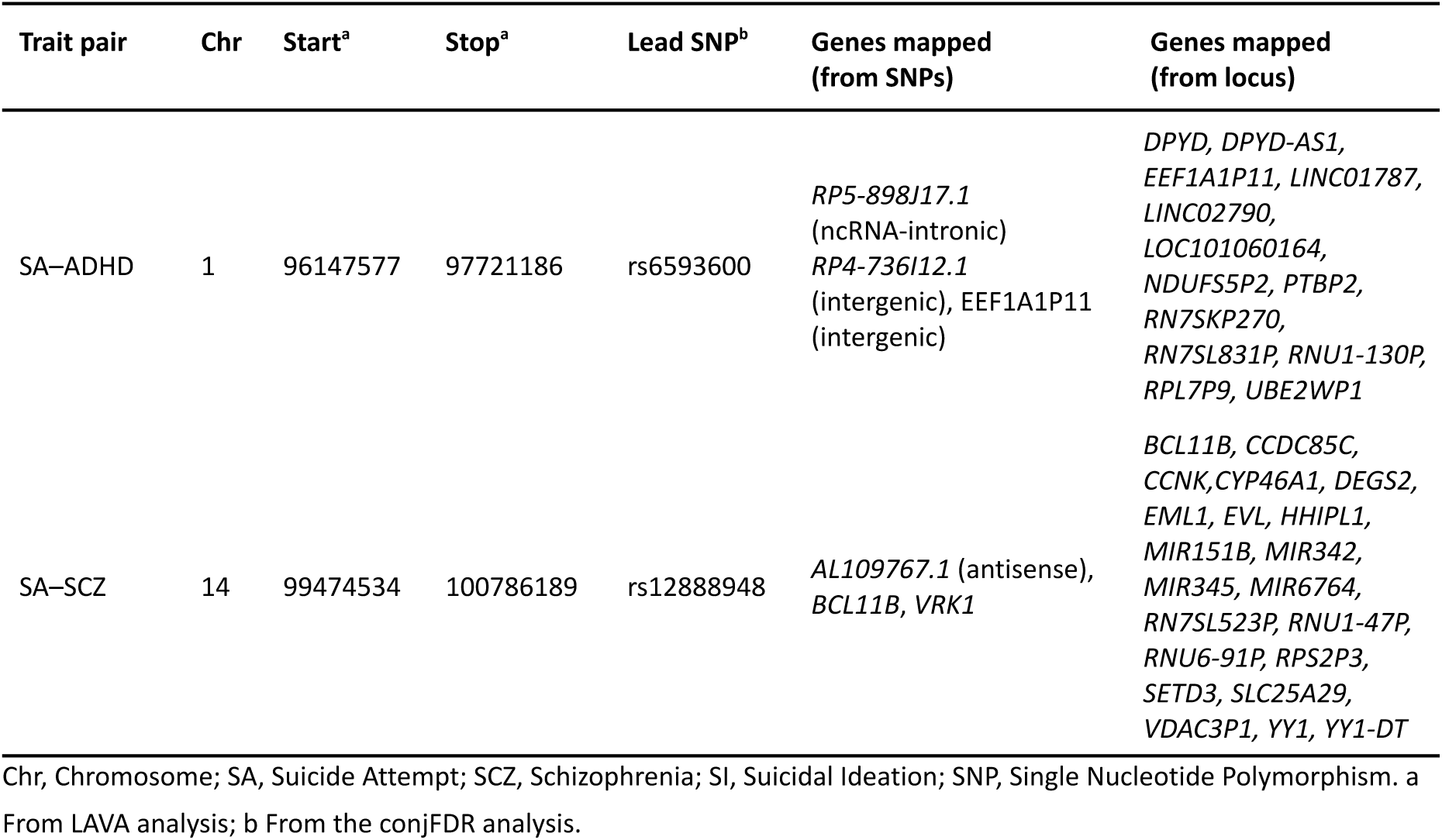
Overlapping results between conjFDR and LAVA.

A substantial proportion of SNPs identified through conjFDR analysis exhibited functional relevance, with 47% of mapped variants having a CADD score ≥ 12.37, indicating a deleterious potential. To further characterize the shared genetic architecture between suicidal and the other selected phenotypes, we mapped genes to the identified loci and examined their functional and biological relevance.

A total of 798 unique genes were mapped to SNPs identified through conjFDR analysis, 65.4% of which were protein-coding, 12.1% pseudogenes, 7.8% antisense RNA, and 7.2% long non-coding RNAs (lncRNAs) (Figure 3). Of these, 482 genes were mapped to loci identified in the analyses including SA and 421 to the loci identified in the analyses with SI.

**Figure 3.**
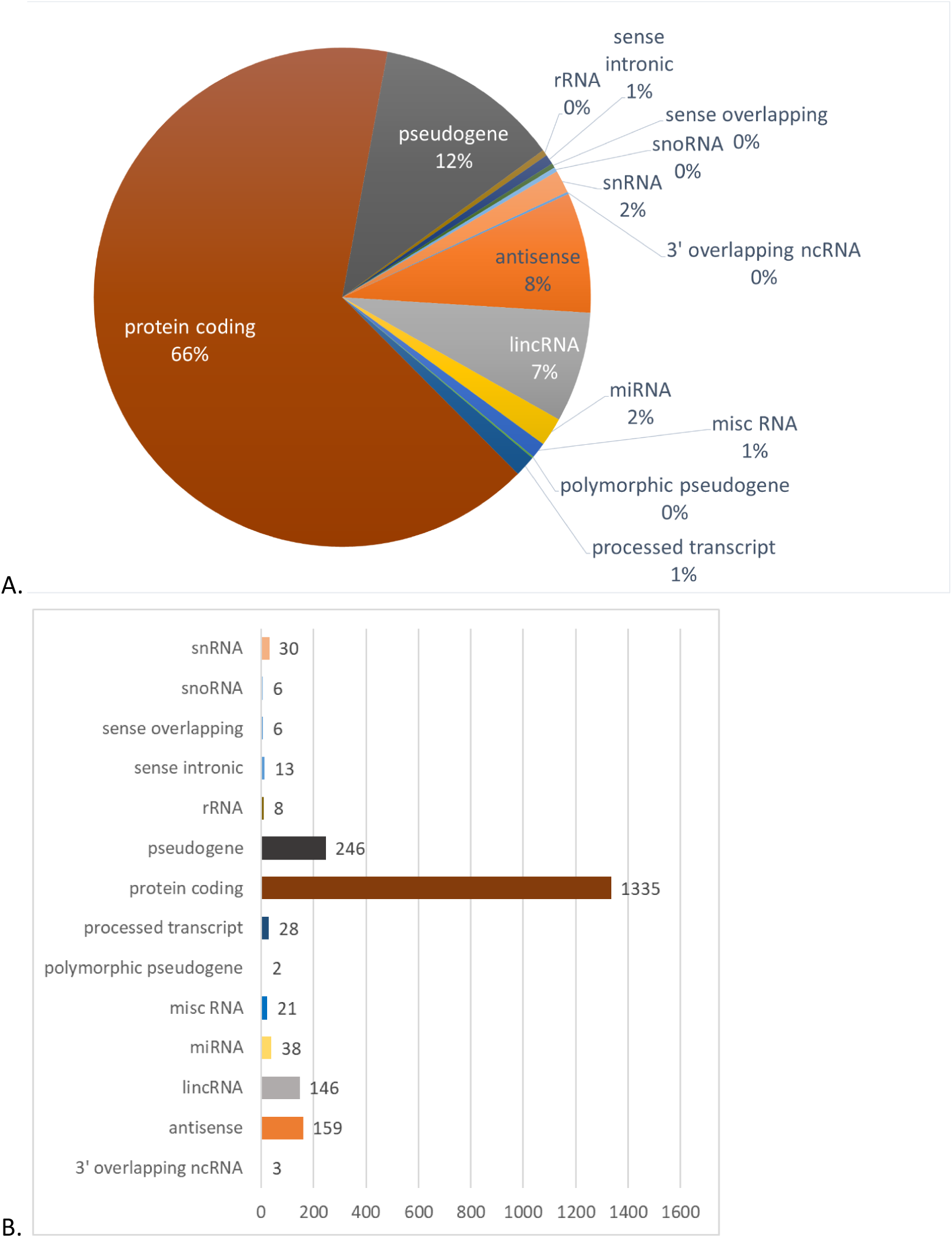
Gene type. A. Overall gene types mapped from ConjFDR results expressed as percentages. B. Overall gene types mapped from ConjFDR results in absolute numbers. lincRN, long non-coding RNA; miRNA, micro RNA; misc RNA, miscellaneous RNA; ncRNA, non-coding RNA; rRNA, ribosomal RNA; snoRNA, small nucleolar RNA.

In the SA subset, the most gene-rich trait pairs were SA–SCZ (217 genes, 154 protein-coding), SA–NEU (155, 109 protein-coding), and SA–DSN (135, 89 protein-coding). Fewer genes were mapped for SA–ASD (11, 3 protein-coding), SA–INS (10, 3 protein-coding), and SA–CPD (3, 2 protein-coding). Recurrently implicated genes in at least 5 phenotype pairs included those involved in cell adhesion, cell migration, and neurite growth (e.g. *NCAM1, LSAMP*), signal transduction (e.g. *FES, FAM212A/INKA1*), preprotein activation and secretion (e.g. *FURIN*), and transcriptional regulation (e.g. *MED27*) (Table S36). Other recurrent genes across SA-based pairs included *AMT*, encoding a glycine cleavage system component, and *ARTN*, encoding a ligand of the glial cell line-derived neurotrophic factor (GDNF) family (Table S36). With regard to SI, the phenotypes that shared more genes with SI were NEU (198 genes, 124 protein-coding), SCZ (157, 97 protein-coding), and ADHD (114, 84 protein-coding), while INS showed the lowest number of jointly associated genes (14 overlapping genes, 8 protein-coding). No gene was mapped to the SI–ESMK trait pair.

Notably, some of the most recurrent genes across SI pairs are in common with those mapped for phenotype pairs including SA; these include *NCAM1*, *FURIN* and *FES*. Other recurrent genes encode for zinc-finger proteins, as well as for proteins involved in transcription (*MAFK*), mitosis (*MAD1L1*), and cell cycle control and DNA repair (*FTSJ2*, also known as *MRM2*).

Gene set enrichment analysis identified 640 unique gene sets —525 for SA-based analyses and 276 for SI-based analyses. Enrichment was observed for genes implicated in brain morphology, subcortical volumes, cortical surface area, and psychiatric phenotypes including ASD, SCZ, anorexia nervosa, mood instability, risk-taking behaviour, age at first intercourse, and NEU. Additional enrichment was seen for gene sets involved in transcription factor targets, immune response, and other cellular processes (Table S37). Extending the gene set enrichment results, eight SA trait pairs (i.e., SA–ADHD, SA–BMI, SA–DSN, SA–EA, SA–ESMK, SA–MSCP, SA–NEU, and SA–SCZ) showed enrichment in genes located in chr3p21, whereas six SI trait pairs (i.e., SI–ADHD, SI–ASD, SI–DSN, SI–NEU, SI–RSKT, and SI–SCZ) exhibited enrichment for genes located in chr17q21. Notable differences emerged among the two subgroups of SA vs SI trait pairs; SA– trait pairs were predominantly enriched for genes related to brain morphology, psychiatric traits, cognitive function, and sleep regulation, whereas SI– trait pairs showed greater enrichment in gene sets linked to NEU, BMI, and gastrointestinal diseases (Table S37). Despite these distinctions, both SA– and SI–trait pairs exhibited enrichment in gene sets involved in glycine, serine, and threonine metabolism (SI–ADHD, SA–DSN, and SA–NEU) and systemic lupus erythematosus (SA/SI–BD, SA/SI–NEU, SI–RSKT, and SA/SI–SCZ). Further enrichments were more broadly observed in pathways associated with DNA packaging, DNA/nucleosome binding, and chromatin remodelling (Table S37).

MAGMA gene-property analysis was used to examine tissue-specific expression profiles of genes mapped through conjFDR analyses. Significant enrichment was observed for SA–NEU-associated genes in tibial nerve tissue, as well as in brain tissue at 26 weeks post-conception. Significant enrichment was also found for SA–DSN associated genes at 2 years of age, whilst no tissue enrichment was found for SI-pairs. Full results, including non-significant findings, can be found in Table S38.

## 4. Discussion

### 4.1. Main findings

This study provides a detailed characterisation of the shared genetic architecture between SI, SA, and a broad set of psychiatric, behavioural, and somatic phenotypes by integrating two complementary analytic frameworks: LAVA and conjFDR. This joint approach offers greater biological resolution than either method alone. Whereas LAVA identifies the genomic regions where polygenic covariance arises, conjFDR detects the specific variants driving cross-trait enrichment.

Together, these region- and SNP-level analyses yield a more granular understanding of the pleiotropy underlying vulnerability to suicidal thoughts and behaviours.

Across traits, SA and SI differed substantially in both the magnitude and distribution of jointly associated loci, reinforcing accumulating evidence that they reflect partially distinct etiological processes rather than points along a simple severity continuum. SA demonstrated greater genetic sharing with traits characterised by behavioural disinhibition, substance use, psychosis liability, and metabolic dysregulation, whereas SI mapped more consistently onto internalizing features, cognitive–affective traits (e.g., neuroticism), and sleep- and pain-related phenotypes. These diverging patterns are consistent with epidemiological findings showing the limited predictive value of SI for subsequent SA and suggest that these phenotypes may arise from separable liability pathways with only partial overlap (Klonsky et al., 2021b). This is relevant for risk-stratification frameworks that historically treat SI and SA as gradations of a common process. It is noteworthy that neurodevelopmental disorders such as ADHD and ASD were associated with both SA and SI, and that this pattern was consistent across LAVA and conjFDR analyses. Moreover, the patterns observed for SA and SI were highly similar when considering ADHD and MSCP, reflecting the strong genetic correlation specifically between ADHD and pain traits (Ciochetti et al., 2023).

A small number of loci emerged across both methods despite stringent conditioning on MDD and PTSD in the LAVA analysis. These convergent signals—one for SA–ADHD and one for SA–SCZ—likely represent higher-confidence pleiotropic regions whose effects are robust across statistical frameworks and not simply reflections of global correlation or collinearity with common psychiatric comorbidities. Genes mapped to these SNPs and regions participate in neuronal and immune cell proliferation, differentiation, and survival. Their cross-trait presence suggests that immune and early neurodevelopmental processes may constitute core mechanisms linking suicidality with external phenotypes.

Enrichment analyses further demonstrated that the biological pathways shared with SA and SI extend beyond classical neurobiological processes. Many phenotype pairs showed enrichment for gene sets related to brain morphology, cortical and subcortical structure, chromatin dynamics, transcriptional regulation, and glycine–serine–threonine metabolism—implicating neurotransmission, neuronal growth, and excitatory–inhibitory balance. Notably, glycine transporter genes (e.g., *SLC6A9*), repeatedly implicated in SA- and SI-based pairs, modulate NMDA receptor tone and astrocytic regulation of glutamatergic signaling (Cummings and Popescu, 2015; Mizzi and Blundell, 2025). This aligns with growing evidence that NMDA modulation can acutely reduce suicidal thoughts and supports the hypothesis that glycine–NMDA pathways may be mechanistically relevant to suicidal vulnerability (Canuso et al., 2021; Fu et al., 2023; Pompili, 2020). An important and often overlooked finding of this study is the reproducible tissue enrichment in non-brain tissues, including liver, intestine, colon, skin, vasculature, and kidney cortex. Rather than representing peripheral noise, these results align with a substantial body of literature documenting bidirectional interactions between psychiatric phenotypes and systemic processes such as inflammation, metabolic regulation, epithelial integrity, microbiome–immune crosstalk, and vascular function. The presence of suicide-associated pleiotropic genes in these tissues suggests that suicidality may not be exclusively neurocentric but may arise from integrated brain–body mechanisms. This perspective is consistent with epidemiological links between suicidality and autoimmune disorders (Brundin et al., 2015; Isung et al., 2020; Yang et al., 2025), cardiometabolic disease (Chen et al., 2023), and gastrointestinal dysregulation (Costanza et al., 2025), pointing to multisystemic biological signatures worthy of further investigation.

Across multiple trait combinations, we also identified shared loci enriched for gene sets associated with neuroticism, behavioural control, and age at first sexual intercourse (AFSI). Previous Mendelian randomisation studies indicate that AFSI may index underlying neurobiological vulnerability to self-harm, potentially via mediating effects of MDD and SCZ polygenic risk (Dong et al., 2024). This supports the hypothesis that early-life behaviours and developmental trajectories may reflect or contribute to latent liabilities shared with suicidal phenotypes.

Taken together, the patterns observed here—methodological convergence at a small number of high-confidence loci; trait-specific and phenotype-specific enrichment signatures; and the presence of both neural and peripheral tissue signals—support a model in which suicidal thoughts and behaviours arise from a heterogeneous architecture composed of partly independent but interacting neurodevelopmental, neurochemical, behavioural, and somatic pathways. These findings reinforce the need for phenotypic precision when studying suicidal outcomes and indicate the usefulness of integrating multilayered genomic information to uncover biologically meaningful pleiotropy.

### 4.2 Strengths and limitations

A key strength of this study is the integration of two complementary analytic frameworks that capture different dimensions of pleiotropy. LAVA provides spatially localised estimates of covariance, whereas conjFDR detects cross-trait enrichment even when single-trait GWAS lack genome-wide significance. Used together, these methods enable both validation and refinement of shared genetic signals. Systematic conditioning on MDD and PTSD in the LAVA analysis further improves interpretability by distinguishing trait-specific from comorbidity-driven effects. Additional strengths include the use of large, up-to-date GWAS datasets and the inclusion of a broad range of psychiatric and non-psychiatric traits, which permits the exploration of shared liability beyond diagnostic boundaries.

However, several limitations warrant consideration. Some phenotypes (e.g., ASD) remain underpowered, which may lead to underestimation of pleiotropy. All analyses rely on measures of genetic correlation and joint association, which do not imply causality. Functional validation will require integration with transcriptomic, proteomic, or cellular models. The predominance of European-ancestry samples limits generalisability and underscores the need for cross-ancestry genomic resources. Finally, enrichment findings in peripheral tissues should be interpreted cautiously and validated in multi-omic datasets, given the complexity of tissue specificity and gene regulation.

### 4.3 Conclusions

This study refines current understanding of the genetic architecture linking SA and SI to a broad spectrum of psychiatric, somatic, and behavioural traits. By integrating region-level genetic covariance with SNP-level cross-trait association mapping, we enhanced the resolution for detecting pleiotropic loci and implicated pathways—particularly those related to neurodevelopment, immune dysregulation, and glycine–NMDA signaling. The distinct and partially overlapping association profiles observed for SA and SI underscore the importance of precise and differentiated phenotypic definitions when interrogating suicidal thoughts and behaviours.

Collectively, these findings may inform future efforts in stratified risk prediction and guide the development of mechanistically grounded interventions.

## Appendix

* ECNP Network on Suicide Research and Prevention [Collaborative Author Group] – Members of the European College of Neuropsychopharmacology (ECNP) Network on Suicide Research and Prevention are: Alan Apter, Schneider, Children’s Medical Center of Israel & Tel Aviv University, Israel; Enrique Baca-García, Hospital Universitario Fundación Jiménez Díaz & Universidad Autónoma de Madrid, Spain; Judit Balázs, Institute of Psychology, Eötvös Loránd University, Hungary; Shira Barzilay, University of Haifa, Israel; Julio Bobes, University of Oviedo, Spain; Raffaella Calati, Department of Psychology, University of Milan-Bicocca, Italy & Department of Adult Psychiatry, Nimes University Hospital, France; Vladimir Carli, Karolinska Institutet, Sweden; Philippe Courtet, Université de Montpellier & CHU Montpellier, France; Giuseppe Fanelli, University of Bologna, Italy & Radboud University Medical Center, The Netherlands; Xenia Gonda, Semmelweis University, Hungary; Iria Grande, Bipolar and Depressive Disorders Unit, Hospital Clinic de Barcelona, Departament de Medicina Universitat de Barcelona, Institut d’Investigacions Biomèdiques August Pi i Sunyer (IDIBAPS), Centro de Investigación Biomédica en Red de Salud Mental (CIBERSAM), Spain., Spain; Vasileios Masdrakis, National and Kapodistrian University of Athens, Greece; Maurizio Pompili, Sapienza University of Rome, Italy; Zoltán Rihmer, Semmelweis University, Hungary; Dan Rujescu, Medical University of Vienna, Austria; Pilar A. Saiz, University of Oviedo & CIBERSAM, Spain; Anjali Sankar, Copenhagen University Hospital Rigshospitalet, Denmark; Marco Sarchiapone, University of Molise, Italy; Alessandro Serretti, “Kore” University of Enna & Oasi Research Institute-IRCCS, Troina, Italy; Marcus Sokolowski, Karolinska Institutet, Sweden; Robertas Strumila, CHU Montpellier, France; Lars-Håkan Thorell, Linköping University, Sweden; Danuta Wasserman, Karolinska Institutet, Sweden; Jerzy Wasserman, Karolinska Institutet, Sweden; Abraham Weizman, Tel Aviv University & Geha Hospital, Israel; Gil Zalsman, Tel Aviv University & Geha Hospital, Israel.

## 8. Author Disclosure

### 8.2. Contributors

Diego L. Rovaris and Giuseppe Fanelli conceptualised, supervised and coordinated the study. Isabella Folego-Temoteo and Alfonso Martone performed the statistical analysis, curated the data and wrote the initial draft of the manuscript. Alessandro Serretti, Chiara Fabbri, Eugenio H. Grevet, Claiton H.D Bau, Cibele E. Bandeira, Gabriel R. Fries, Janita Bralten, Anna R. Docherty, and Allison E. Ashley-Koch, Iria Grande and Xenia Gonda reviewed the manuscript and provided critical insights. Giuseppe Fanelli, Diego L. Rovaris, Alfonso Martone and Isabella Folego-Temoteo finalised the manuscript. All authors contributed to and have approved the final manuscript.

## Data Availability

All data produced in the present study are available upon reasonable request to the authors.

https://docs.google.com/spreadsheets/d/1nleghR8D4q81jVQ7t4akSDbGkiAuMpFWcoiaRyTLyJk/edit?gid=0#gid=0

https://docs.google.com/document/d/1LFUIUvjLs6eJLHf8Ht24QFbaWbjxdG0P8WYEvQwRINg/edit?usp=drive_link

