## Supplementary Figures for "Dissecting the pleiotropic genetic architecture of suicide attempt, suicidal ideation, and thirteen correlated traits"

#### Supplementary Figure 1. Global genetic correlations

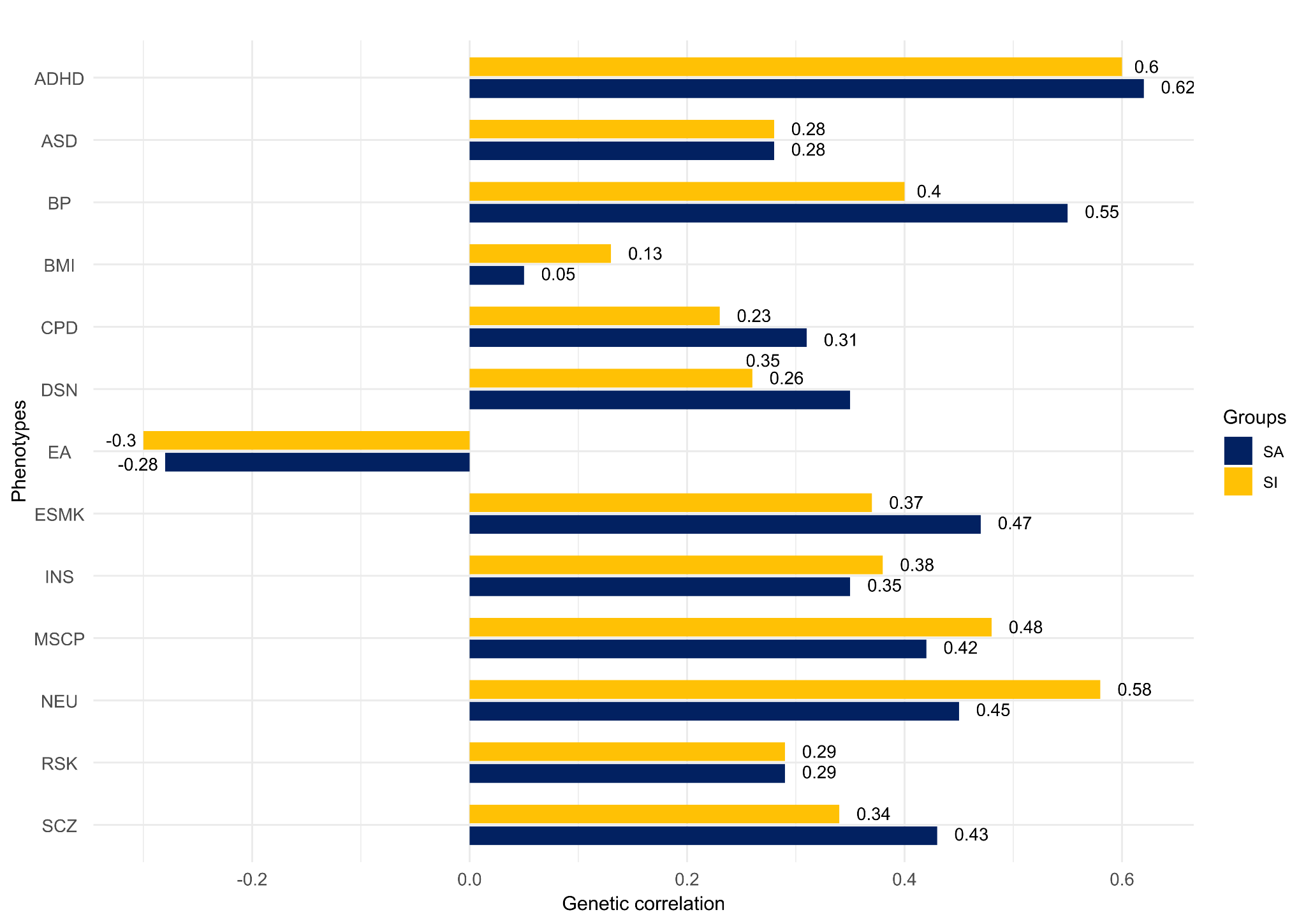

ASD, Autistic Spectrum Disorder; BD, Bipolar Disorder; CPD, Cigarettes per Day; DSN, Disinhibition; EA, Educational Attainment; ESMK, Ever Smoker; INS, Insomnia; MSCP, Multi-Site Chronic Pain; NEU, Neuroticism; RSKT, Risk Tolerance; SA, Suicide Attempt; SCZ, Schizophrenia; SI, Suicidal Ideation.

### Supplementary Figures 2-5.

A. Distribution of differentially expressed genes across 30 general tissue types, as defined by FUMA based on GTEx version 8. Highlighted tissues represent those in which genes mapped to shared locus between the analyzed phenotypes showed significantly elevated expression (adjusted p < 0.05).

B. Distribution of differentially expressed genes across 54 specific tissue types, including detailed subdivisions such as individual brain regions. This analysis helps identify tissues potentially relevant to the biological mechanisms underlying the observed genetic associations.

#### Supplementary Figure 2. SAxEA - GTEx v8 - 30 general tissue types and 54 tissue types

A.

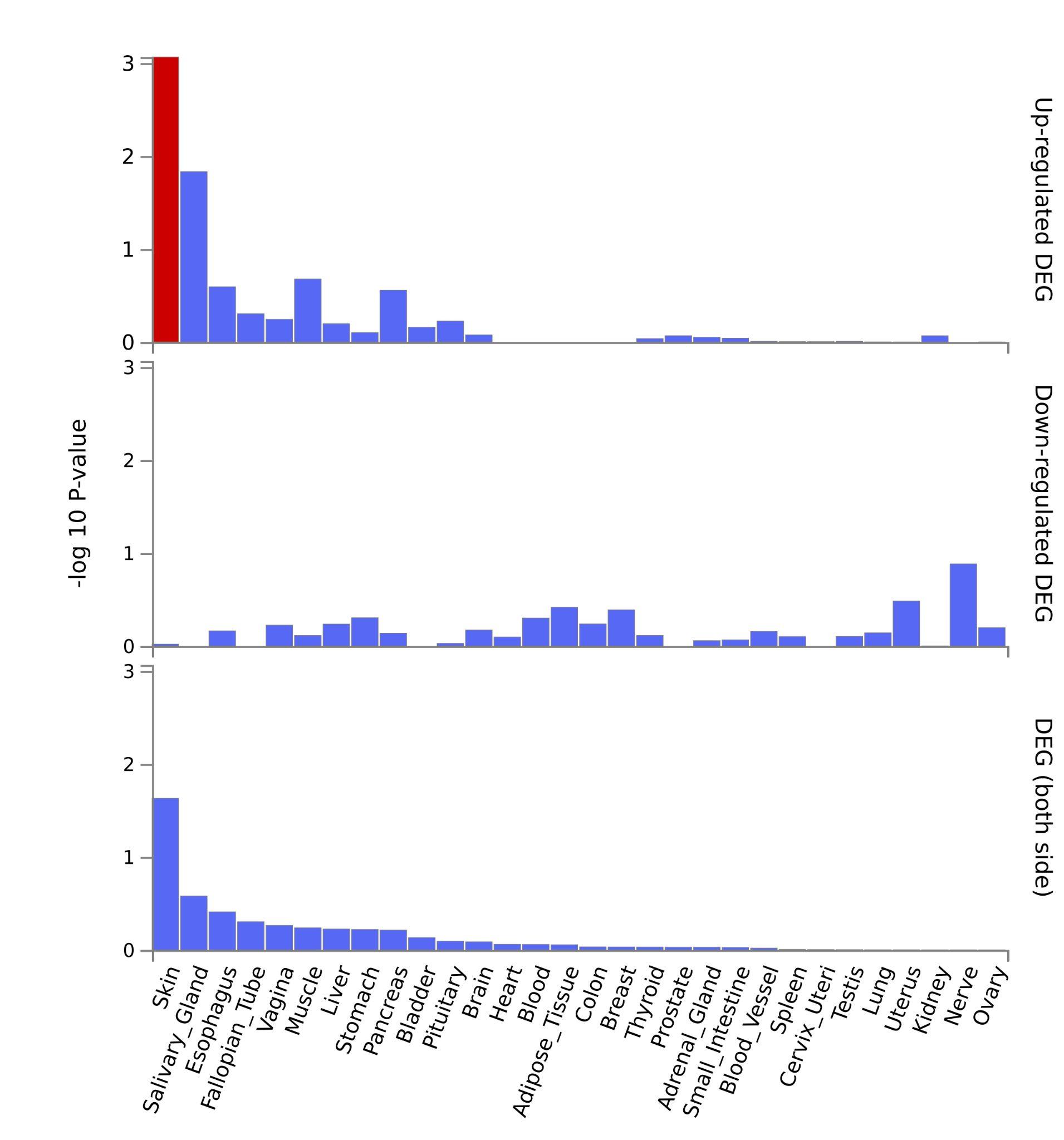

B.

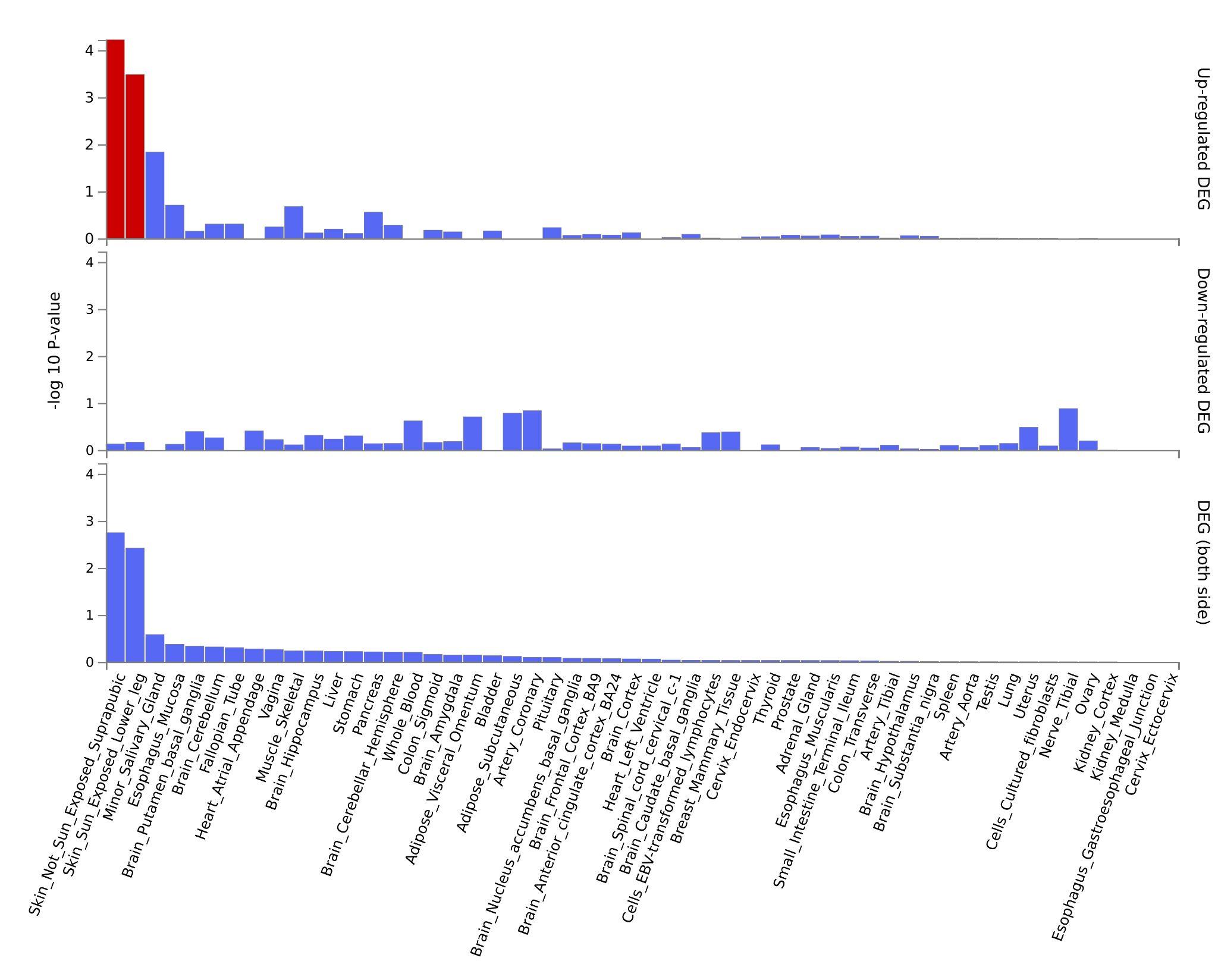

#### Supplementary Figure 3. SAxESMK - GTEx v8 - 30 general tissue types

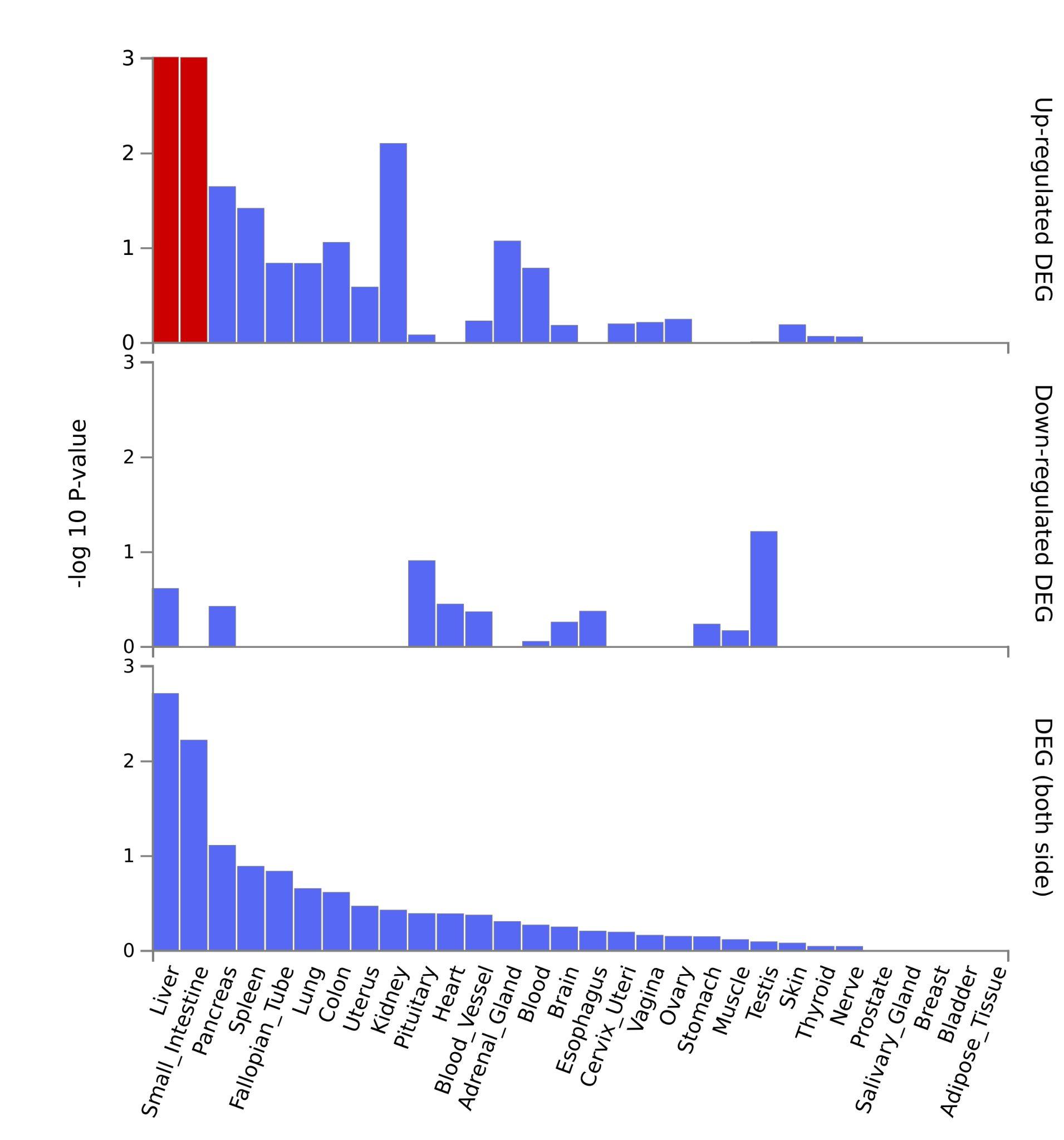

#### Supplementary Figure 4. SIxADHD - GTEx v8 - 30 general tissue types and 54 tissue types

A.

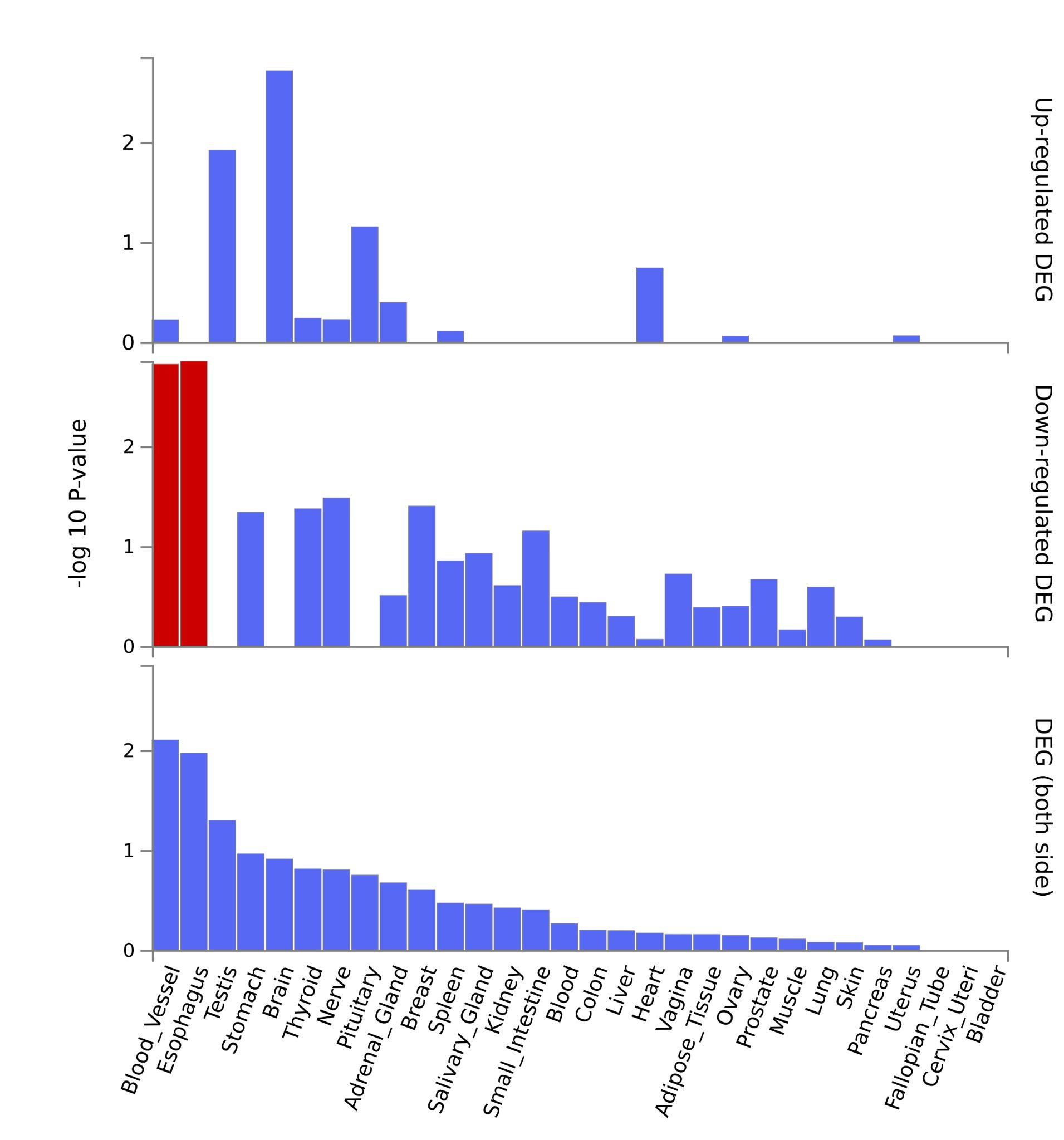

B.

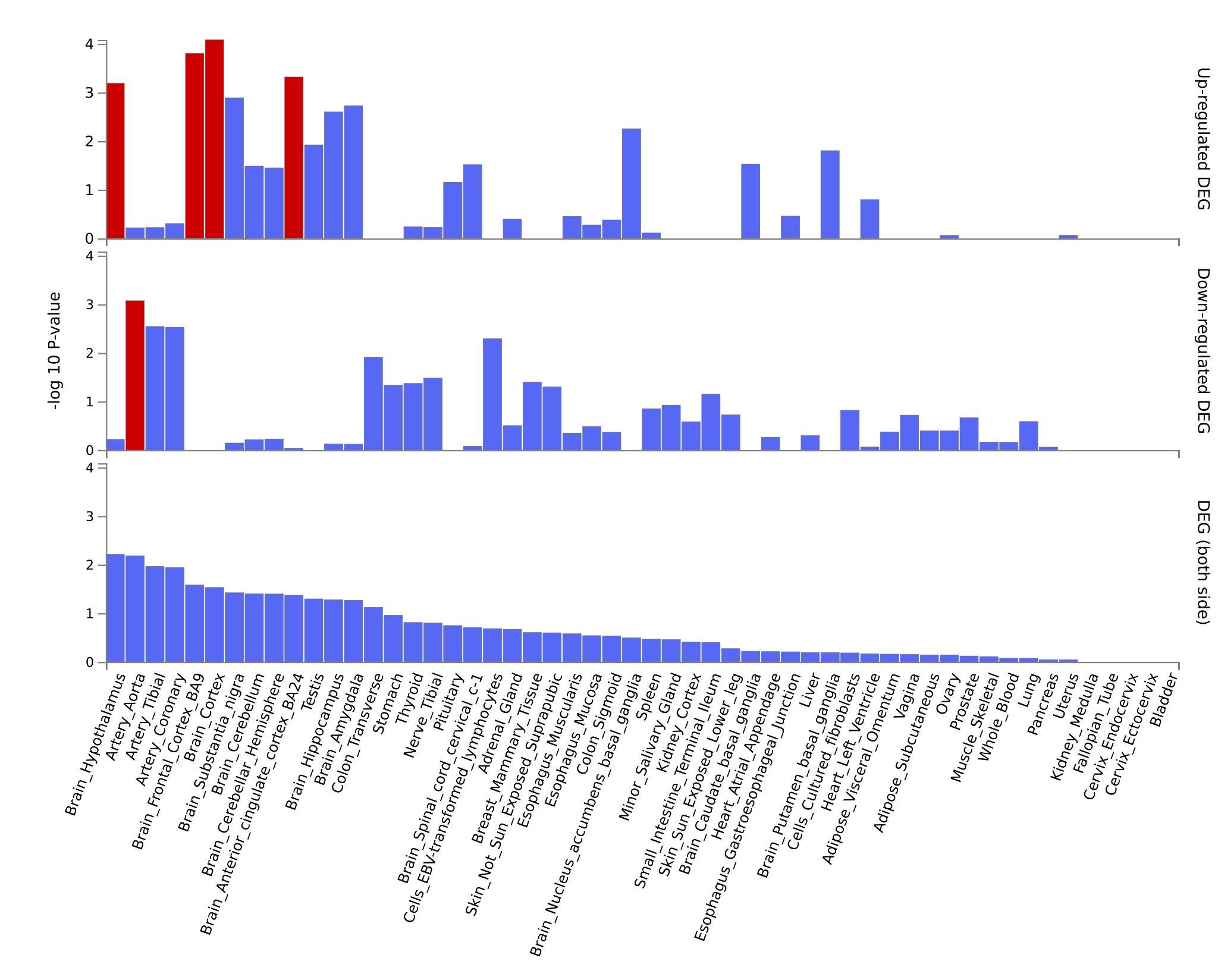

#### Supplementary Figure 5. SIxEA - GTEx v8 - 30 general tissue types and 54 tissue types

A.

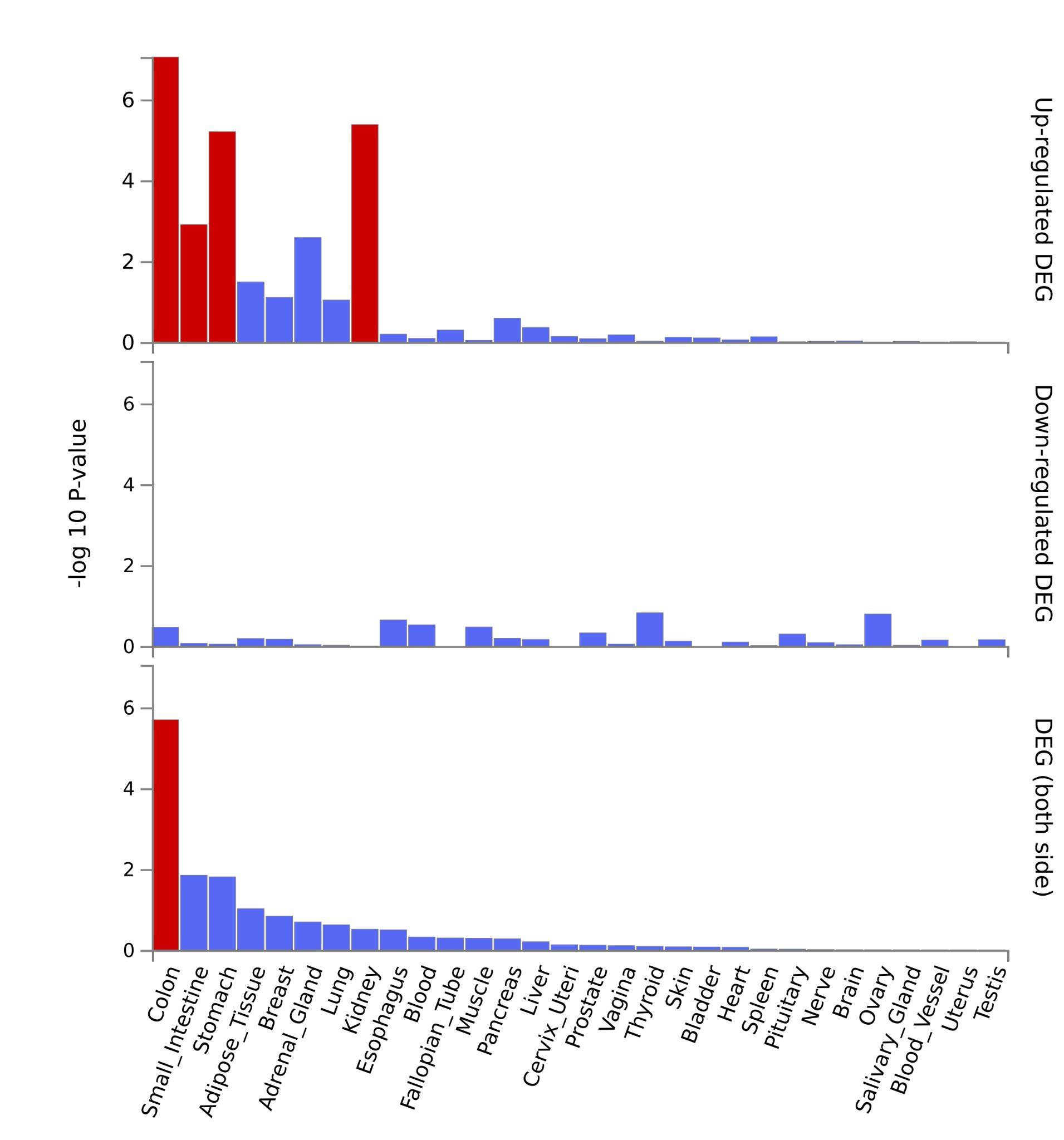

B.

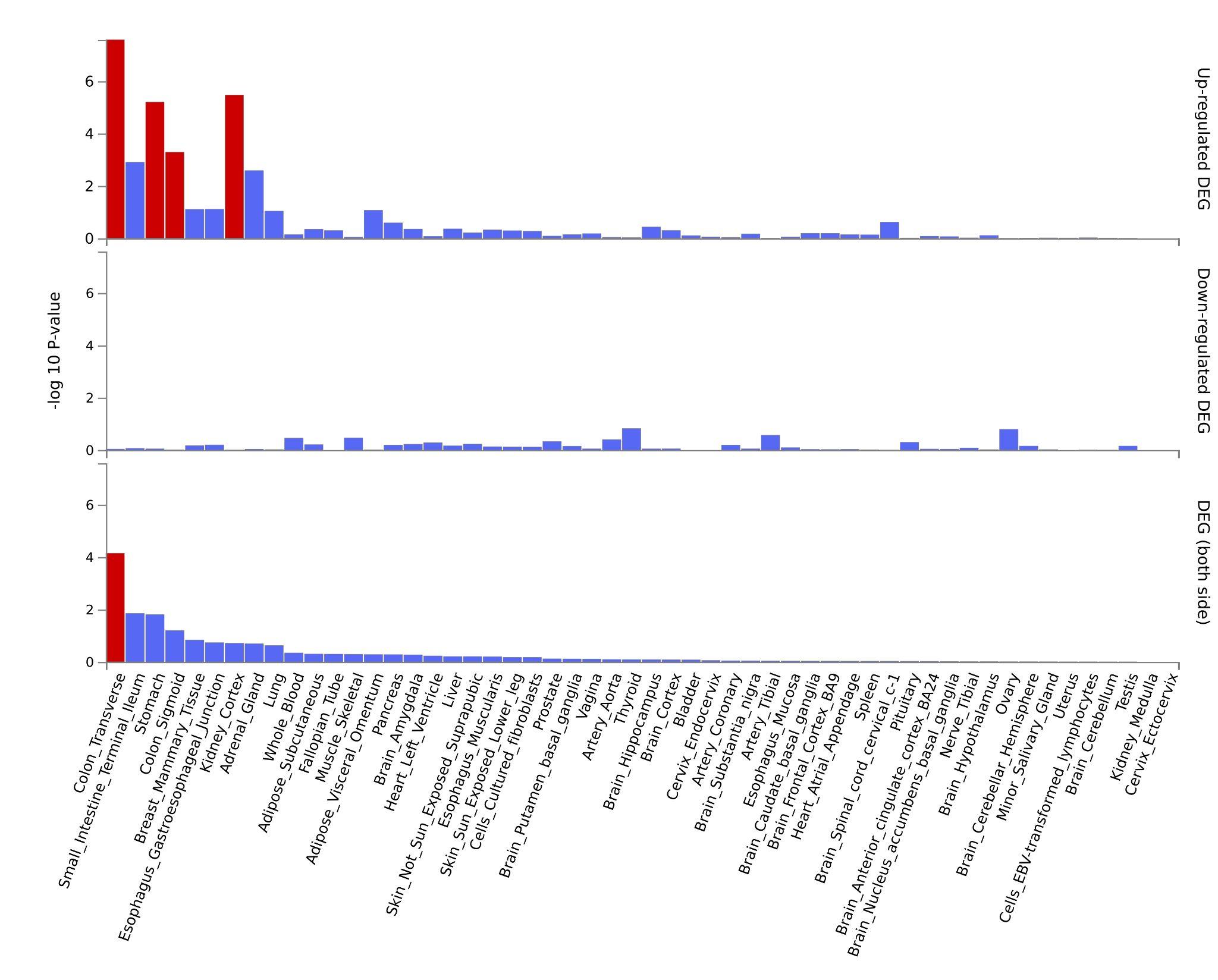

### Supplementary Figures 6-9.

The following figures summarizes the results of gene-set enrichment analyses performed using mapped genes across shared loci between phenotype pairs. Each panel presents a bar plot with three components: The left bar (red) shows the proportion of overlapping genes in each gene set. The middle bar (blue) shows the enrichment significance as the –log10 of the adjusted p-value. The right bar (orange) lists the specific overlapping genes contributing to each enriched term.

**Reactome -** Reactome enrichment is a curated database of biological pathways. This panel reveals enriched pathways involving multiple mapped genes, indicating potential molecular cascades and systems-level processes that may be implicated in the genetic overlap between the traits.

**(GO:BP) - Gene Ontology – Biological Processes:** This panel highlights biological processes significantly enriched among the mapped genes. These processes reflect the broader cellular and physiological functions in which the genes may be involved, such as ion transport, metabolism, and signaling. Enrichment in specific GO:BP terms may suggest biological pathways relevant to the shared genetic basis of the phenotypes analyzed.

**(GO:MF) - Gene Ontology – Molecular Functions:** This analysis identifies the molecular activities performed by gene products, such as binding, catalysis, or transport. Enriched GO:MF terms indicate that the genes mapped from shared loci may converge on specific biochemical roles, which can provide insights into mechanisms of action at the protein level.

**(GO:CC) - Gene Ontology – Cellular Components:** This panel shows which subcellular structures or cellular compartments the gene products are most likely to be localized in (e.g., membrane, synapse, nucleus). Enrichment in GO:CC terms helps to contextualize where in the cell the genetic effects might manifest, contributing to understanding of spatial cellular dynamics.

Not all phenotype pairs or ontology categories are represented in the figures. This absence reflects the fact that no gene sets reached statistical significance for those specific comparisons or categories, and therefore they were not included in the enrichment plots.

#### Supplementary Figure 6. SA-EA - (GO:BP and GO:CC)

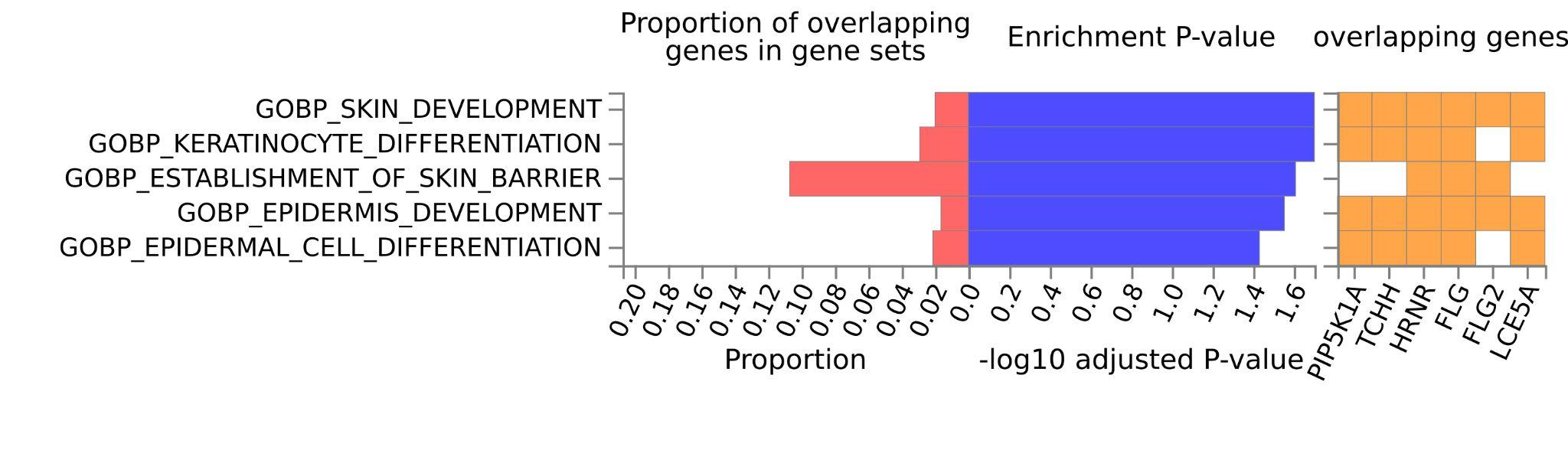

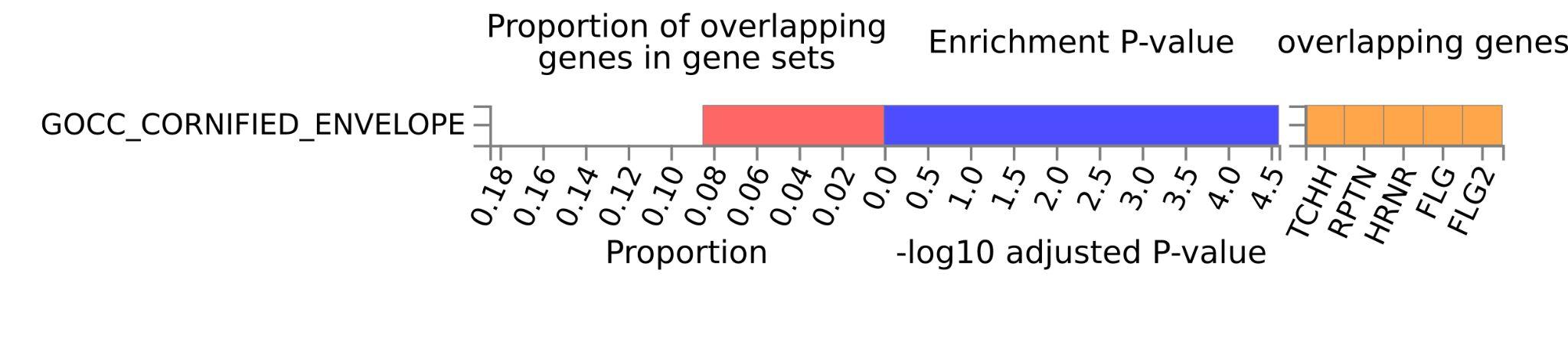

#### Supplementary Figure 7. SA-ESMK - (GO:BP, GO:CC and GO:MF)

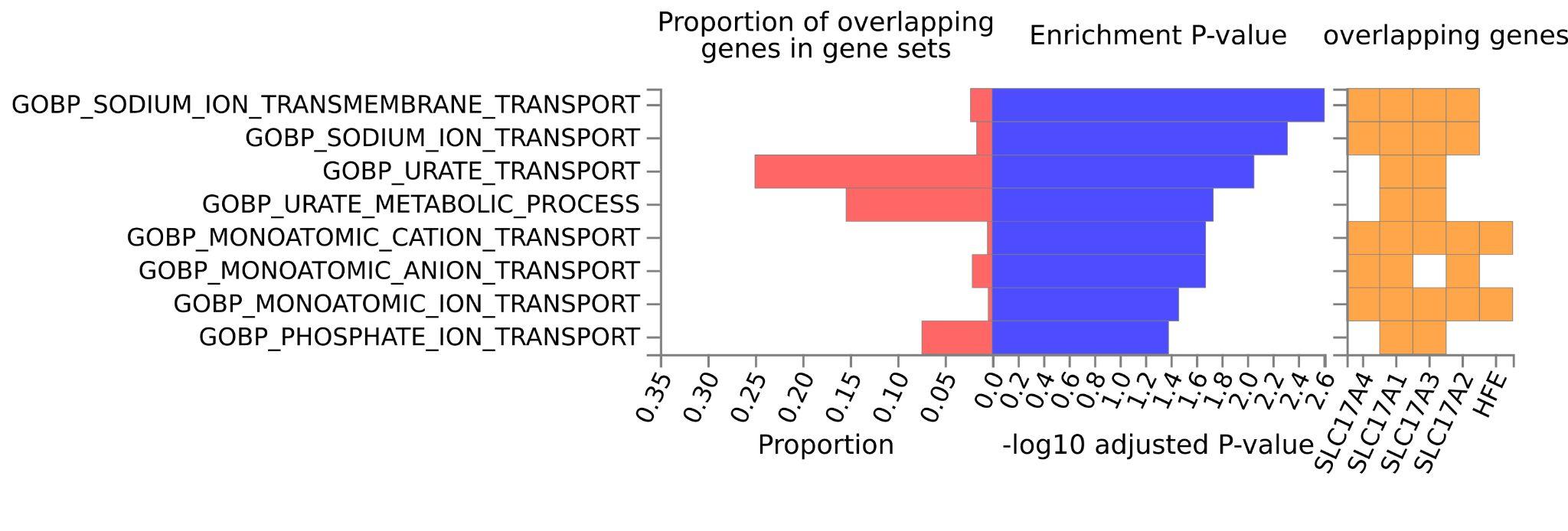

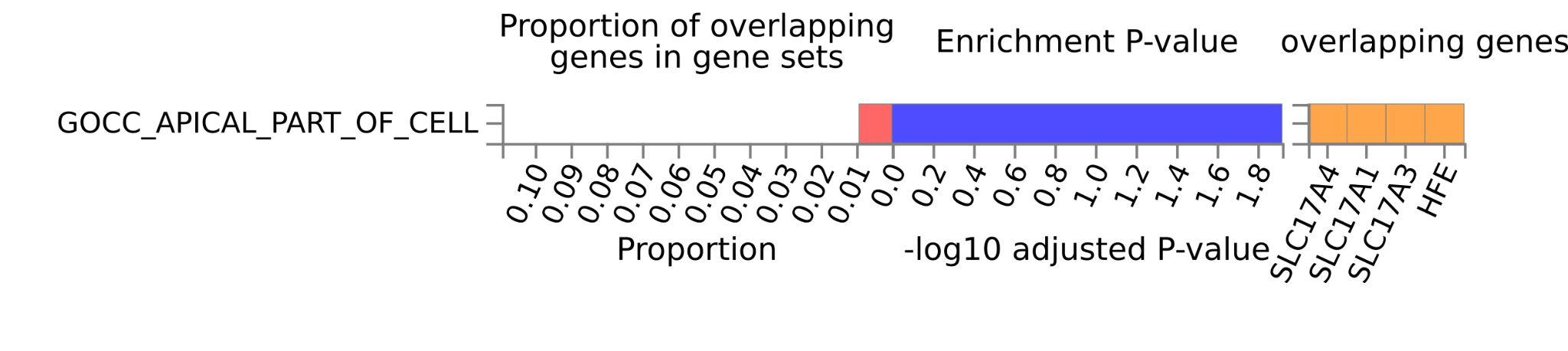

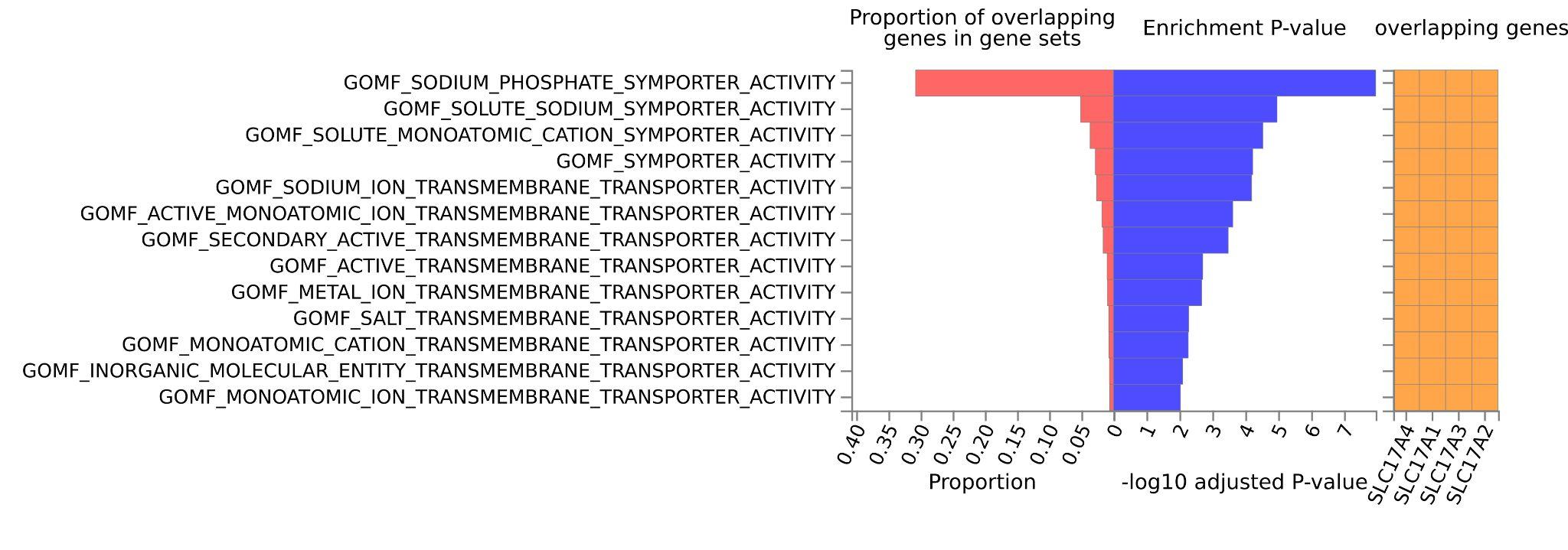

#### Supplementary Figure 8. SI-EA - (GO:BP, GO:CC and GO:MF)

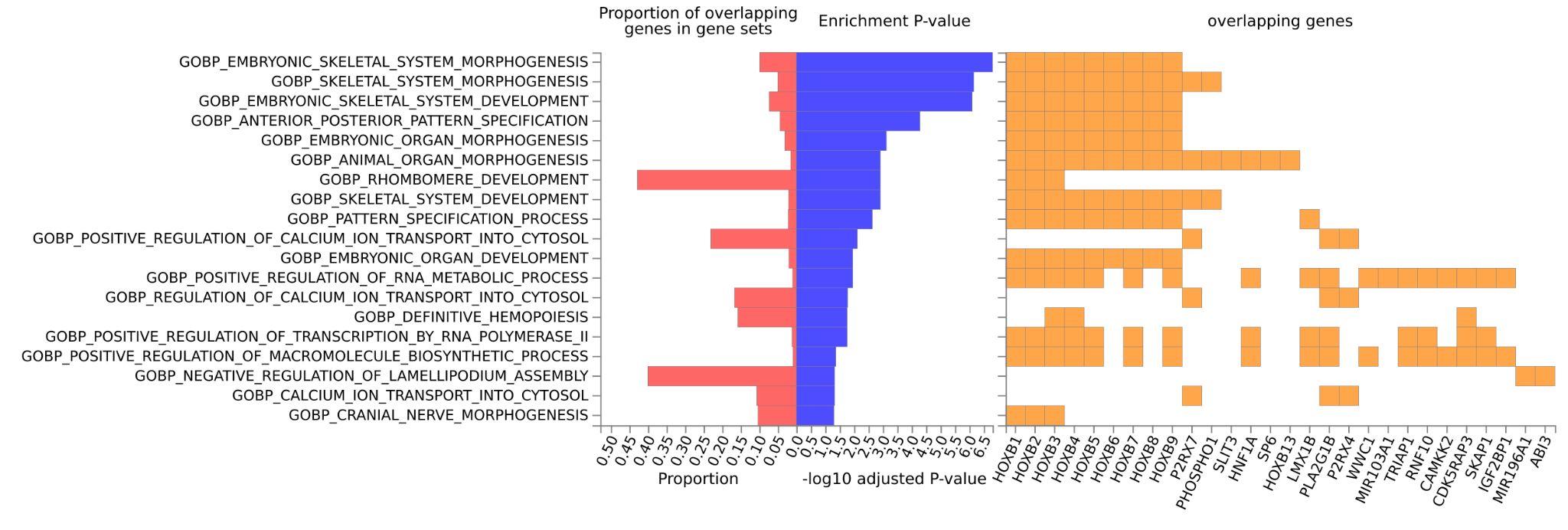

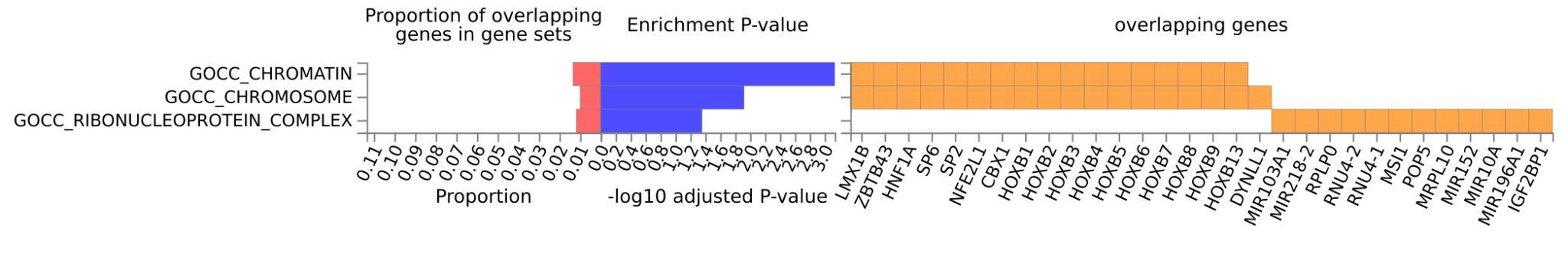

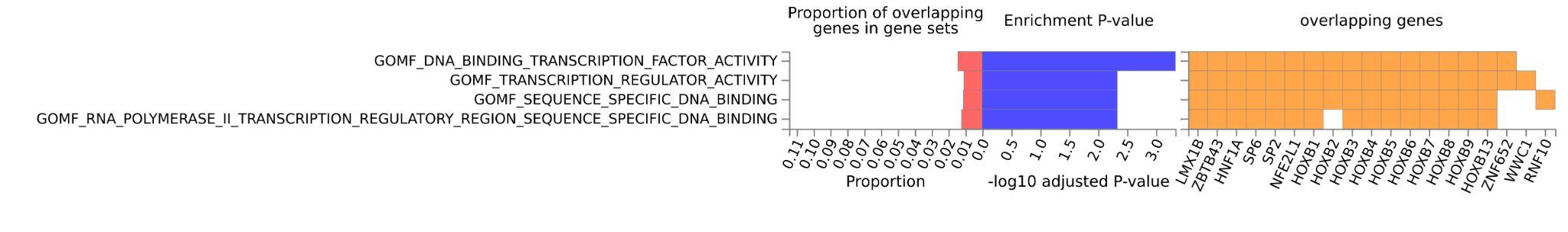

#### Supplementary Figure 9. SI-SCZ - (GO:BP, GO:CC and GO:MF)

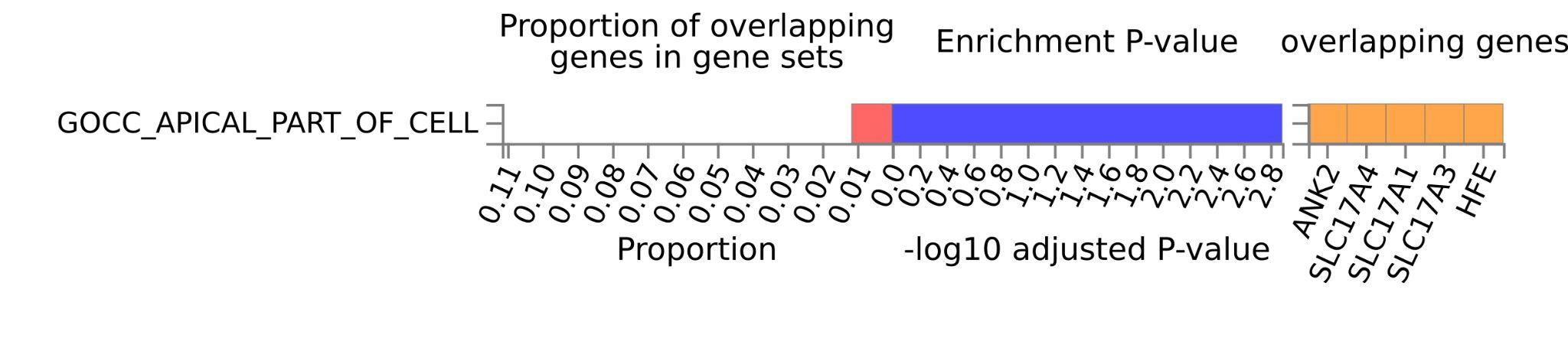

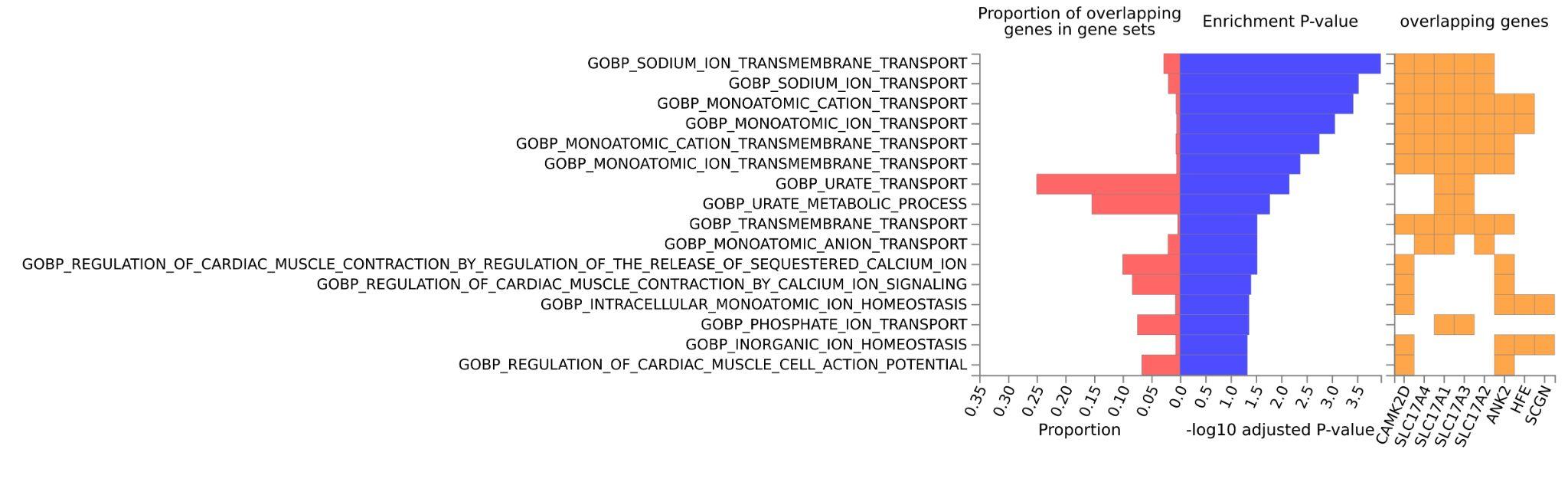

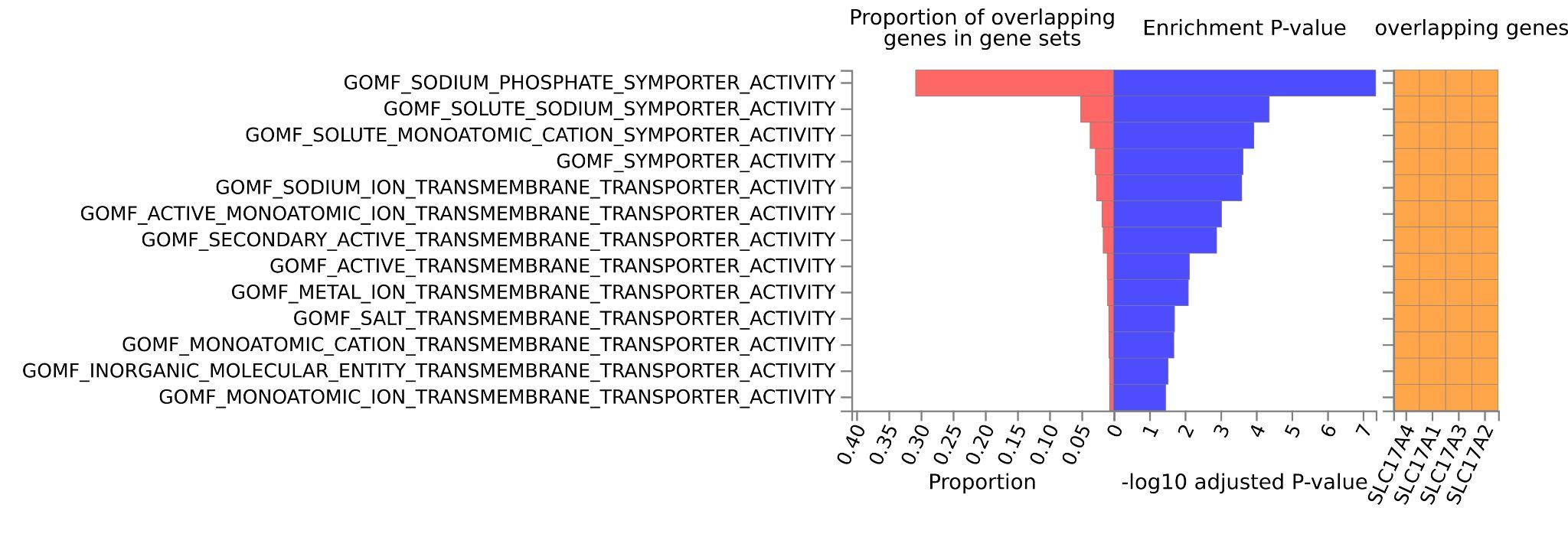

### Supplementary Figures 10-35.

A. Stratified Q-Q plot of nominal versus empirical -log_10_ p-values (corrected for inflation) in the trait of interest below the standard GWAS threshold of p<5x10^-8^ as a function of significance of the association with SA or SI at the level of -log_10_(p)>0, -log_10_(p)>1, -log_10_(p)>2, -log_10_(p)>3 corresponding to p<1, p<0.1, p<0.01, and p<0.001, respectively. Dotted lines indicate the null-hypothesis.

B. Stratified Q-Q plot of nominal versus empirical -log_10_ p-values (corrected for inflation) in SA/SI below the standard GWAS threshold of p<5x10^-8^ as a function of significance of the association with the trait of interest at the level of -log_10_(p)>0, -log_10_(p)>1, -log_10_(p)>2, -log_10_(p)>3 corresponding to p<1, p<0.1, p<0.01, and p<0.001, respectively. Dotted lines indicate the null-hypothesis.

C. Manhattan plots showing the jointly associated SNPs, significant at the standard GWAS threshold of p<5x10^-8^

#### Supplementary Figure 10. SA-ADHD

A.
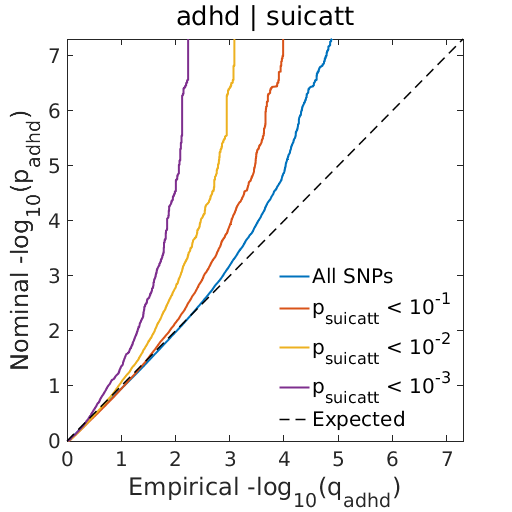
 B.
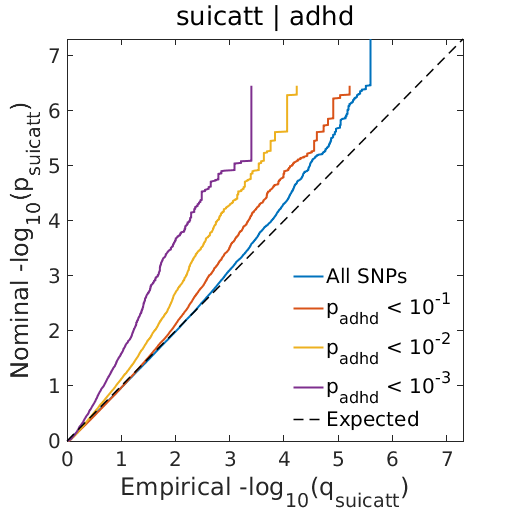

C.
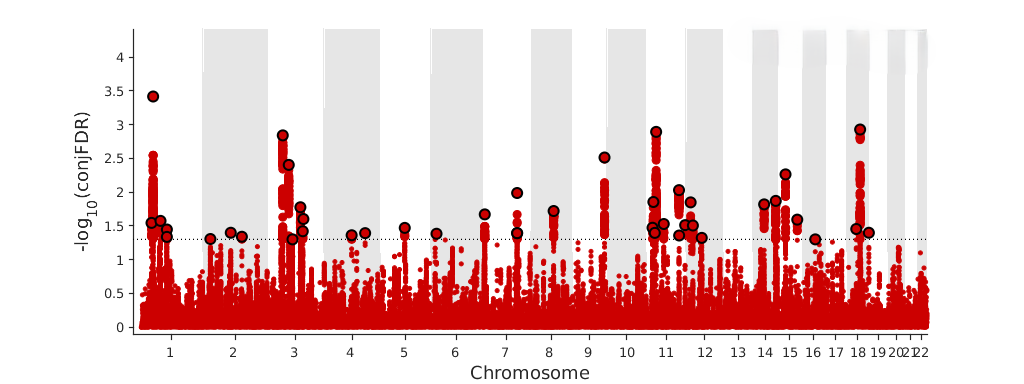

#### Supplementary Figure 11. SA-ASD

A.
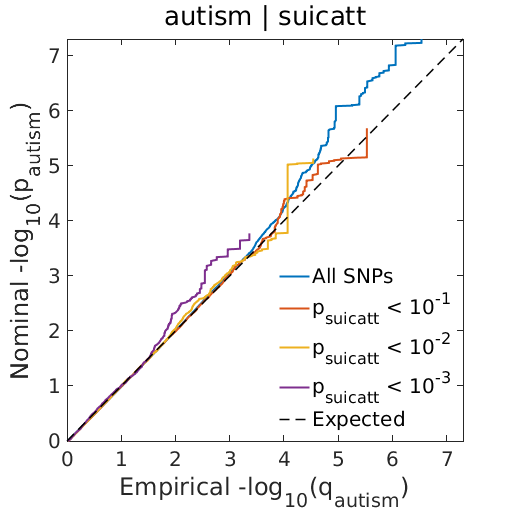
 B.
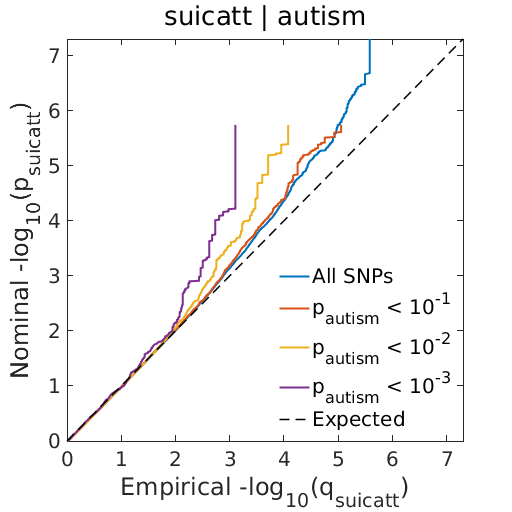

C.
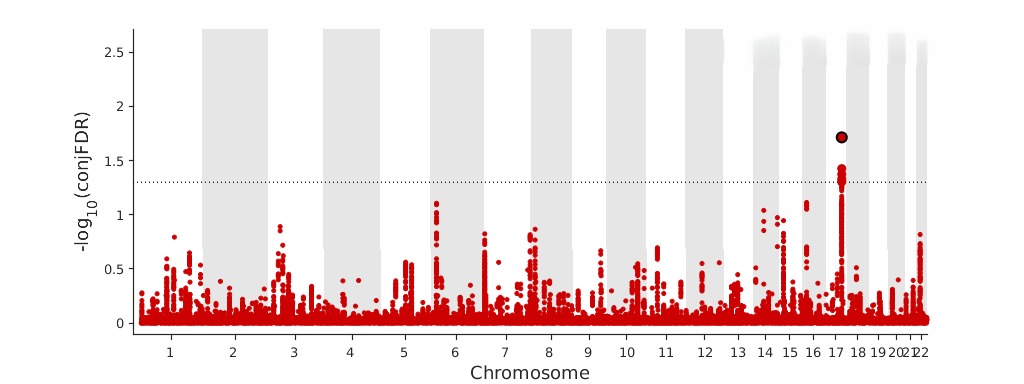

#### Supplementary Figure 12. SA-BMI.

A.
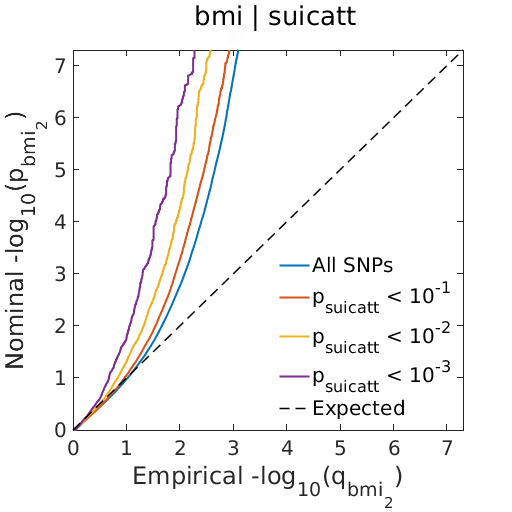
 B.
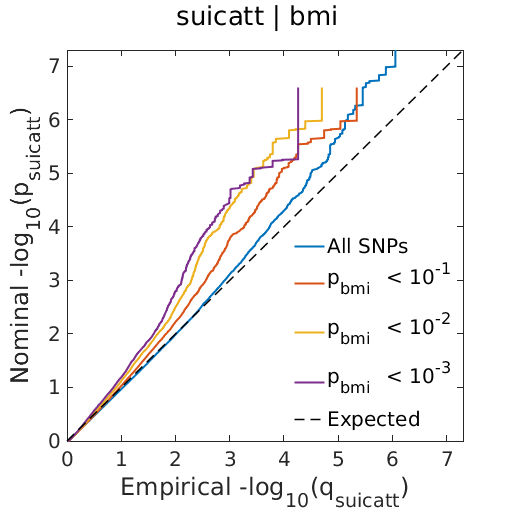

C.
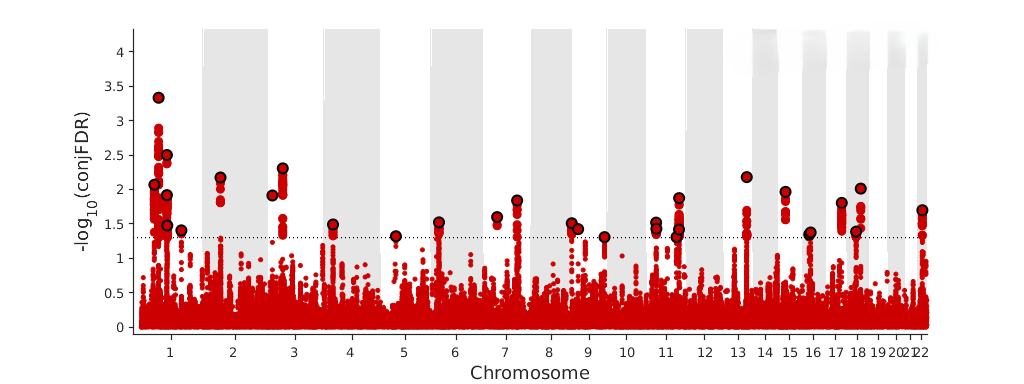

#### Supplementary Figure 13. SA-BP.

A.
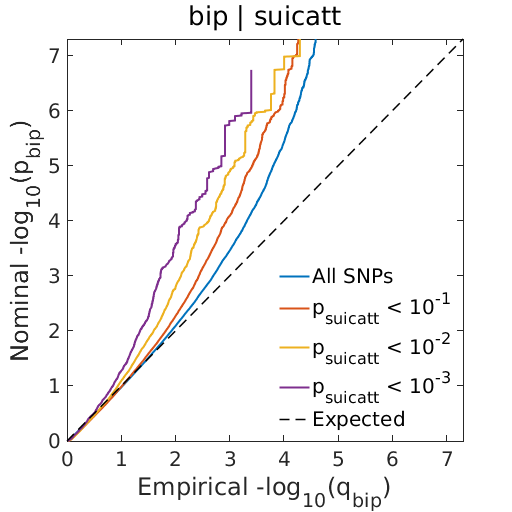
 B.
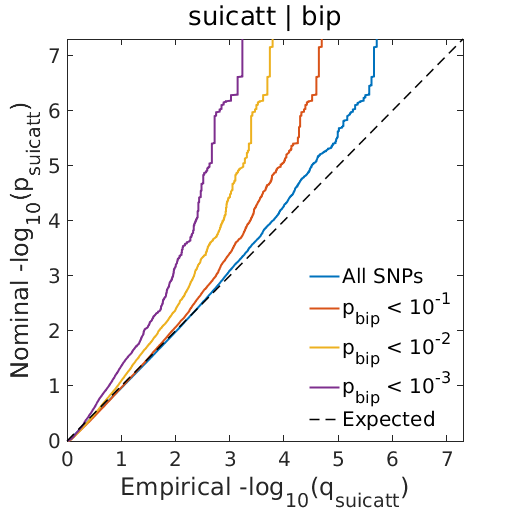

C.
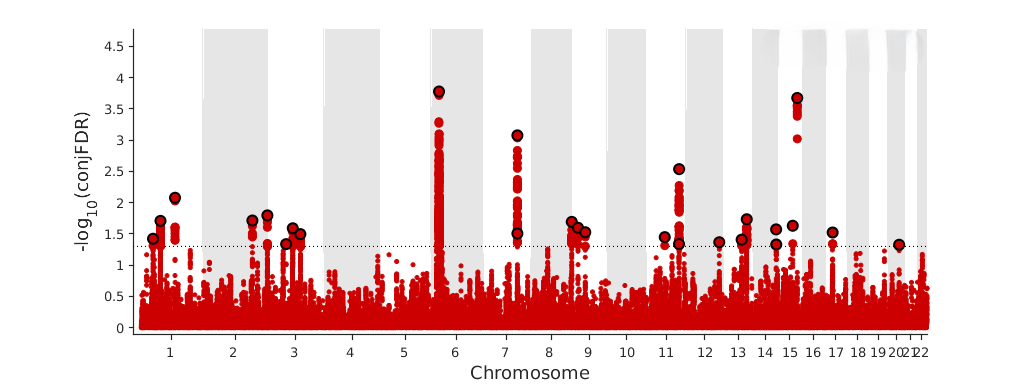

#### Supplementary Figure 14. SA-CPD.

A.
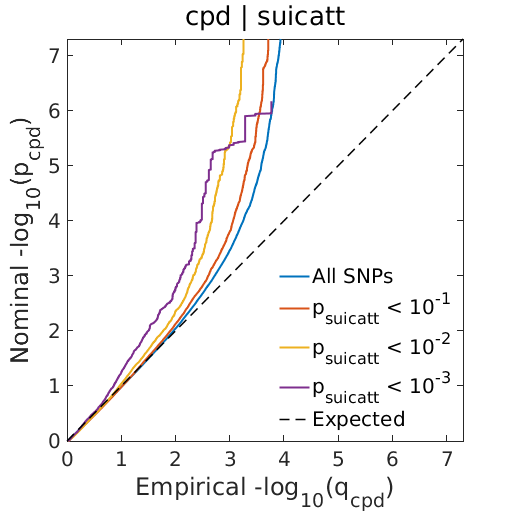
 B.
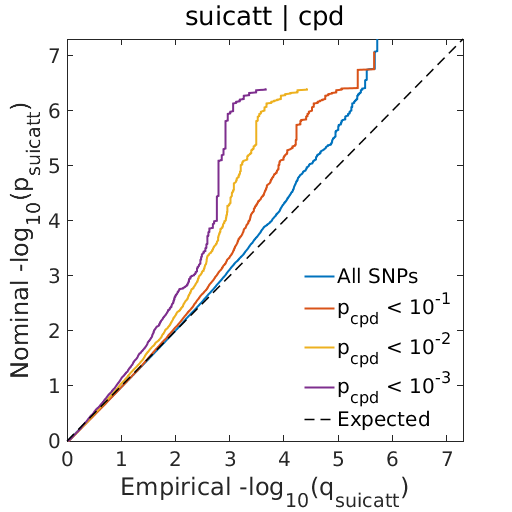

C.
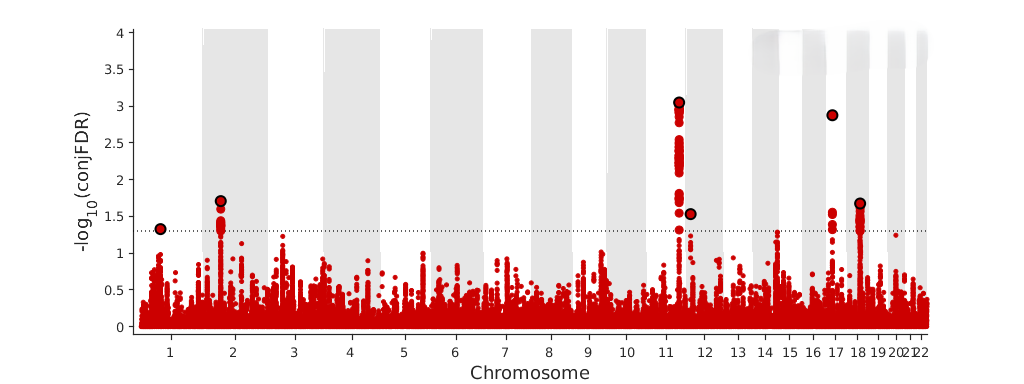

#### Supplementary Figure 15. SA-DSN.

A.
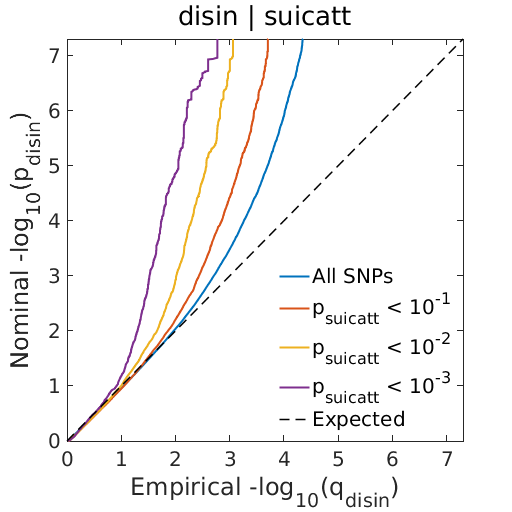
 B.
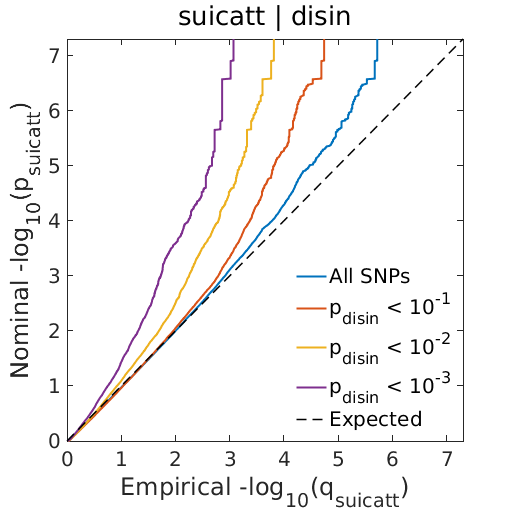

C.
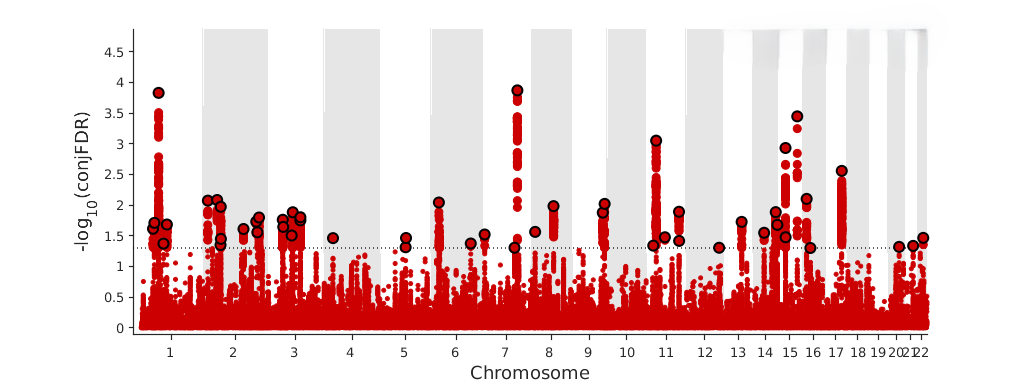

#### Supplementary Figure 16. SA-EA.

A.
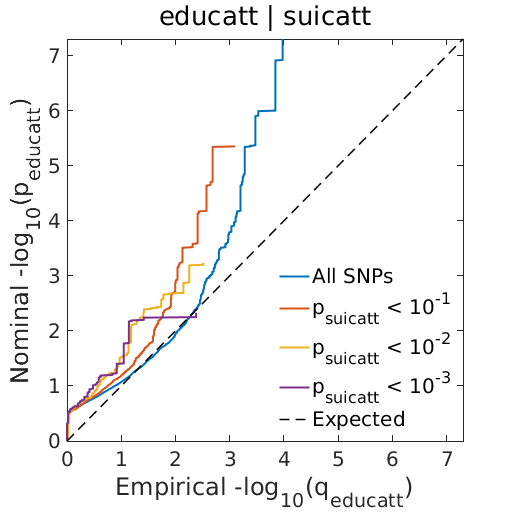
 B.
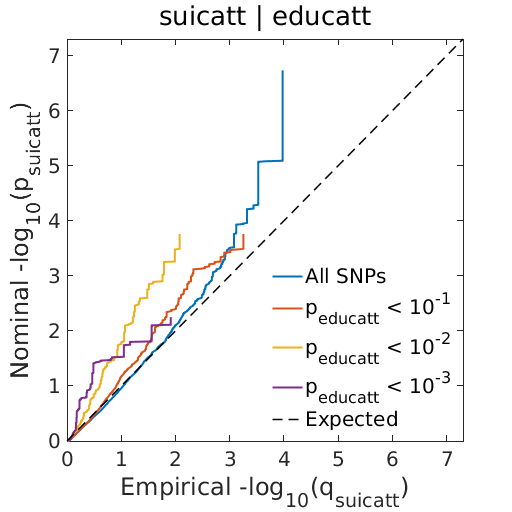

C.
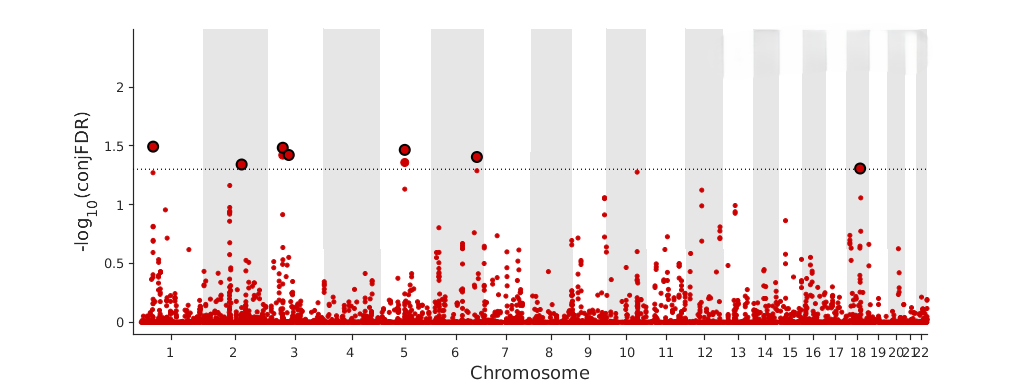

#### Supplementary Figure 17. SA-ESMK.

A.
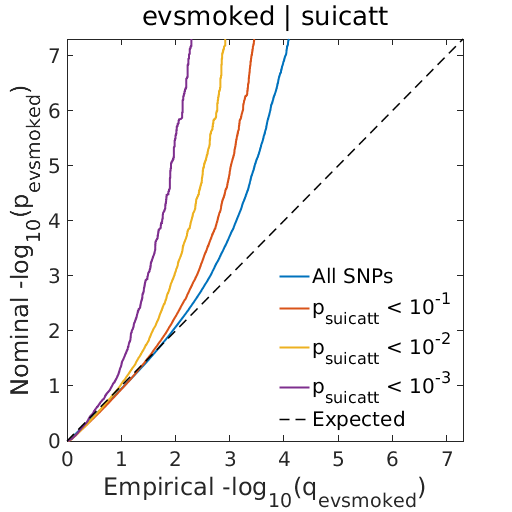
 B.
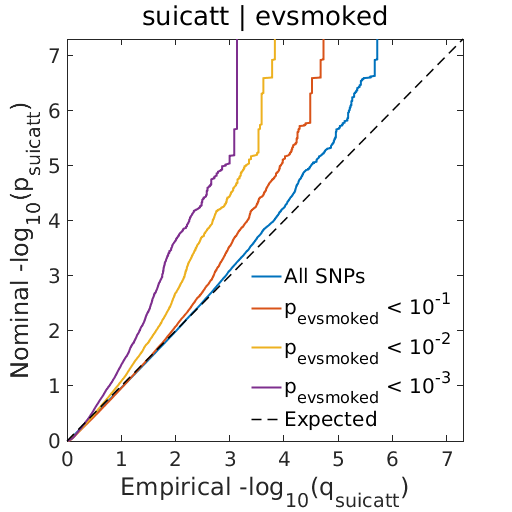

C.
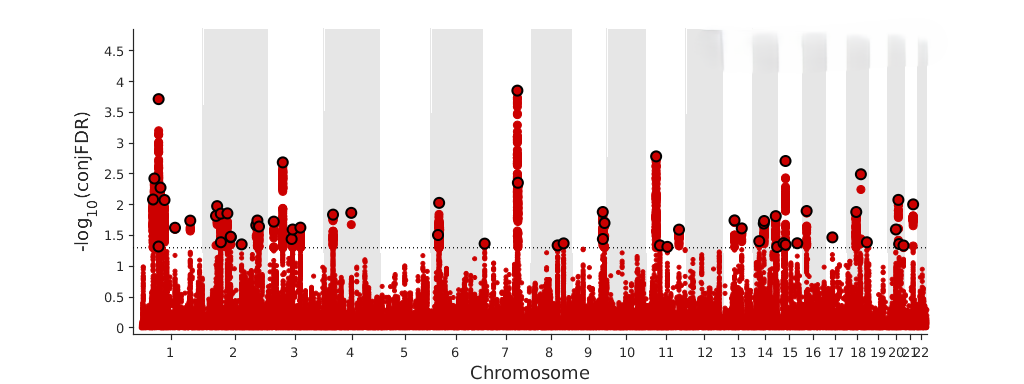

#### Supplementary Figure 18. SA-INS.

A.
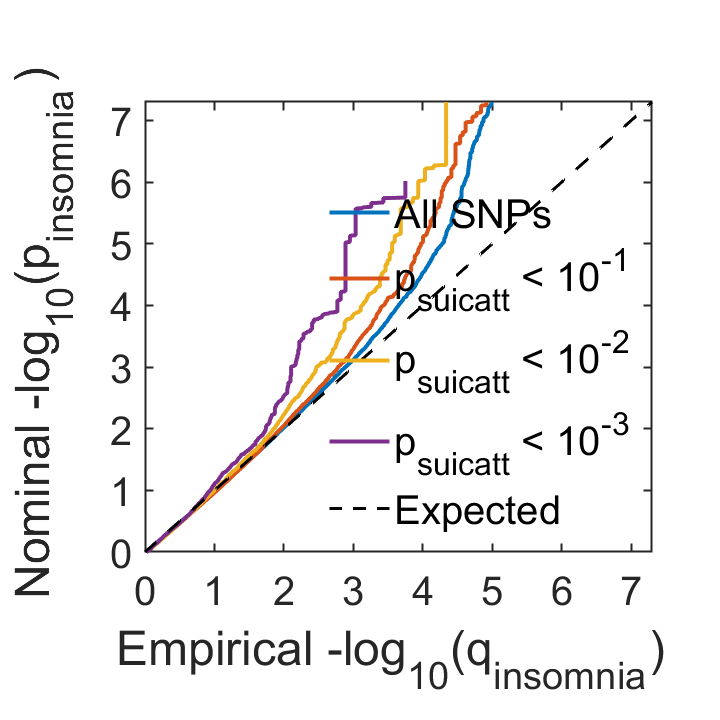
 B.
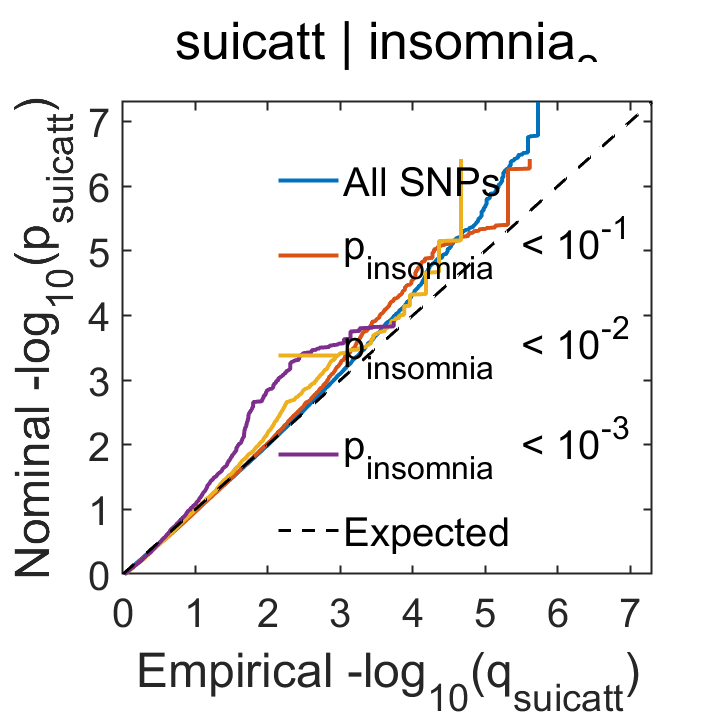

C.
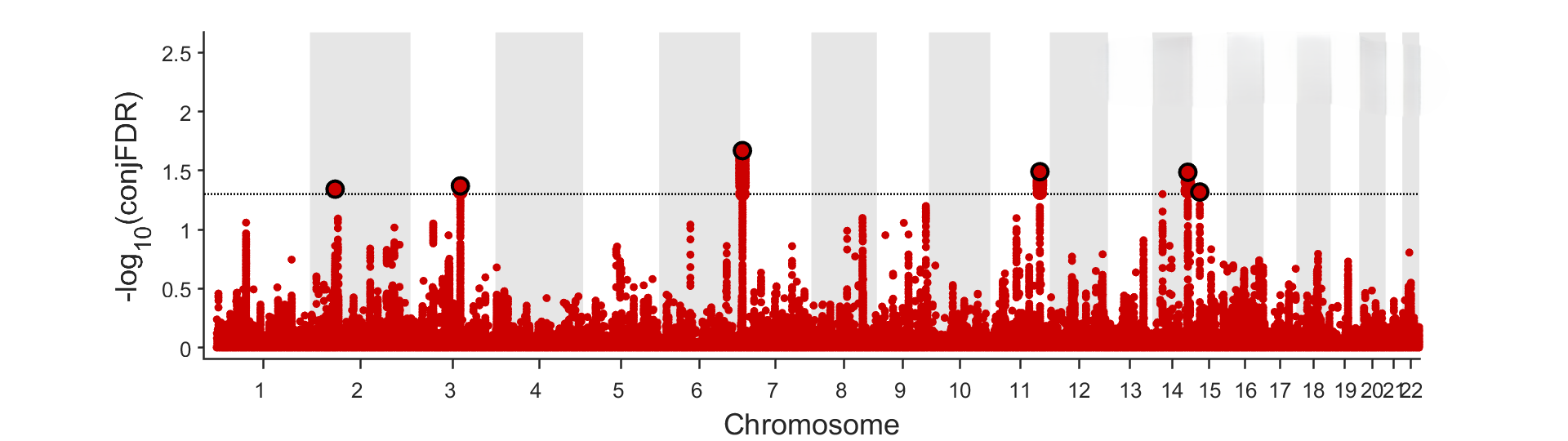

#### Supplementary Figure 19. SA-MSCP.

A.
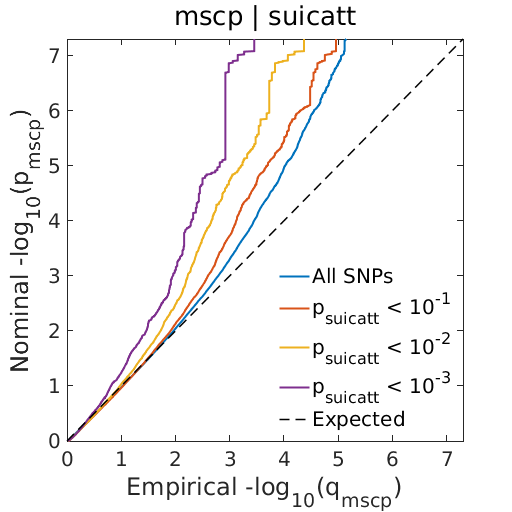
 B.
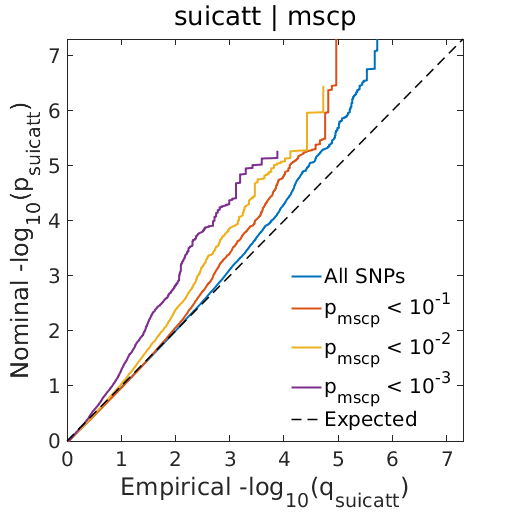

C.
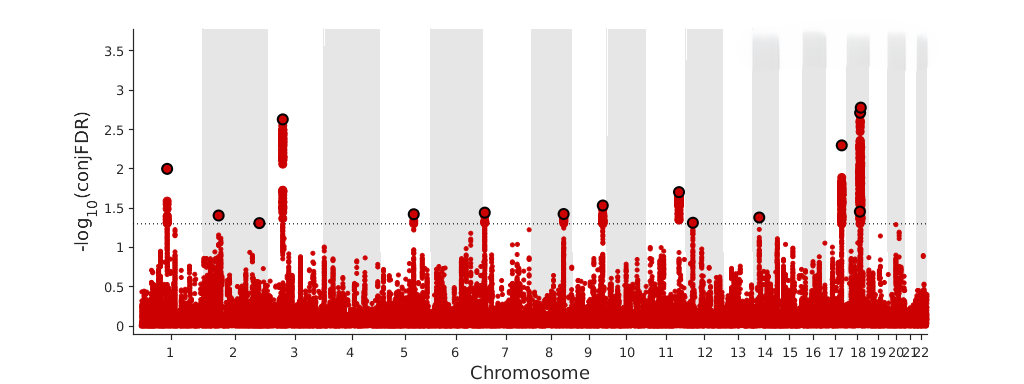

#### Supplementary Figure 20. SA-NEU.

A.
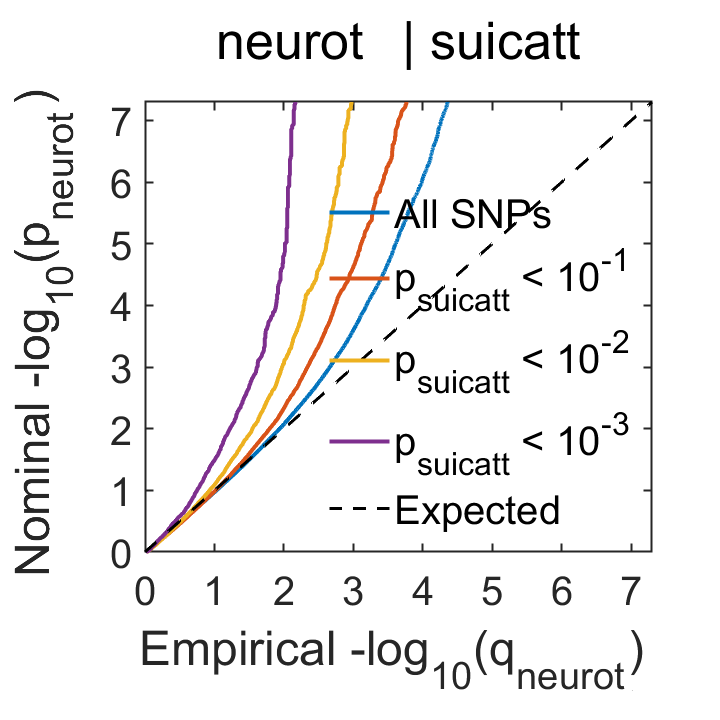
 B.
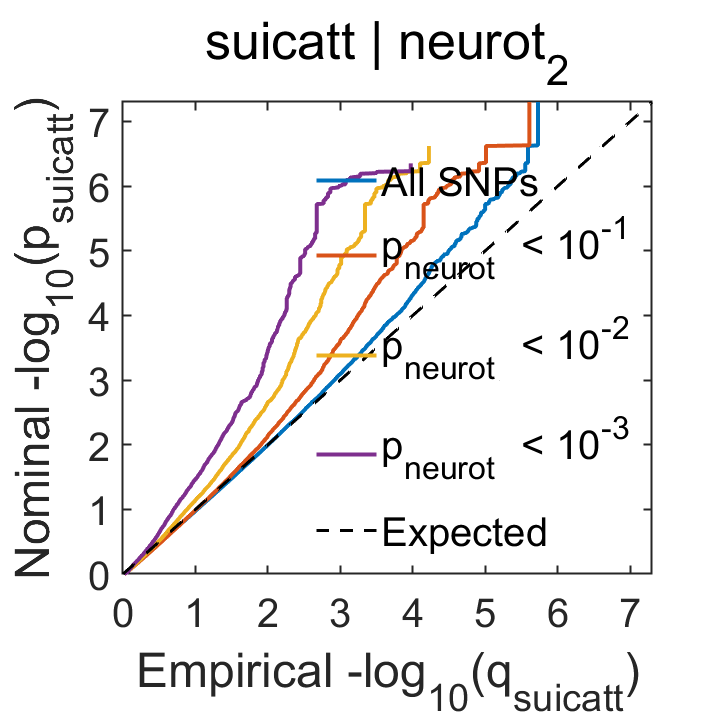

C.
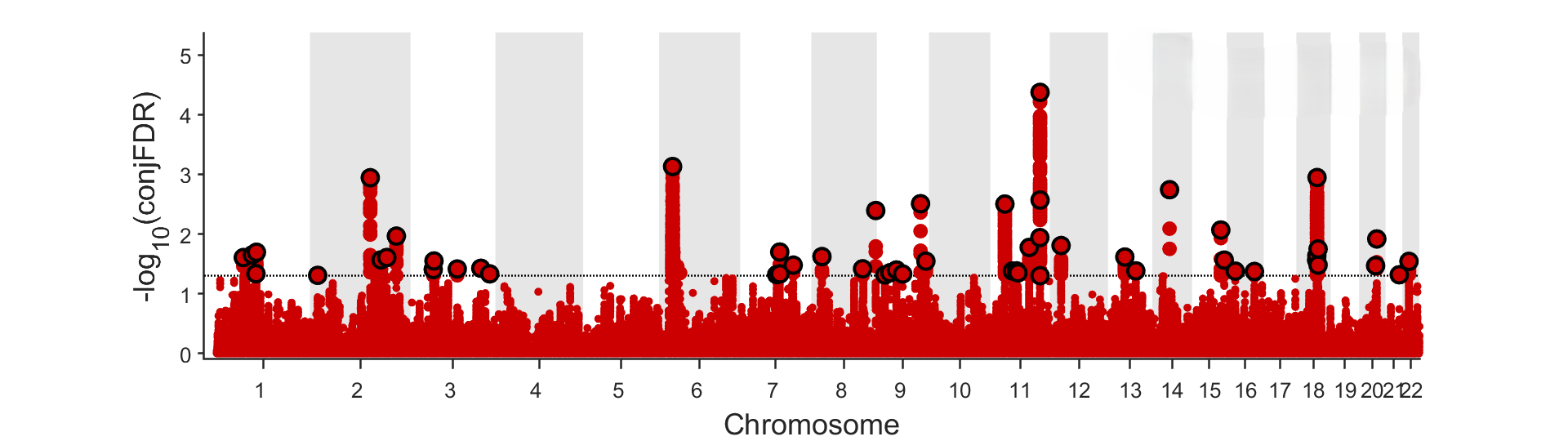

#### Supplementary Figure 21. SA-RSKT.

A.
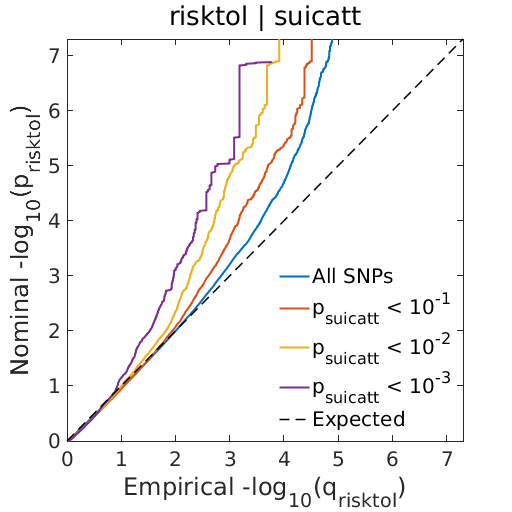
 B.
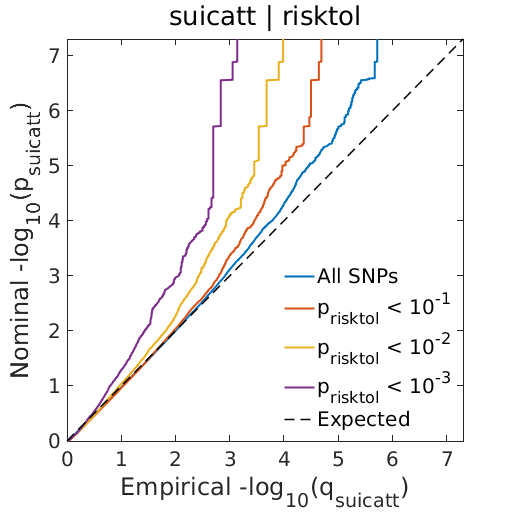

C.
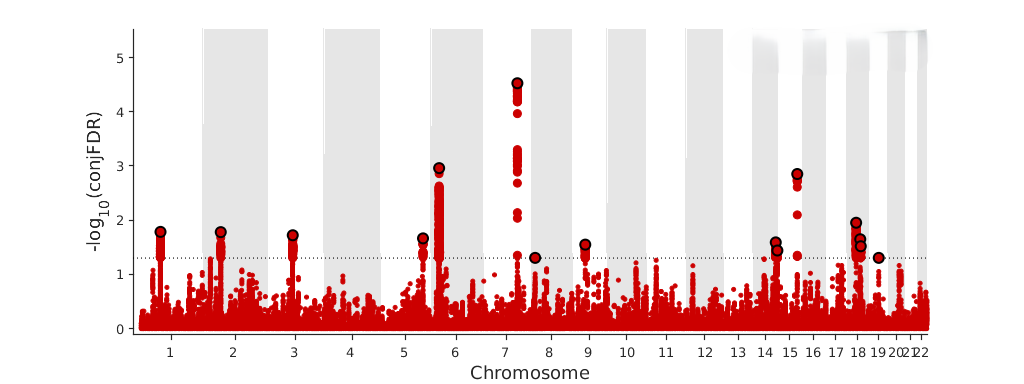

#### Supplementary Figure 22. SA-SCZ.

A.
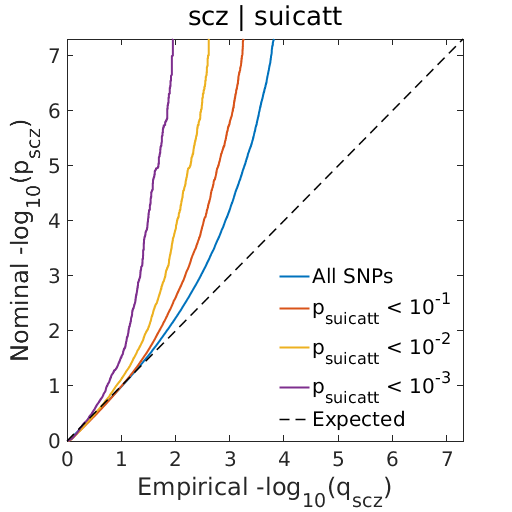
 B.
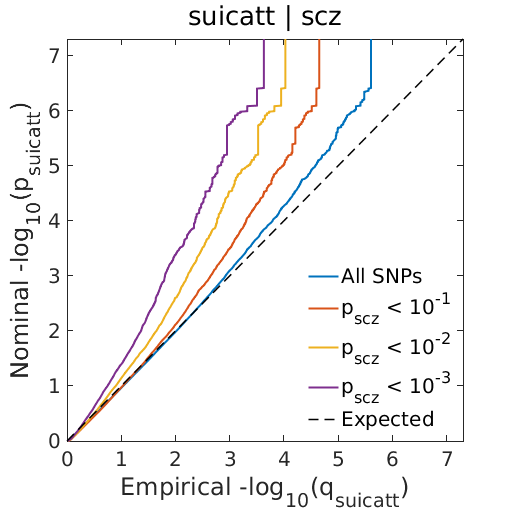

C.
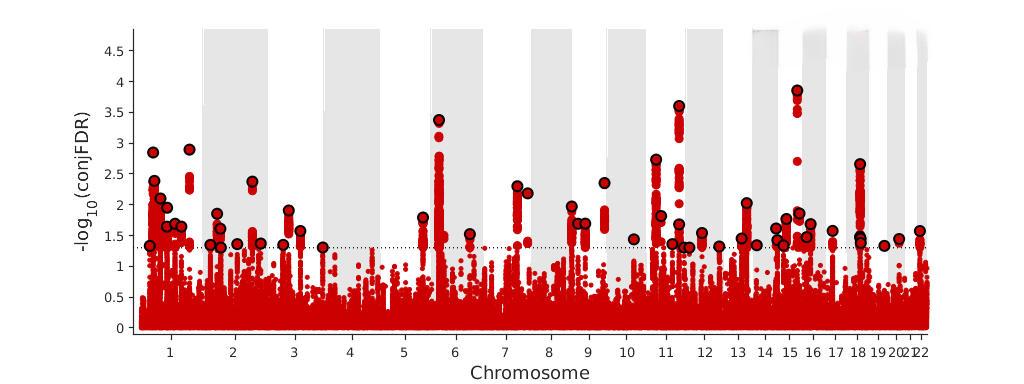

#### Supplementary Figure 23. SI-ADHD.

A.
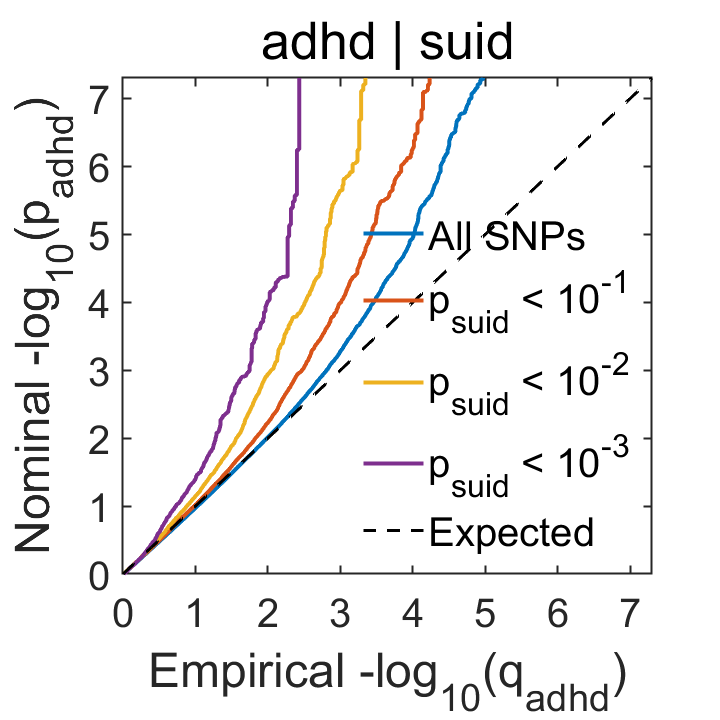
 B.
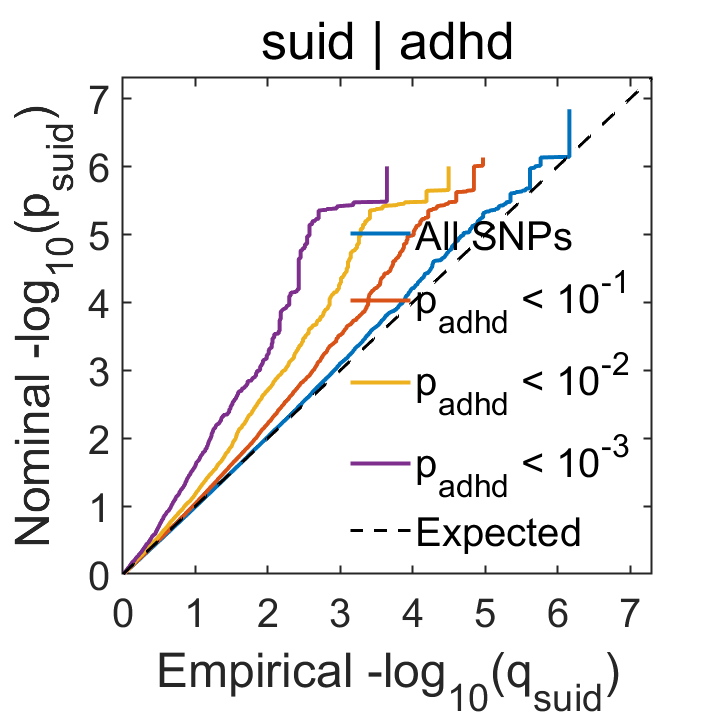

C.
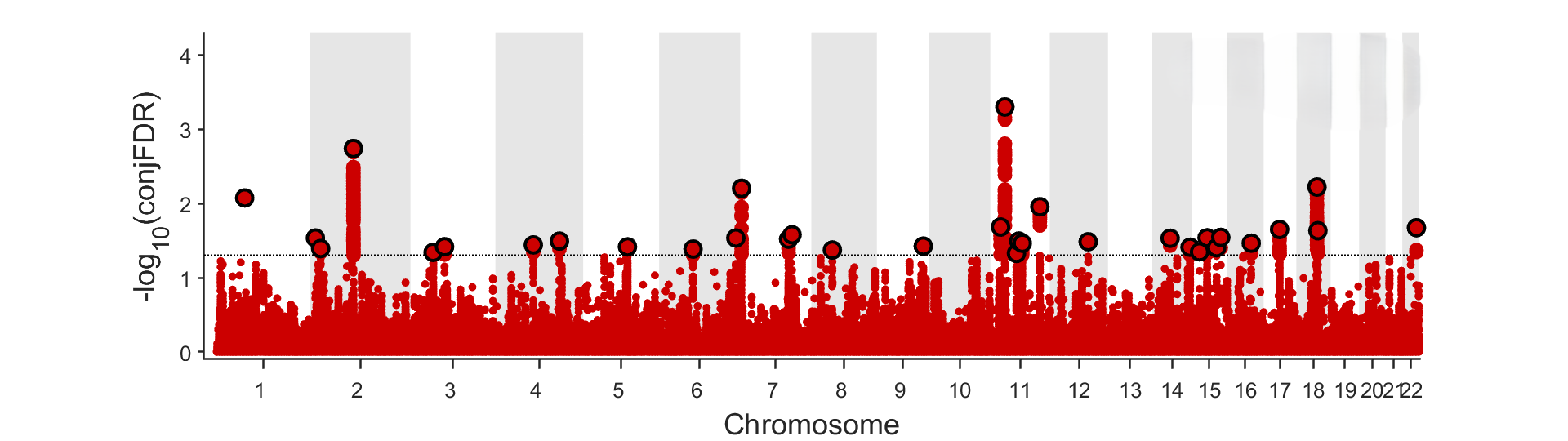

#### Supplementary Figure 24. SI-ASD.

A.
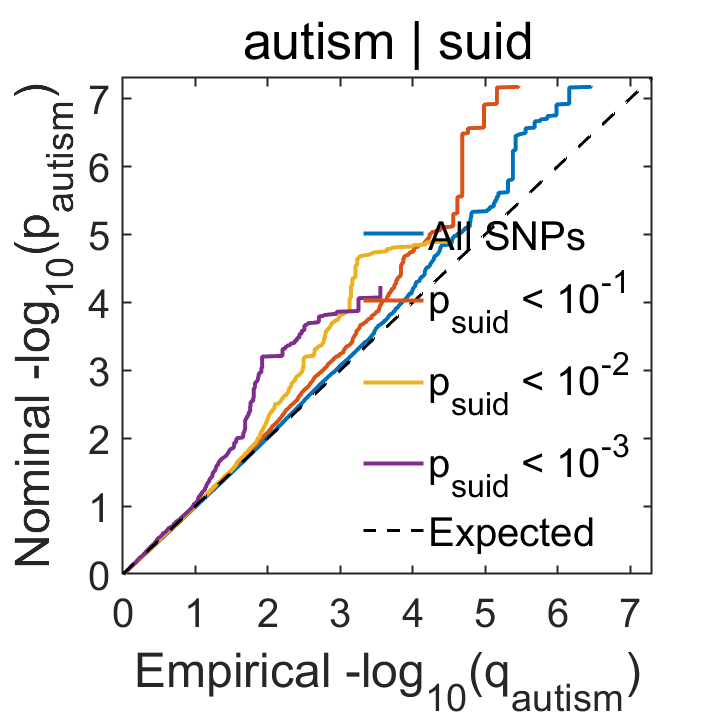
 B.
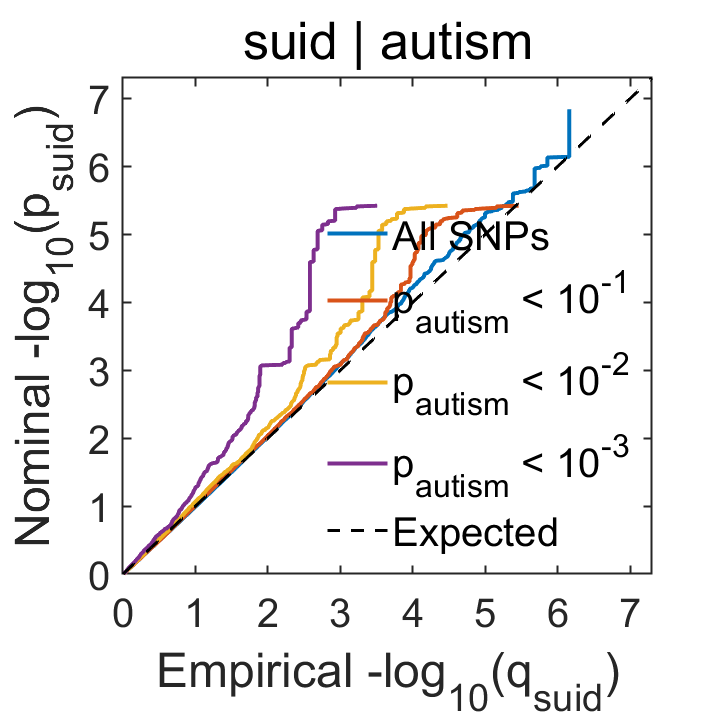

C.
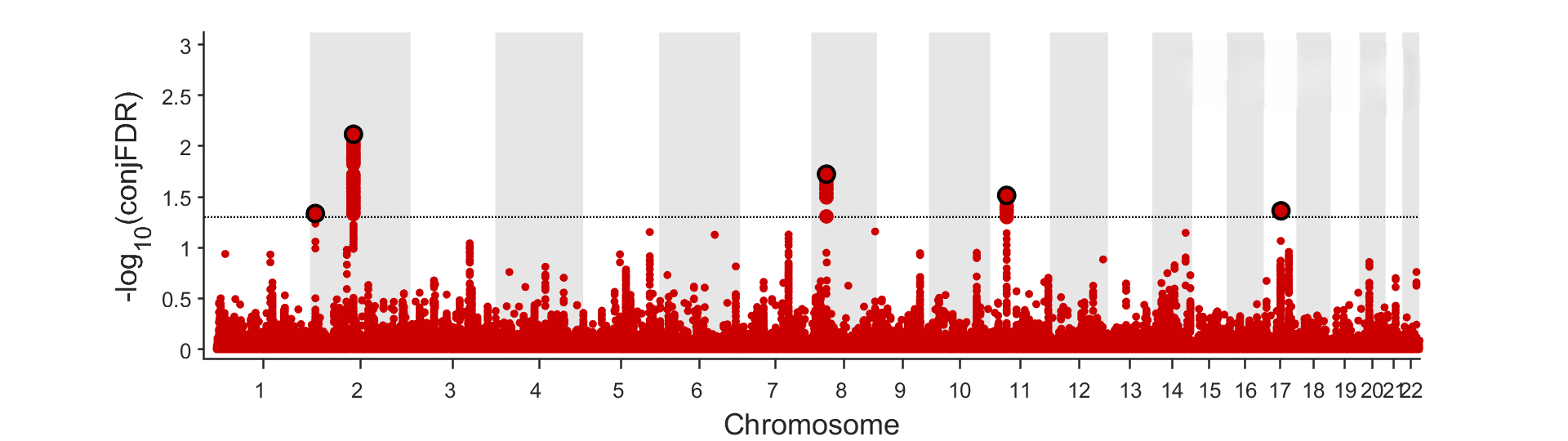

#### Supplementary Figure 25. SI-BMI.

A.
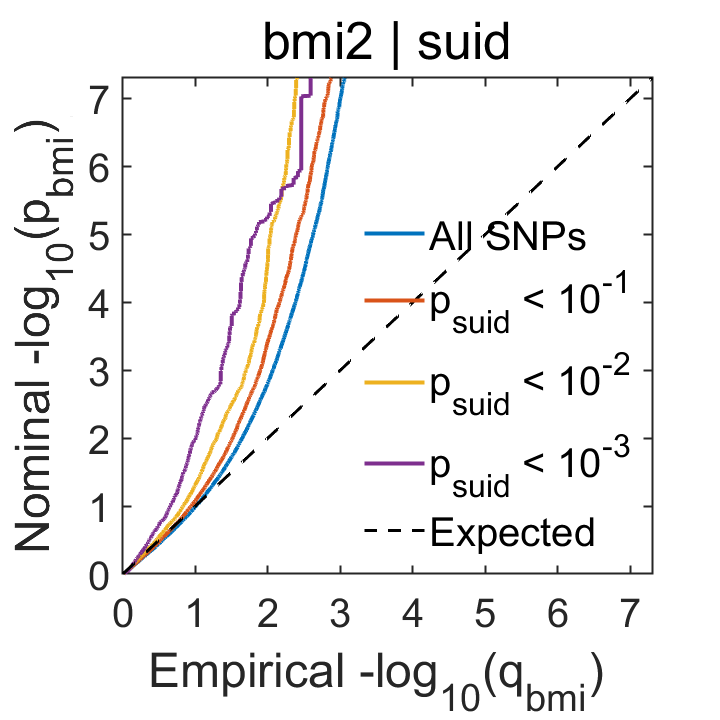
 B.
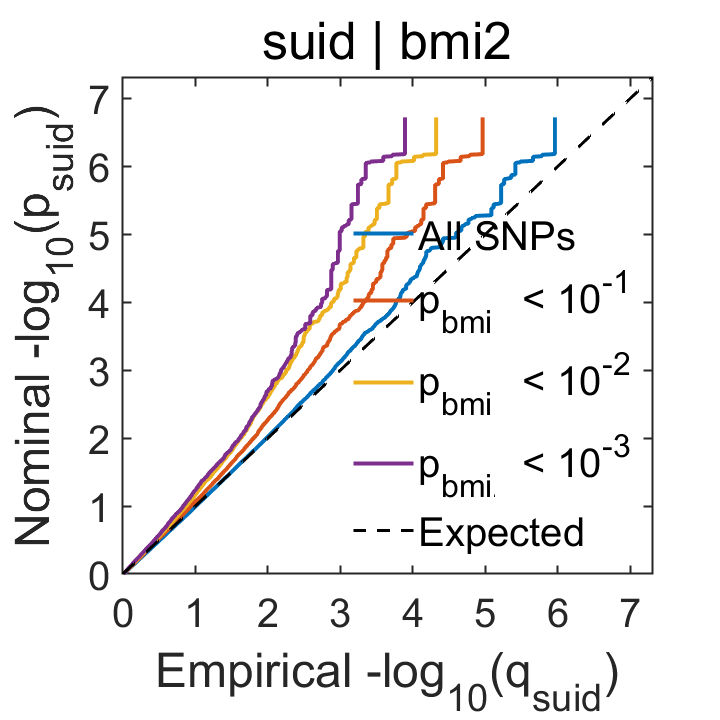

C.
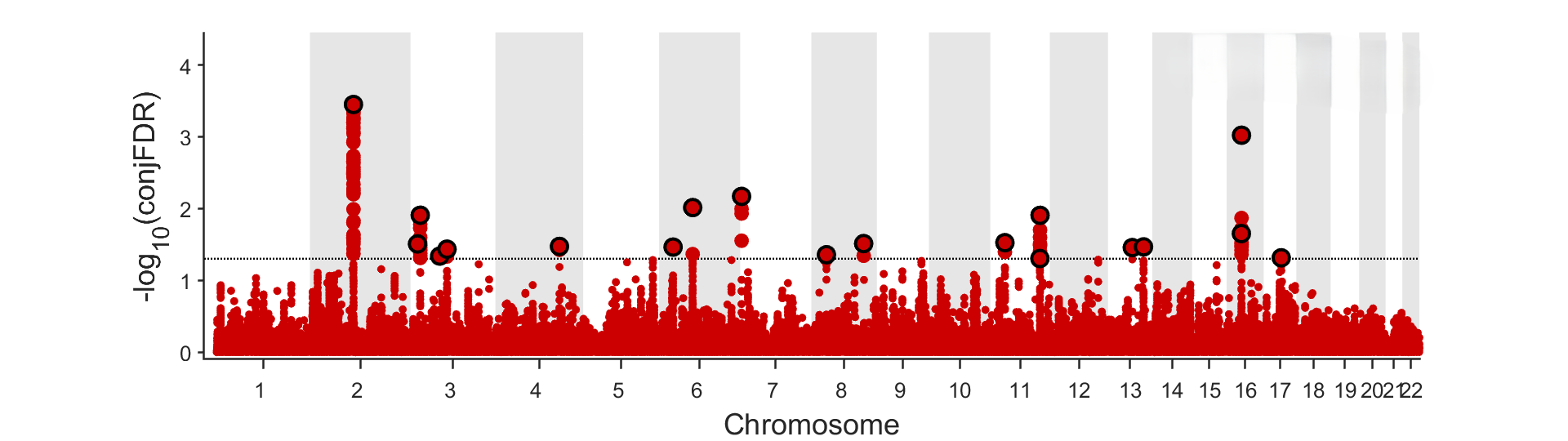

#### Supplementary Figure 26. SI-BP.

A.
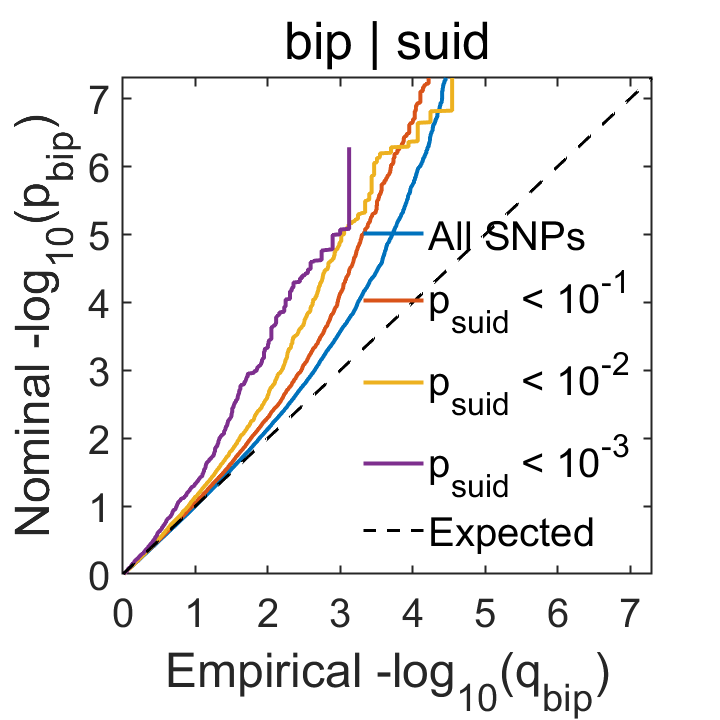
 B.
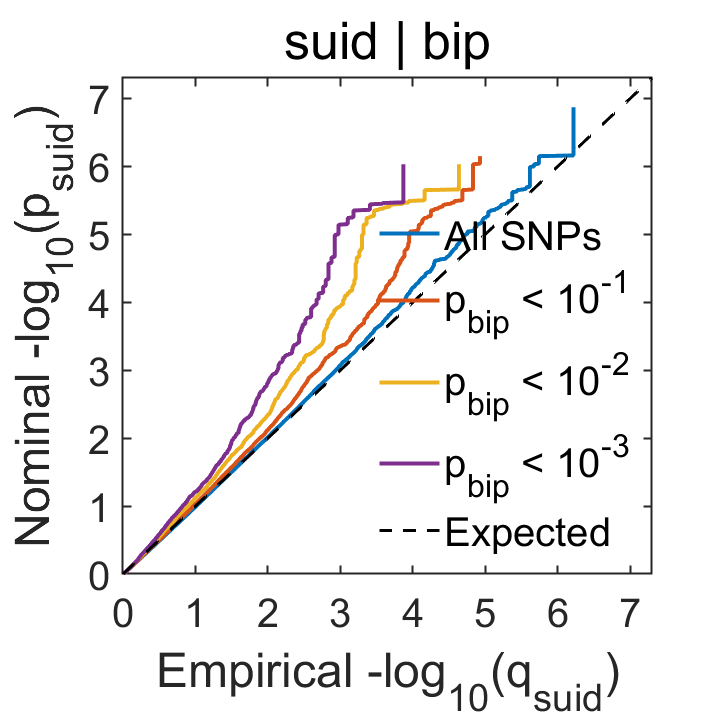

C.
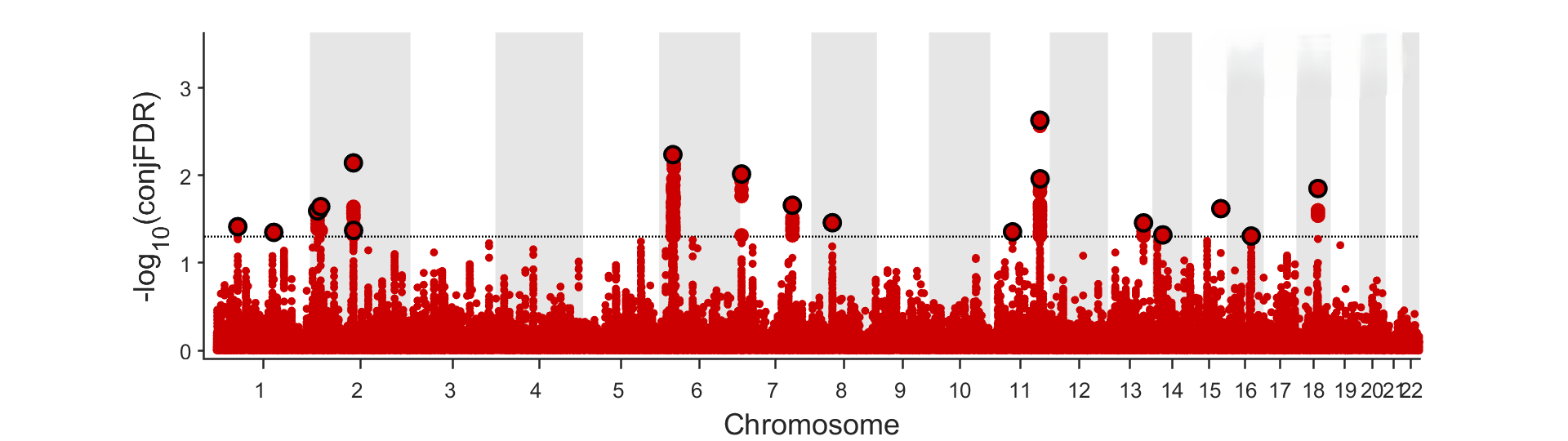

#### Supplementary Figure 27. SI-CPD.

A.
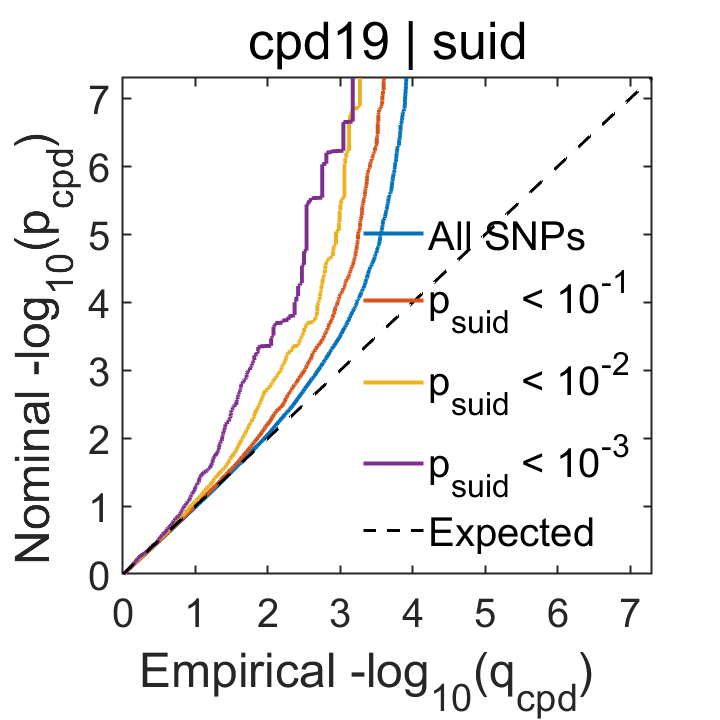
 B.
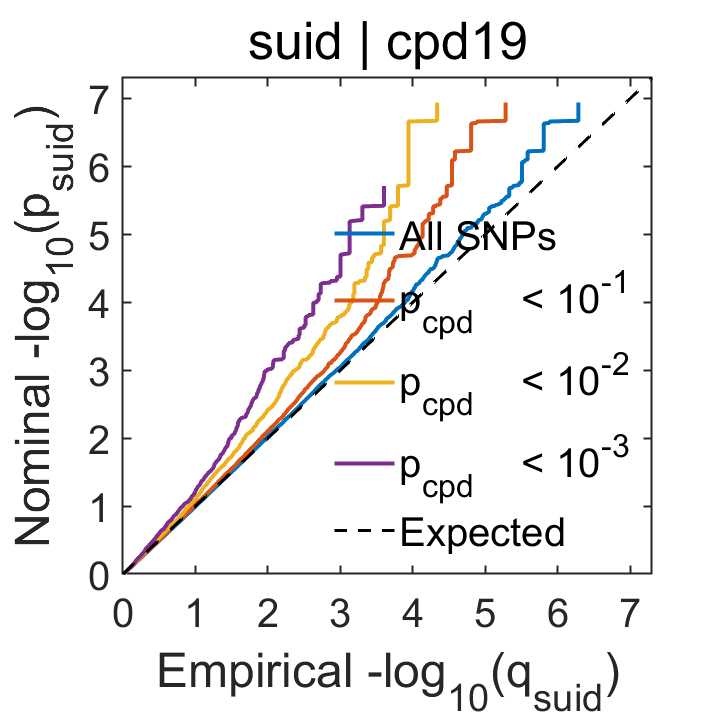

C.
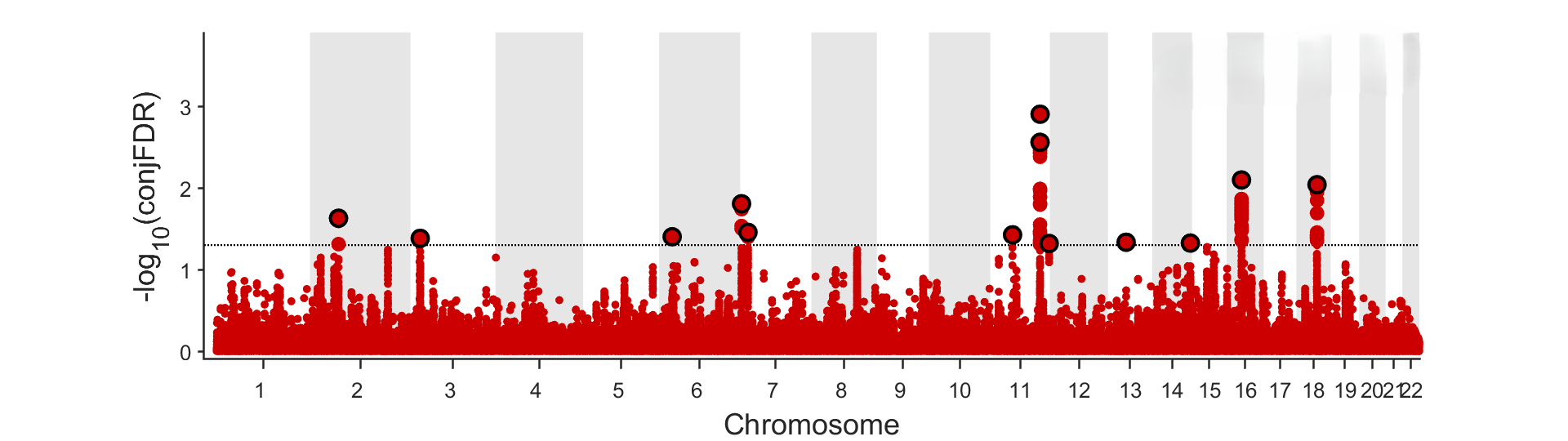

#### Supplementary Figure 28. SI-DSN.

A.
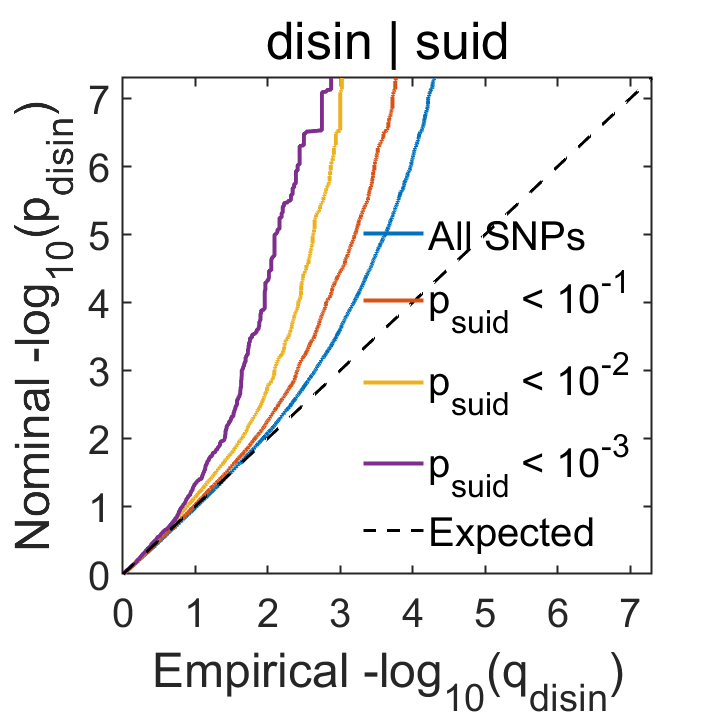
 B.
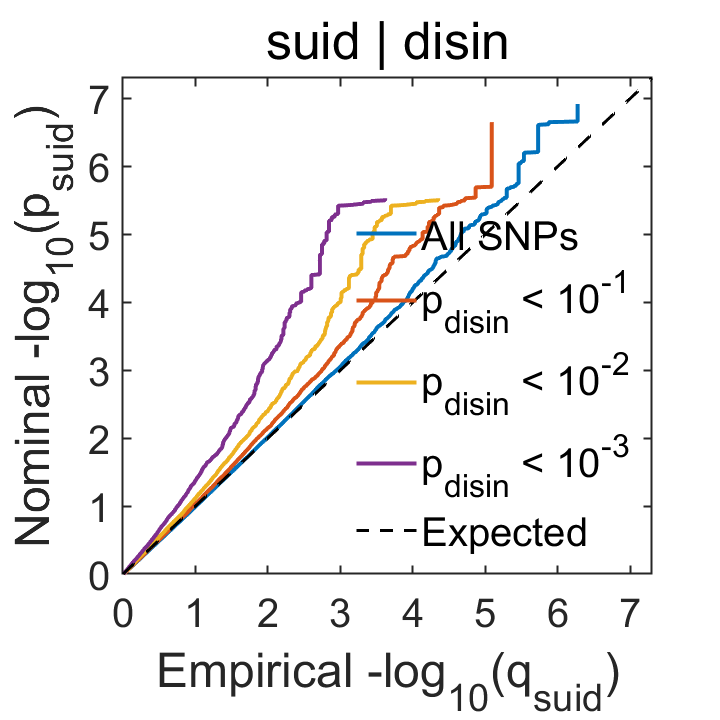

C.
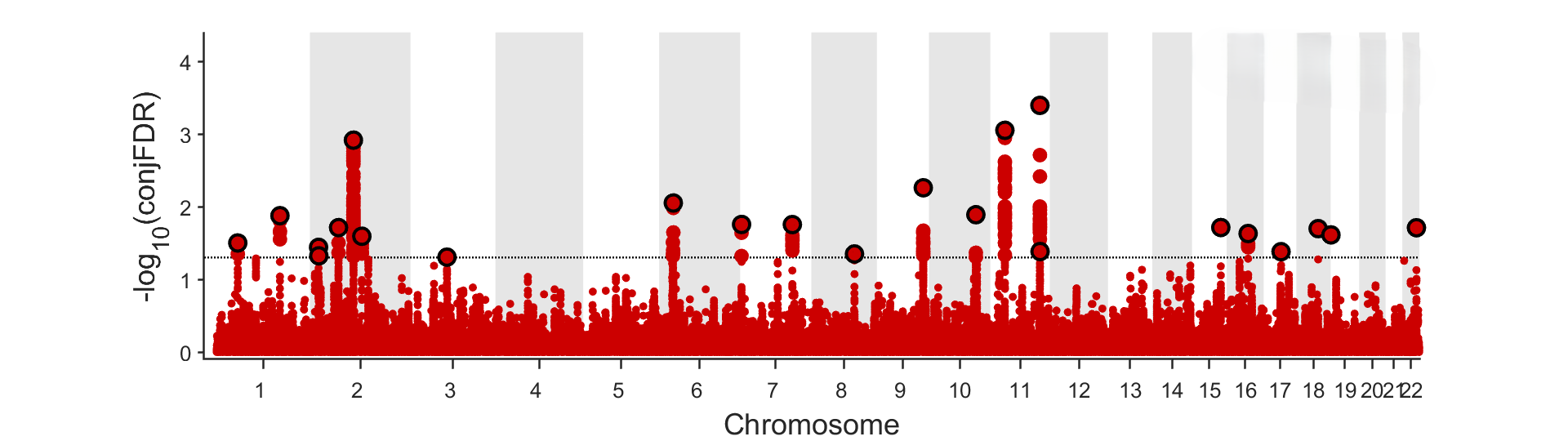

#### Supplementary Figure 29. SI-EA.

A.
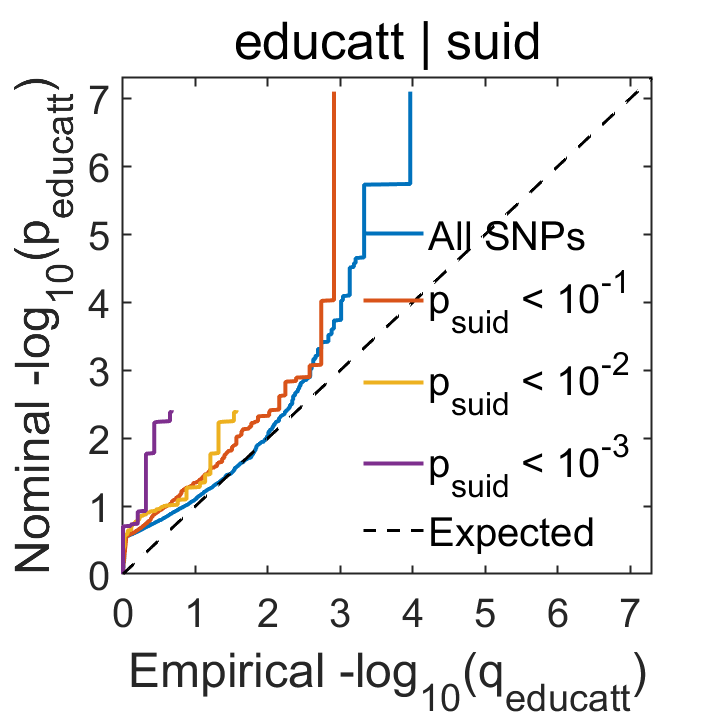
 B.
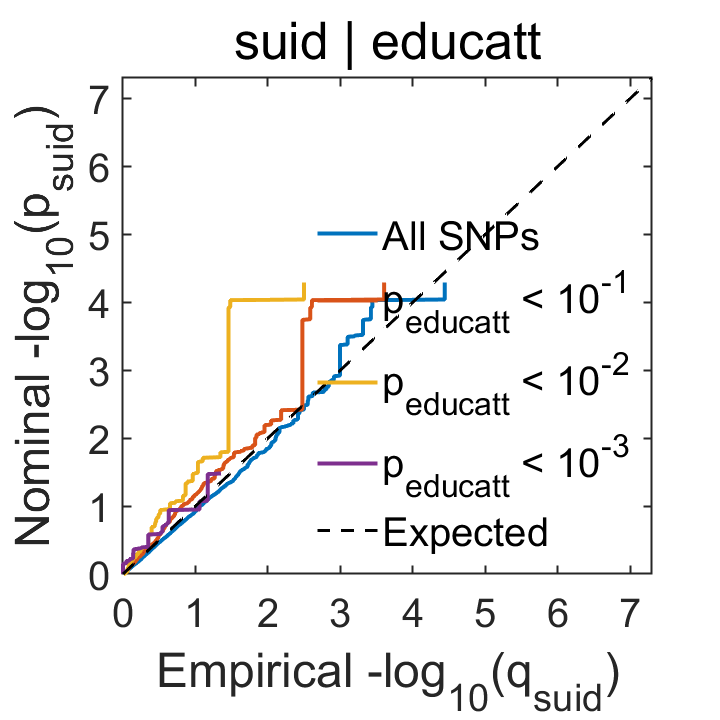

C.
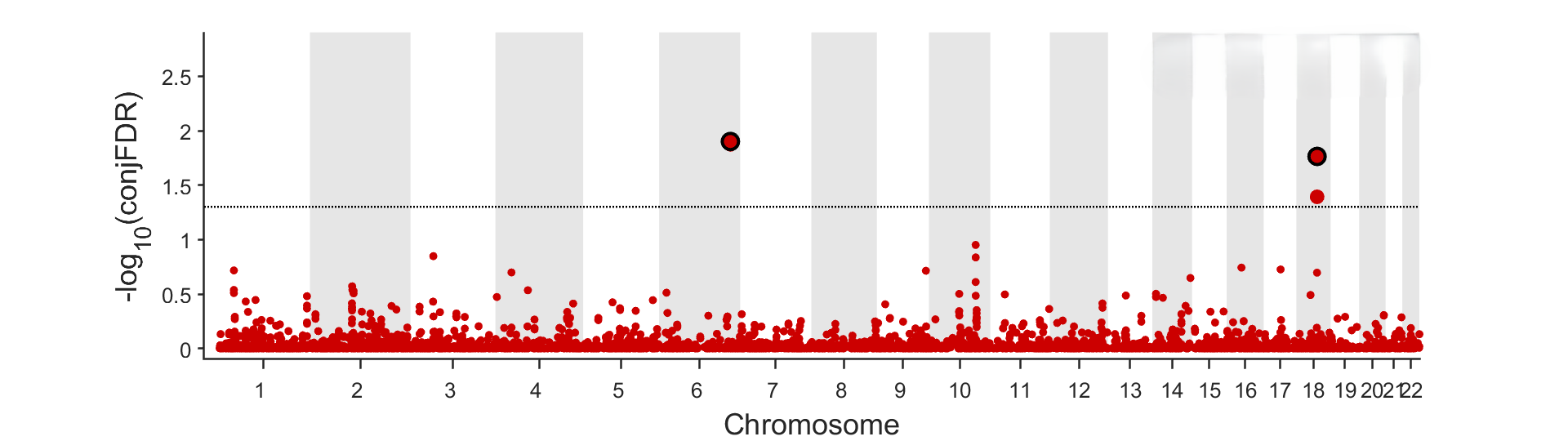

#### Supplementary Figure 30. SI-ESMK.

A.
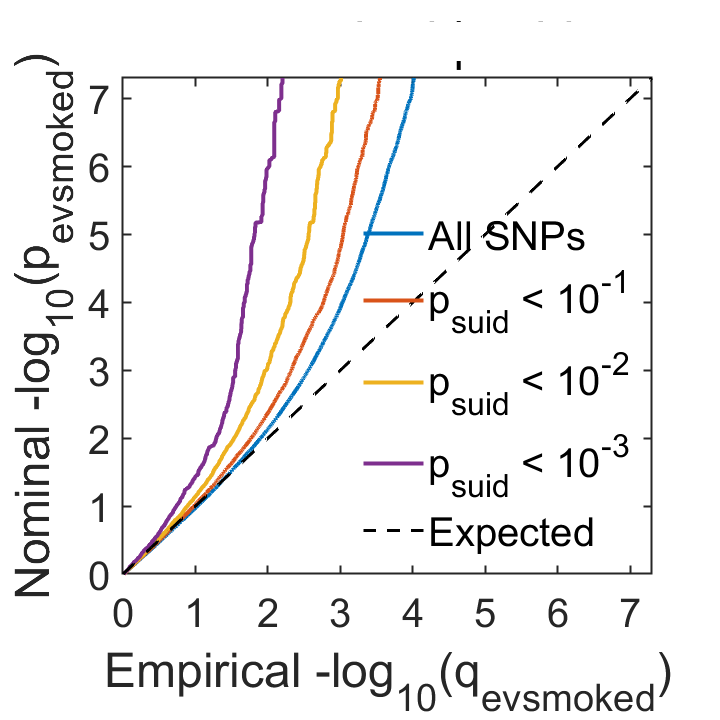
B.
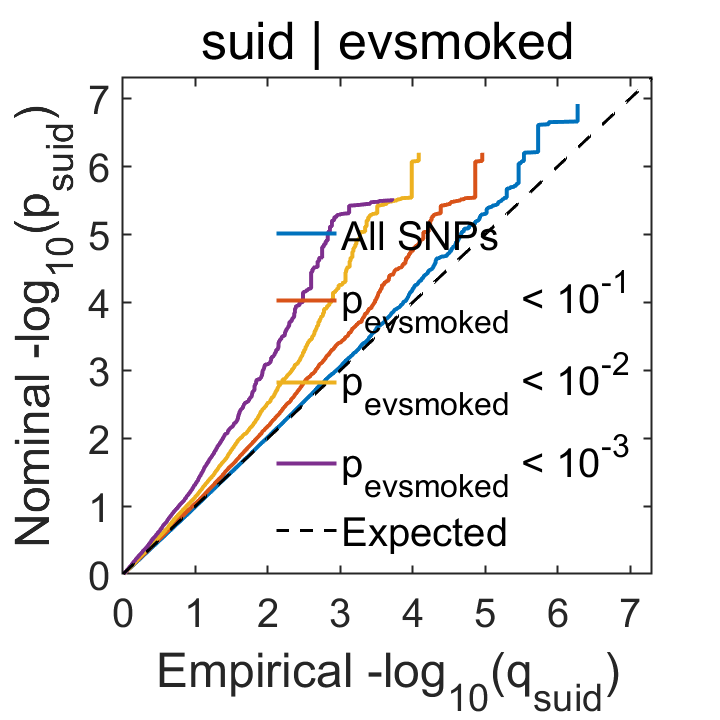

C.
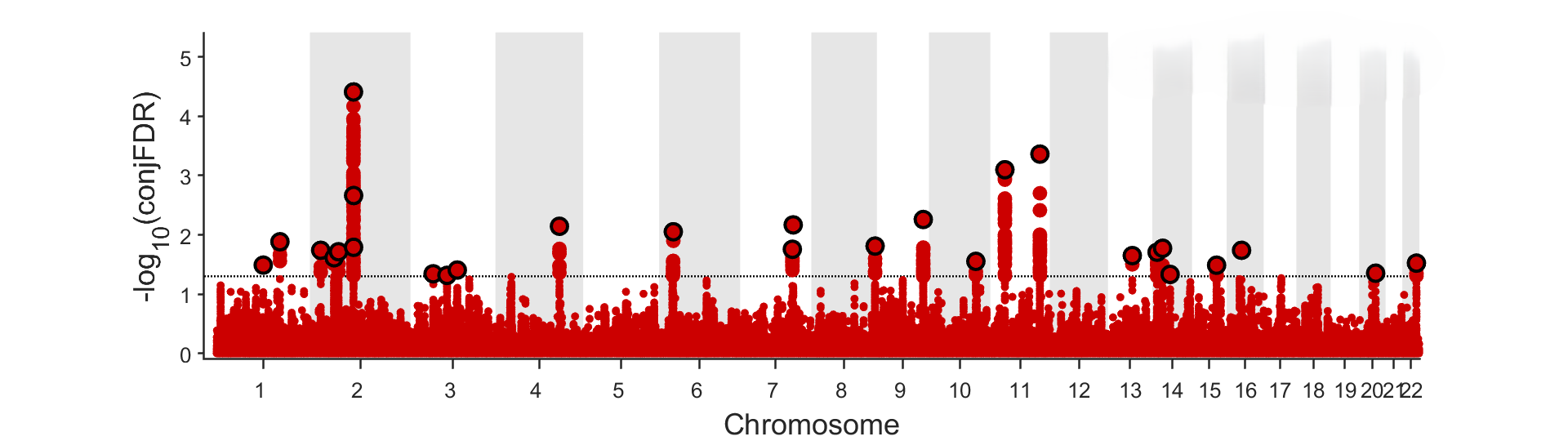

#### Supplementary Figure 31. SI-INS.

A.
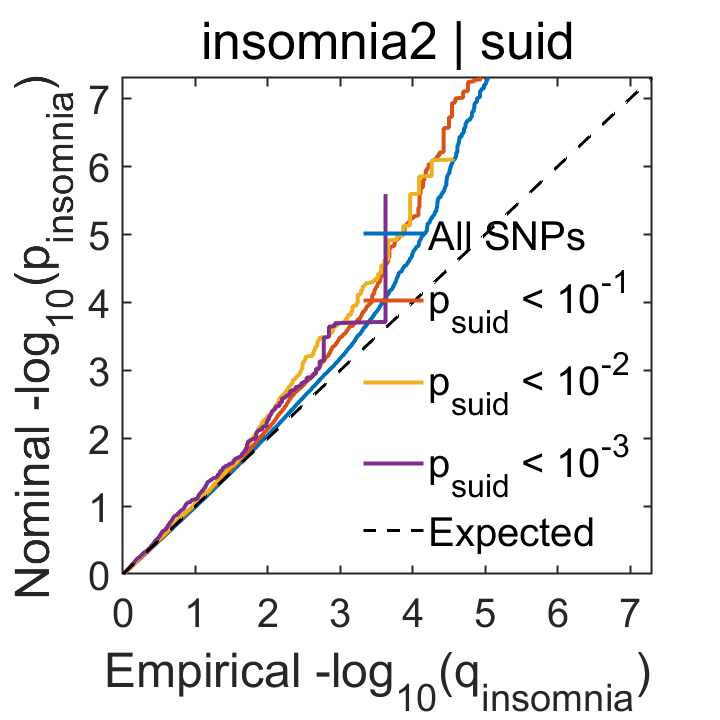
 B.
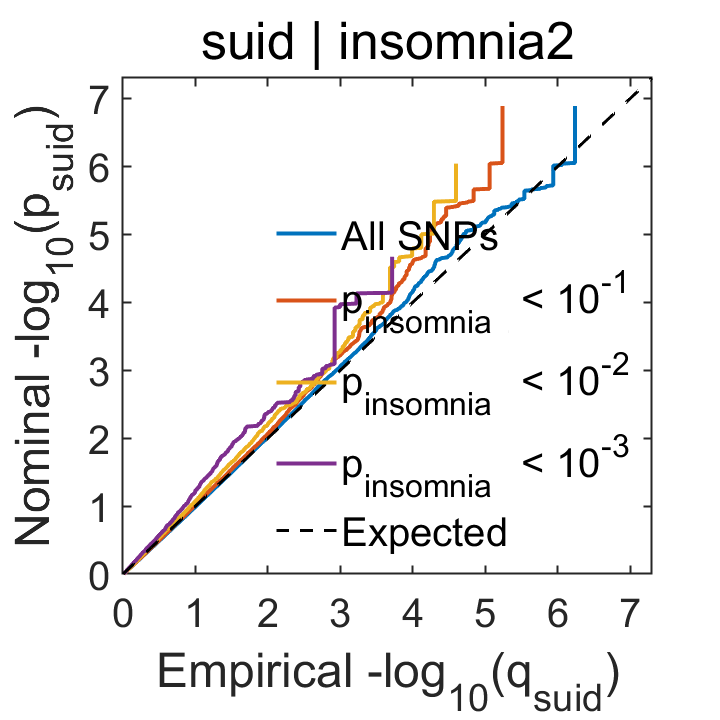

C.
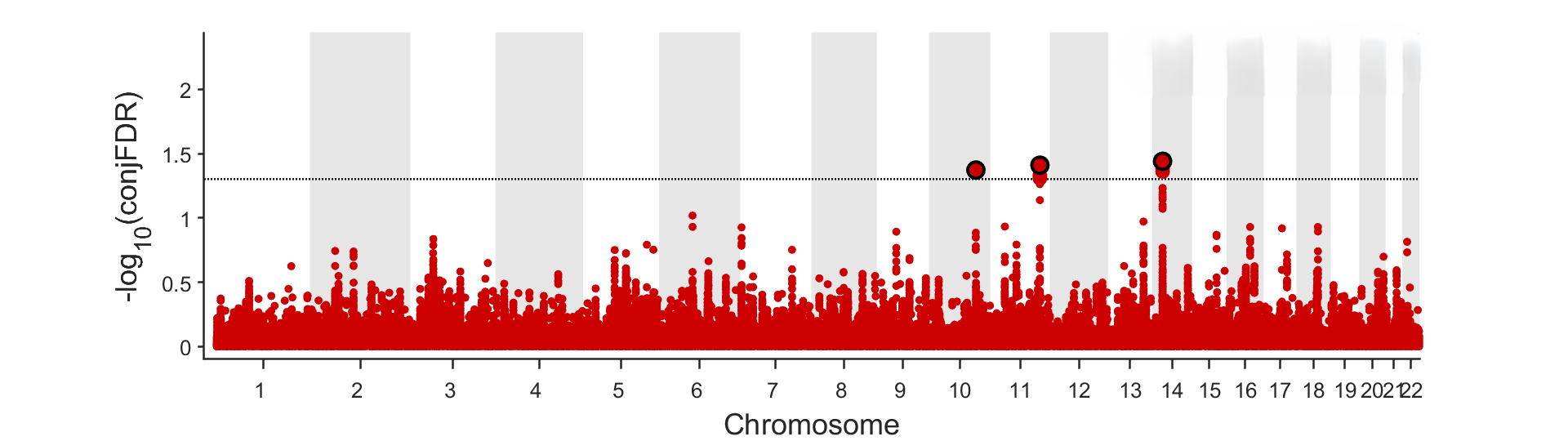

#### Supplementary Figure 32. SI-MSCP.

A.
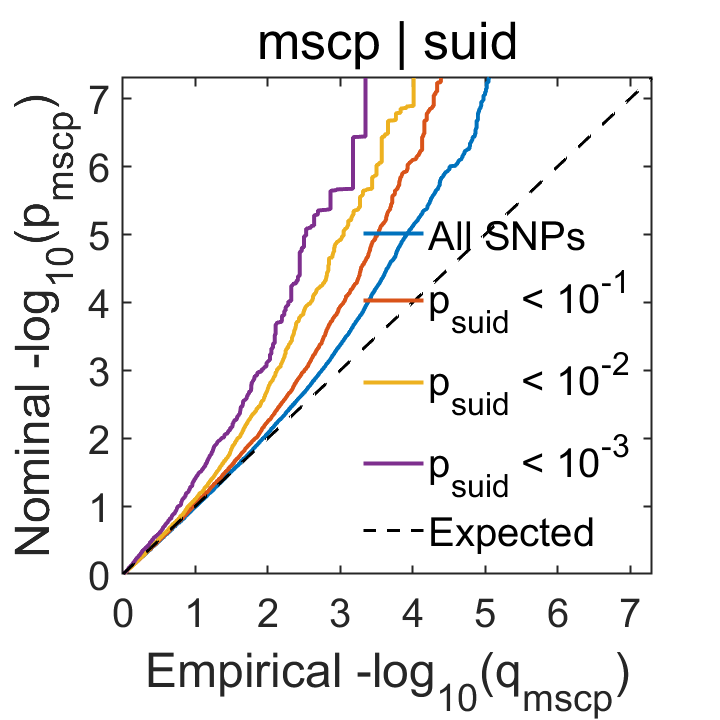
 B.
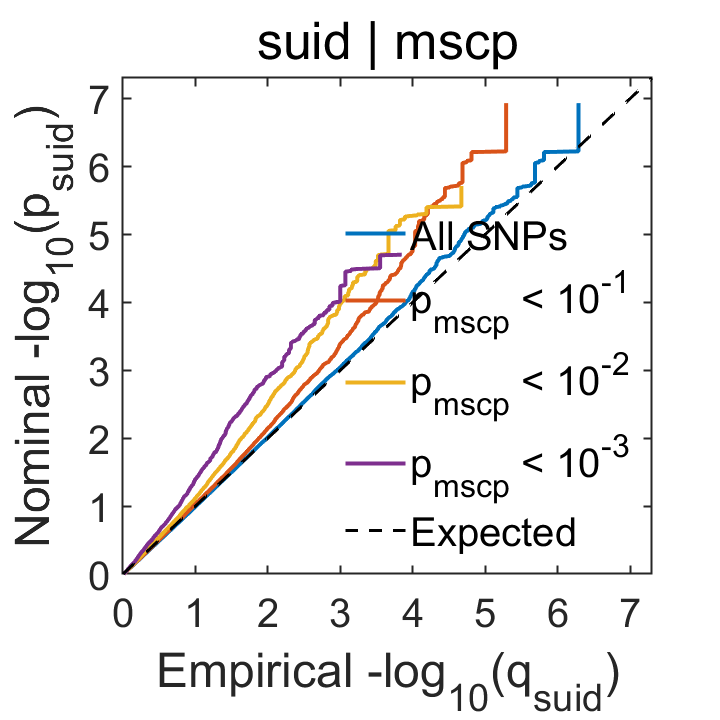

C.
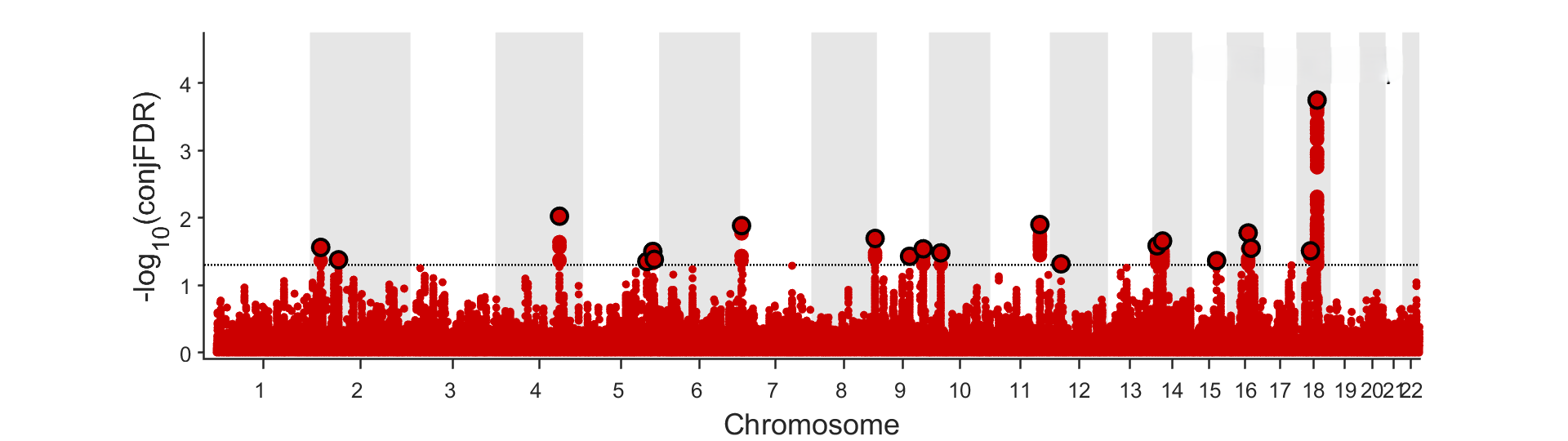

#### Supplementary Figure 33. SI-NEU.

A.
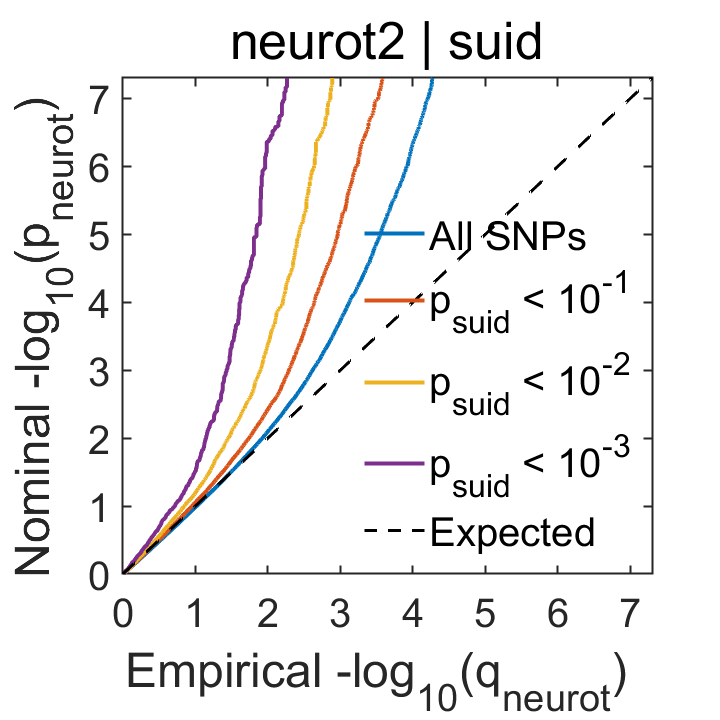
 B.
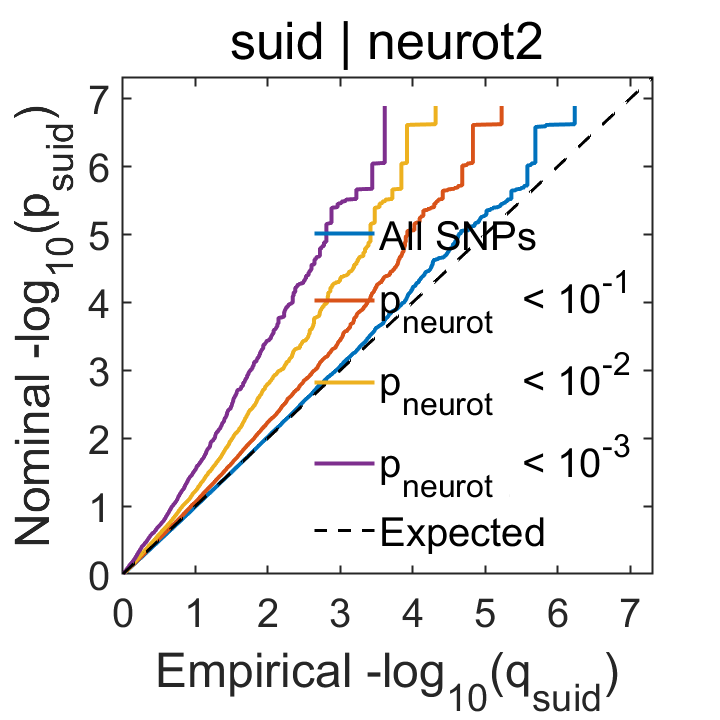

C.
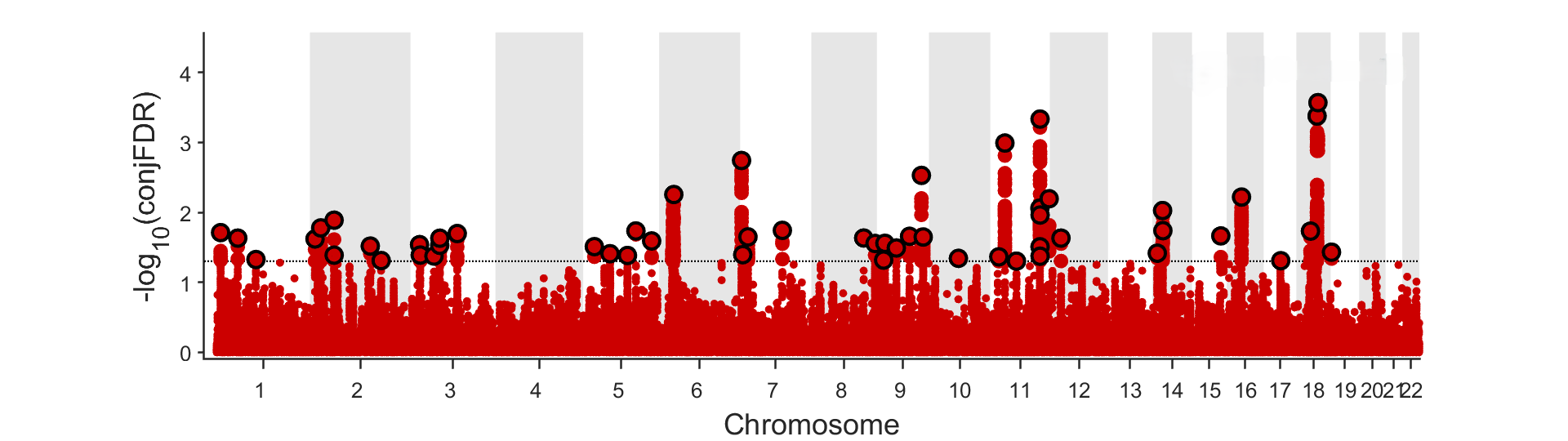

#### Supplementary Figure 34. SI-RSKT.

A.
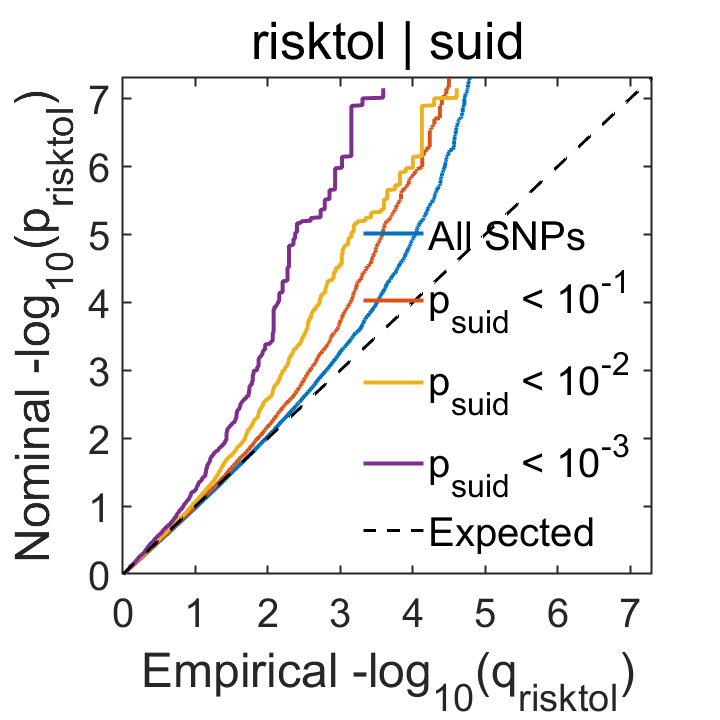
 B.
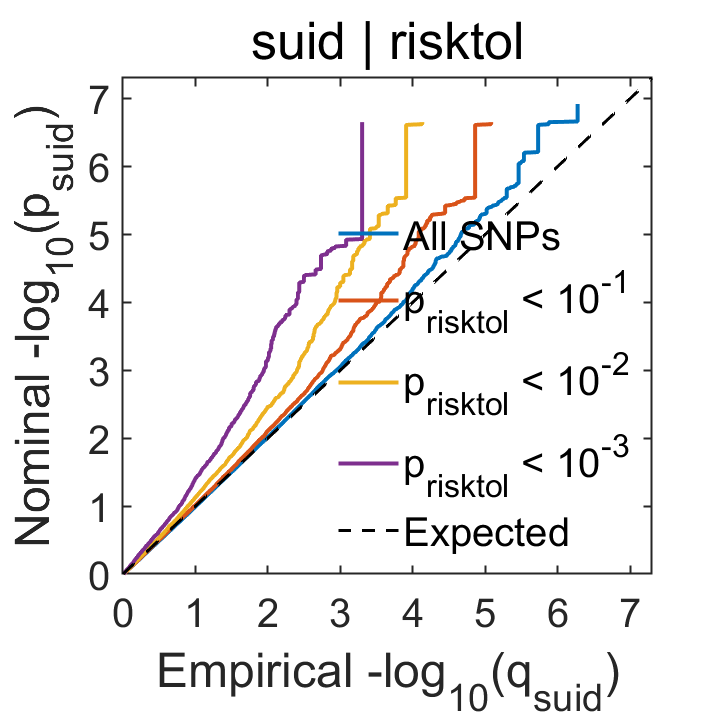

C.
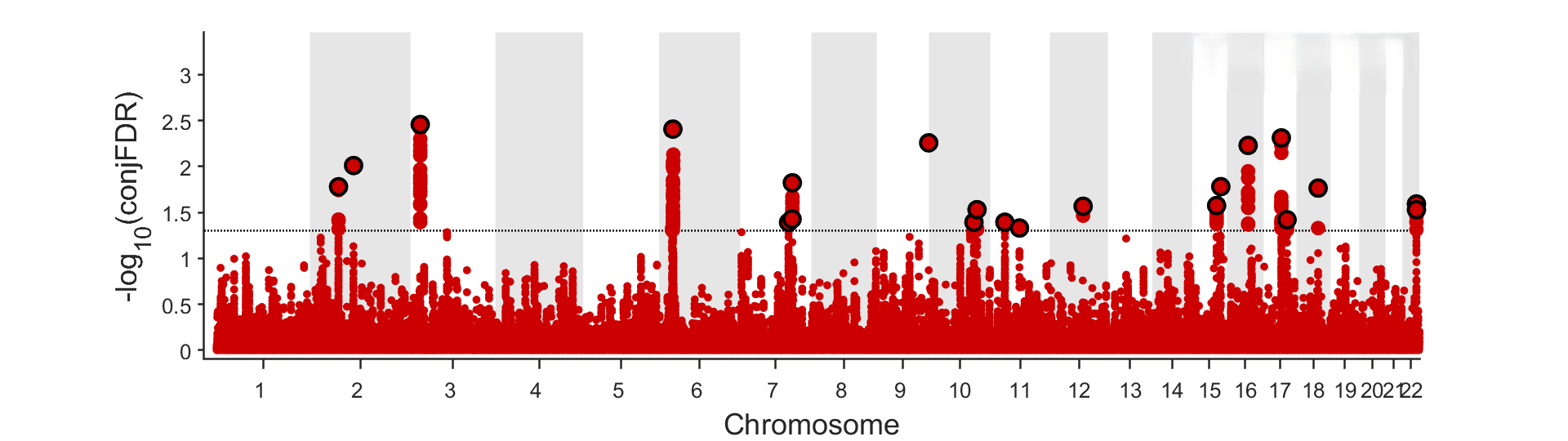

#### Supplementary Figure 35. SI-SCZ.

A.
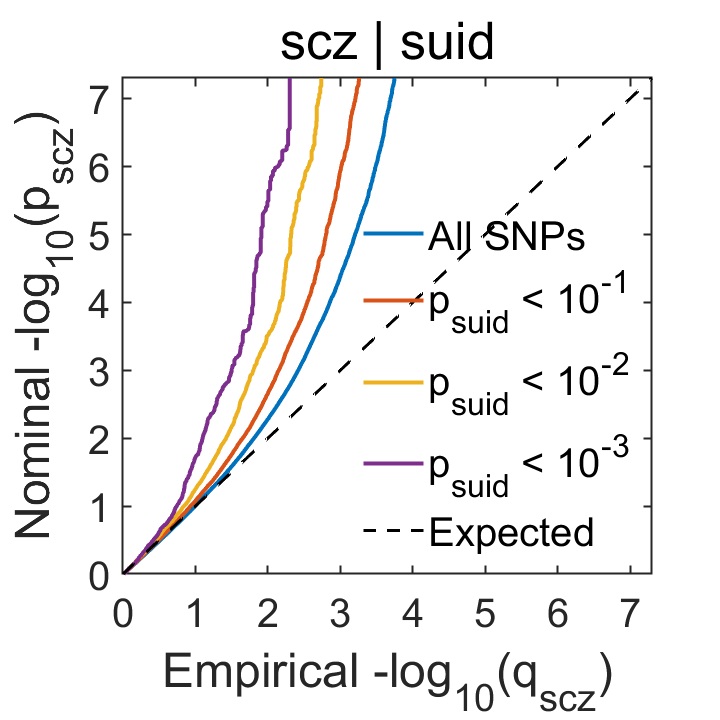
 B.
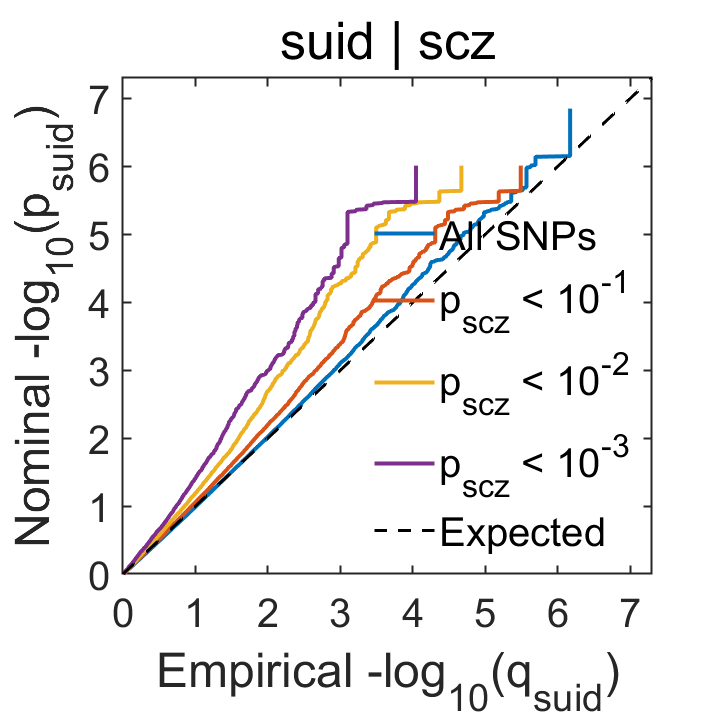

C.
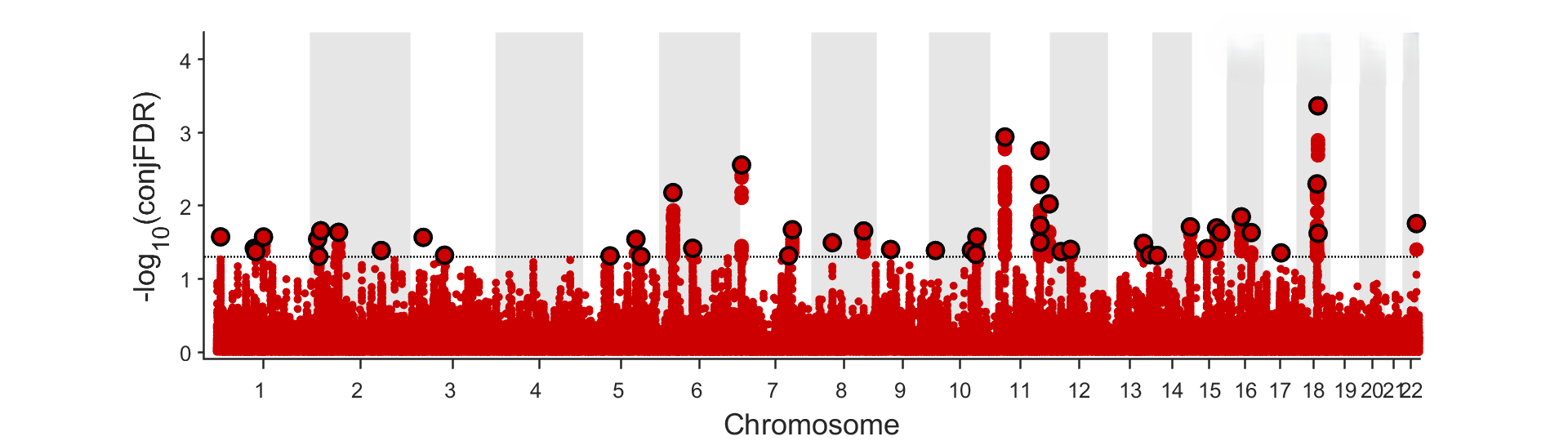
